# Examining inappropriate medication in UK primary care for type 2 diabetes patients with polypharmacy

**DOI:** 10.1101/2023.05.24.23290466

**Authors:** Maria Luisa Faquetti, Géraldine Frey, Dominik Stämpfli, Stefan Weiler, Andrea M. Burden

## Abstract

**Aims:** To estimate the prevalence of potentially inappropriate prescriptions (PIPs) in patients starting their first non-insulin antidiabetic treatment (NIAD) using two explicit process measures of the appropriateness of prescribing in UK primary care, stratified by age and polypharmacy status.

**Methods:** A descriptive cohort study between 2016 and 2019 was conducted to assess PIPs in patients aged ≥45 years at the start of their first NIAD, stratified by age and polypharmacy status. The American Geriatrics Society (AGS) Beers criteria 2015 was used for older (≥65 years) and the Prescribing Optimally in Middle-age People’s Treatments (PROMPT) criteria for middle-aged (45-64 years) patients. Prevalence of overall PIPs and individual PIPs criteria was reported using the IQVIA Medical Research Data incorporating THIN, a Cegedim Database of anonymised electronic health records in the UK.

**Results:** Among 28,604 patients initiating NIADs, 18,494 (64.7%) received polypharmacy. In older and middle-aged patients with polypharmacy, 39.6% and 22.7%, respectively, received ≥1 PIPs. At the individual PIPs level, long-term PPI use and strong opioid without laxatives were the most frequent PIPs among older and middle-aged patients with polypharmacy (11.1% and 4.1%, respectively).

**Conclusions:** This study revealed that patients starting NIAD treatment receiving polypharmacy have the potential for pharmacotherapy optimisation.

## 1 Introduction

Type 2 diabetes mellitus (T2DM) is a complex chronic metabolic disorder affecting approximately 537 million adults worldwide, with nearly 90% having T2DM.[1] T2DM is characterised by insulin resistance and progressive decline of insulin production by pancreatic β-cell dysfunction.[2] While T2DM incidence rises with age, rates of T2DM onset in younger patients are increasing, leading to new public health challenges.[3] Pharmacological management of individuals with T2DM is often complex, especially when concomitant chronic conditions and diabetes complications are present, which may result in the use of multiple medications, also known as polypharmacy.[4] Moreover, patients with multimorbidity may attend multiple physicians, leading to fragmented, uncoordinated medication prescriptions.

Polypharmacy, generally defined as the concomitant use of five or more drugs, may be required to manage multiple coexisting conditions, presenting a risk of both undertreatment and overtreatment.[5] Although the correct combination of drugs in patients with T2DM can improve their health status and quality of life, polypharmacy increases the likelihood of potentially inappropriate prescriptions (PIPs), which can thereby increase the risk of experiencing an adverse drug event (ADE), drug-drug interaction (DDI), or drug-disease interaction.[6,7]

Furthermore, patients and society expect the benefits of medicines to outweigh their risks. However, patients are not equally receptive to beneficial effects and not equally susceptible to ADE. Factors such as comorbidities, concurrent prescribing of drugs with potential for interaction, and inappropriate prescribing can all contribute to this variability.[8] Therefore, it is crucial to ensure that medications are prescribed appropriately, considering individual patients’ situations, to minimise the risks and maximise the benefits.

The appropriateness of prescribing medication can be assessed by using implicit process measures (i.e., judgement based, focused on individual patients) or by using explicit process measures (i.e., criterion-based). Among the most commonly used explicit process measures are the American Geriatrics Society (AGS) Beers criteria[9] and the Prescribing Optimally in Middle-age People’s Treatments (PROMPT),[10] which were tailored to identify drugs to be avoided in older and middle-aged adults, respectively, independent of a diagnosis or in the context of a disease. While previous studies, predominantly among the elderly, have identified that the prevalence of PIPs is between 20% and 79%, depending on the population and setting,[11–16] less is known among patients with T2DM, particularly non-elderly.

The study by Oktora and colleagues revealing that the prevalence of ≥1 PIP among patients aged 45+ years with T2DM and polypharmacy using Beers and PROMPT ranged from 24.9% to 39.0% was limited to information on drug prescriptions, which hindered implementation of most of the criteria.[14] Thus, no large population-based study encompassing clinical information has applied these measures to identify drugs to be avoided in the context of diseases or under specific situations in patients with T2DM.

The overall aim of this study was to estimate the prevalence of PIPs in middle-aged and older adults starting their first non-insulin antidiabetic treatment (NIAD) using two explicit process measures of appropriateness of prescribing medication in a primary care database from the UK, stratified by age and polypharmacy status. This study aimed to identify if pharmacotherapy could be optimised and, ultimately, to provide clinical insights into the management of patients with T2DM receiving multiple medications in primary care.

## 2 Materials and Methods

### 2.1 Study design and data source

We conducted a descriptive cohort study using data from primary care practices in the UK between 2016 and 2019 to assess PIPs in patients at the start of their first NIAD, stratified by age and polypharmacy status. The data were obtained from IQVIA Medical Research Data (IMRD; incorporating data from THIN, A Cegedim Database of anonymised electronic health records in the UK). IMRD is an extensive database containing longitudinal non-identified electronic medical records collected from general practices in the UK from over 18 million patients, of which approximately 2.9 million are currently active. IMRD has been shown to be representative of the UK and valid in terms of age and sex comparisons and a wide range of diseases.[17] Demographic information, clinical outcomes and diagnosis, lab tests (e.g., estimated glomerular filtration rate [eGFR] and haemoglobin A1c [HbA1c]), drug prescriptions, specialist referrals, and other information (e.g., smoking status, pregnancy, and death) are recorded in the database by general practitioners (GPs). Medications are recorded using the British National Formulary (BNF) classification and were manually mapped to their corresponding International Anatomical Therapeutic Classification (ATC) code. As the IMRD consists of primary care records, medication dispensed during hospitalisation, nor those prescribed by specialists, are included in the database. All diagnoses are recorded using Read codes.[17]

### 2.2 Study population

The study population consisted of all adults aged ≥45 years who received a first-ever NIAD prescription between January 2016 and December 2019. Cohort entry (index date) was defined as the date of the first-ever NIAD prescription. Patients included were classified by age at index date as older adults (≥65 years) or middle-aged adults (45-64 years). Patients were allowed to have multiple co-prescribed NIADs at the index date. All patients were required to have ≥1 year of database history and at least one GP visit before the index date. We only considered patients registered with a practice with a valid up-to-standard date, who had valid entries (i.e., a valid patient flag), and with a first NIAD prescription with a valid flag in the database. Patients diagnosed with gestational diabetes, polycystic ovary syndrome, cancer (except by non-melanoma skin cancers), or insulin therapy previous to, or at, the index date were excluded. Additionally, patients in palliative care or at end-of-life were excluded. All comorbidity or palliative/end-of-life care patients were identified using Read codes.

### 2.3 Polypharmacy definition

We leveraged an existing cohort of patients with T2DM and polypharmacy, which is described in detail elsewhere.[18] In brief, polypharmacy was defined as the prescription of ≥5 different drug compounds on or within 90 days before the index date. We identified patients with polypharmacy at the start of NIAD therapy and only considered drugs and diagnoses (i.e., history of comorbidities and medical conditions in the Beers or PROMPT criteria for assessing PIP) with a valid flag in the database. Additional details on the selection of drugs for the polypharmacy analysis are available in Supplementary Information.

### 2.4 Potentially inappropriate prescriptions (PIPs)

We used the AGS Beers criteria for assessment of PIPs in patients aged ≥65 years,[9] and the PROMPT criteria for patients aged 45 to 64 years, at the start of the first NIAD.[10] The Beers criteria includes lists of potentially inappropriate drugs that should be avoided in patients aged ≥65 years as a whole or those with certain medication conditions or at an increased risk of an ADE or DDI.[9] The 2015 version was the latest published version for the years 2016-2019. The PROMPT criteria are used to optimise medication prescribing practices for middle-aged adults (i.e., 45-64 years old).[10] It includes recommendations relevant to this age group considering age-related factors, such as differences in the prevalence of diseases and polypharmacy.

While Beers list encompasses 92 criteria, PROMPT has 22 criteria. For the assessment of PIPs, we included only those criteria that can be fully or partially applied in a primary care database, without information on drugs prescribed during hospital stay or in secondary care. We excluded 17 criteria from Beers (i) because of unavailability of data (e.g., severity of disease, medication prescribed during hospital stay, or dose reduction information in patients with reduced kidney function), or (ii) they were ruled out by study design (i.e., due to exclusion criteria), resulting in 75 indicators from Beers and 22 from PROMPT included in the analysis. Nevertheless, the availability of clinical and prescription information allowed a larger number of Beers and PROMPT criteria to be applied than in previous studies.[14,19] Moreover, a multidisciplinary team collaborated to align the applicability of each criterion using primary care data, aiming to find consensus through the lists and achieve optimal results in a consistently manner. To accommodate differences between the US and UK drug availability, we adapted the Beers criteria by including drugs from the BNF previously published.[20] As PROMPT was tailored to the UK and the Republic of Ireland, no adaptation regarding the drug list was needed.

The list of criteria used, their recommendation, rationale, applicability using primary care data (i.e., fully applied or partially applied), limitation, and an extensive description used in each criterium to assess PIPs according to Beers and PROMPT are listed in the Supplementary Table S1 and Supplementary Table S2, respectively. Additionally, information on ATC codes considered for the identification of the corresponding drugs and Read codes considered for identification of the corresponding diagnosis in each criterion are listed in Supplementary Table S3 for Beers and Supplementary Table S4 for PROMPT criteria. All codes were manually reviewed and confirmed by a medical doctor and clinical pharmacist.

Patients were categorised into those who received a PIP criteria drug, drug combination, or drug-disease combination. Criteria which specified a particular dosage not to exceed, such as prescription of proton pump inhibitors (PPIs) above recommended maintenance dosages for >8 weeks (PROMPT criteria 22), were assessed by calculating the daily dose using the strength, quantity prescribed, and the dose per day or duration for each prescription in the period of assessment.

Whenever dosage information was not available in the data set for drugs of which judgment of appropriate use was dependant on prescribed dose or duration, the cumulative dose (CD) was calculated per drug compound in the database using the defined daily dose (DDD) by the ATC index whenever provided (available at https://www.whocc.no/atc_ddd_index/). This index represents an average maintenance dose per day for a drug used for its main indication in adults, and thus allows for comparisons of drug consumption at an international level. Moreover, the ATC index is freely available worldwide. Whenever the ATC index was not available, we adopted the (minimal) daily dose recommended by the BNF formulary, as specified in the ‘PIPs definition’ in Supplementary Table S1 and Supplementary Table S2. Finally, for multiple prescriptions with only one record having information on dosage/frequency, we assumed the same prescription mode for all prescriptions of the same drug compound for the same patient.

### 2.5 Statistical analysis

We reported patient characteristics at index date, stratified by age groups (40-64 years and 65+) and polypharmacy status (<5 drugs and 5+), as a mean and standard deviation or median and interquartile range (IQR) or counts and proportions, as applicable. We identified alcohol use and smoking based upon the most recent value recorded in the database (i.e., either at or before index date). Body mass index was calculated using the value closest to the index date. However, as the height of an individual is not likely to vary significantly during adulthood, the closest value to the index date (before or after) was considered. We summarised the mean HbA1c and eGFR using the most recently recorded values up to the previous six months prior to the index date, and a history of comorbidities was assessed if ever registered in the database before the index date. We reported all patient characteristics among older and middle-aged adults, stratified by polypharmacy status.

We estimated the number of PIPs per patient at the start of the first NIAD treatment in each of the strata, which was identified as the number of patients with at least one day with any PIPs. Additionally, for the individual PIPs criteria, prevalence was reported in patients receiving at least one day of the drug(s) under evaluation of PIPs at or within 90 days to index date. As a sensitivity analysis, for drugs of which judgment of PIPs was dependent on prescribed dose or duration, the prevalence of PIPs was reported separately for patients having complete information on dose or duration by dividing the patient number by the total number of patients with complete information on dose or duration. Numbers fewer than seven are suppressed according to data use agreement with IMRD. All analyses were performed using the R software version (4.1.2). The study protocol was approved by the IQVIA Scientific Research Committee (study reference number: 22SRC047).

## 3 Results

A total of 28,604 patients initiating a NIAD between 2016 and 2019 (Figure 1) were eligible for inclusion. The demographic characteristics at the index are provided in Table 1, stratified by age and polypharmacy status. Overall, 18,494 (64.7%) patients had polypharmacy at the index date or within 90 days before the index date and were categorised as having polypharmacy. The prevalence of polypharmacy increased with age (older adults = 77.8%; middle-aged adults = 55.2%), and sulfonylureas (SU), dipeptidyl peptidase 4 (DPP-4) inhibitors, and sodium-glucose co-transporter (SGLT-2) inhibitors were prescribed more frequently (alone or in combination with metformin) in patients receiving polypharmacy compared to no polypharmacy regardless of age.

**Figure 1.**
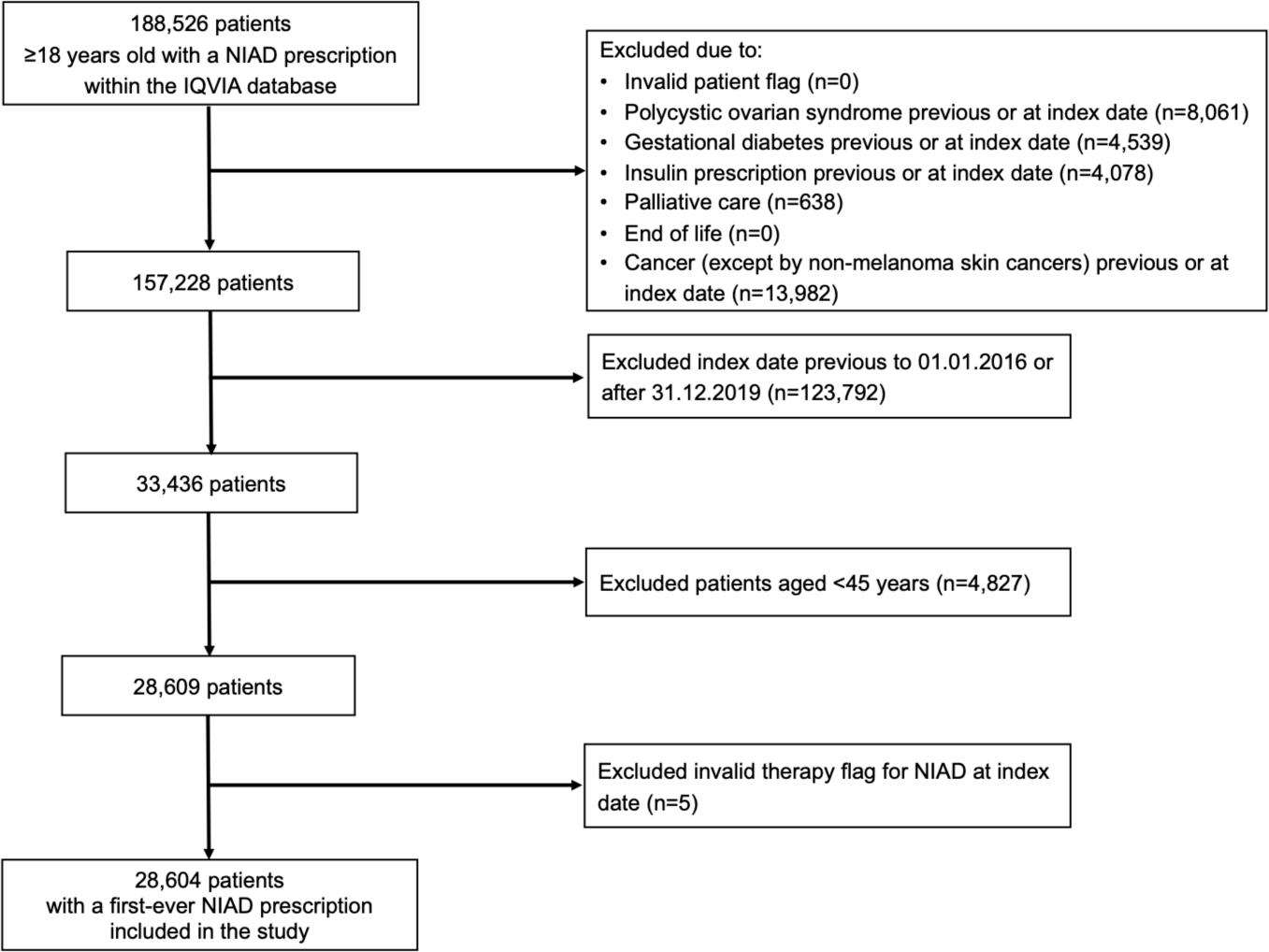
Flow diagram of patient selection. NIAD, non-insulin antidiabetic drug.

**Table 1.** Baseline characteristics of non-insulin antidiabetic drug (NIAD) new-users, stratified by age and polypharmacy status.

| Characteristic | Overall study cohort<br>(N = 28,604) |  | Middle-aged (≥45 and <65 years)<br>(N = 16,654) |  | Older adults (≥65 years)<br>(N = 11,950) |  |
| --- | --- | --- | --- | --- | --- | --- |
|  | no<br>polypharmacy | polypharmacy | no<br>polypharmacy | polypharmacy | No<br>polypharmacy | Polypharmacy |
| **N (%)** | 10,110 (35.3%) | 18,494 (64.7%) | 7,457 (44.8%) | 9,197 (55.2%) | 2,653 (22.2%) | 9,297 (77.8%) |
| **Females** | 3,462 (34.2%) | 8,429 (45.6%) | 2,446 (32.8%) | 4,163 (45.3%) | 1,016 (38.3%) | 4,266 (45.9%) |
| **Mean age (SD)** | 58.8 (9.3) | 64.9 (10.8) | 54.3 (5.4) | 55.9 (5.4) | 71.4 (5.5) | 73.7 (6.6) |
| **Median number of drugs (IQR)** | 3 (2 – 4) | 8 (6 – 11) | 3 (2 – 4) | 8 (6 – 11) | 3 (2 – 4) | 8 (6 – 11) |
| **Mean number of drugs (SD)** | 2.7 (1.1) | 9.1 (4.0) | 2.6 (1.1) | 8.8 (4.0) | 3.0 (1.0) | 9.5 (4.0) |
| **Distribution of drugs n (%)** |  |  |  |  |  |  |
| 1 (NIAD) | 1,653 (5.8%) | - | 1,376 (8.2%) | - | 277 (2.3%) | - |
| 2 - 4 | 8,457 (29.6%) | - | 6,081 (36.5%) | - | 2,376 (19.9%) | - |
| 5 - 9 | - | 11,848 (41.4%) | - | 6,259 (37.6%) | - | 5,589 (46.8%) |
| 10 - 19 | - | 6,180 (21.6%) | - | 2,714 (16.3%) | - | 3,466 (29.0%) |
| ≥ 20 | - | 466 (1.6%) | - | 224 (1.3%) | - | 242 (2.0%) |
| Max n° of drugs | 4 | 41 | 4 | 41 | 4 | 40 |
| **Number of NIADs prescribed at**<br>**index date n (%)** |  |  |  |  |  |  |
| ≥ 2 | 293 (2.9%) | 557 (3.0%) | 230 (3.1%) | 348 (3.8%) | 63 (2.4%) | 209 (2.2%) |
| **NIAD class<sup>1</sup>** |  |  |  |  |  |  |
| Biguanides (Metformin) | 9,611 (96.8%) | 17,082 (94.5%) | 7,231 (97.0%) | 8,890 (96.7%) | 2,551 (96.2%) | 8,592 (92.4%) |
| Sulfonylureas | 615 (6.2%) | 1,319 (7.4%) | 472 (6.3%) | 659 (7.2%) | 157 (5.9%) | 712 (7.7%) |
| DPP-4 inhibitors | 112 (1.1%) | 503 (2.8%) | 61 (0.8%) | 125 (1.4%) | 53 (2.0%) | 393 (4.2%) |
| SGLT-2 inhibitors | 47 (0.5%) | 115 (0.6%) | 45 (0.6%) | 77 (0.8%) | <7 | 40 (0.4%) |
| GLP-1 analogues | 10 (0.1%) | 22 (0.1%) | 12 (0.2%) | 18 (0.2%) | <7 | <7 |
| Thiazolidinediones | 8 (0.08%) | 12 (0.06%) | <7 | <7 | <7 | 7 (0.8%) |
| Others | 7 (0.07%) | 18 (0.1%) | <7 | 14 (0.2%) | <7 | <7 |
| **Smoking status<sup>2</sup>, n (%)** |  |  |  |  |  |  |
| Current | 1,629 (16.1%) | 3,243 (17.5%) | 1,336 (17.9%) | 2,242 (24.4%) | 293 (11.0%) | 1,001 (10.8%) |
| Never | 5,513 (54.5%) | 8,096 (43.8%) | 4,080 (54.7%) | 3,897 (42.4%) | 1,433 (54.0%) | 4,199 (45.2%) |
| Former | 2,910 (28.8%) | 7,079 (38.3%) | 2,003 (26.9%) | 3,012 (32.7%) | 907 (34.2%) | 4,067 (43.7%) |
| Unknown / Missing value | 58 (0.6%) | 76 (0.4%) | 38 (0.5%) | 46 (0.5%) | 20 (0.8%) | 30 (0.3%) |
| **Alcohol use<sup>2</sup>, n (%)** |  |  |  |  |  |  |
| Current | 7,313 (72.3%) | 11,950 (64.6%) | 5,412 (72.6%) | 6,030 (65.6%) | 1,901 (71.7%) | 5,920 (63.7%) |
| Never (Lifelong teetotaler) | 1,826 (18.1%) | 4,590 (24.8%) | 1,332 (17.9%) | 2,146 (23.3%) | 494 (18.6%) | 2,444 (26.3%) |
| Former | 322 (3.2%) | 1,187 (6.4%) | 219 (2.9%) | 591 (6.4%) | 103 (3.9%) | 596 (6.4%) |
| Unknown / Missing value | 649 (6.4%) | 767 (4.1%) | 494 (6.6%) | 430 (4.7%) | 155 (5.8%) | 337 (3.6%) |
| **BMI<sup>3</sup>, n (%)** |  |  |  |  |  |  |
| Obese (BMI ≥30.0 kg/m²) | 5,921 (58.6%) | 12,083 (65.3%) | 4,675 (62.7%) | 6,862 (74.6%) | 1,246 (47.0%) | 5,221 (56.1%) |
| Overweight (BMI 25.0 – 29.9<br>kg/m²) | 2,944 (29.1%) | 4,616 (25.0%) | 1,973 (26.5%) | 1,708 (18.6%) | 971 (36.6%) | 2,908 (31.3%) |
| Normal/ Underweight (BMI < 25.0<br>kg/m²) | 878 (8.7%) | 1,380 (7.5%) | 540 (7.2%) | 404 (4.4%) | 338 (12.7%) | 976 (10.5%) |
| Missing value | 367 (3.6%) | 415 (2.2%) | 269 (3.6%) | 223 (2.4%) | 98 (3.7%) | 192 (2.1%) |
| **History of disease, n (%)** |  |  |  |  |  |  |
| Hypertension | 3,246 (32.1%) | 11,409 (61.7%) | 2,321 (28.5%) | 4,939 (53.7%) | 1,123 (42.3%) | 6,470 (69.6%) |
| Coronary heart disease | 172 (1.7%) | 3,780 (20.4%) | 90 (1.2%) | 1,373 (14.9%) | 82 (3.1%) | 2,407 (25.9%) |
| Heart failure | 10 (0.1%) | 782 (4.2%) | <7 | 234 (2.5%) | <7 | 548 (5.9%) |
| Cerebrovascular disease | 185 (1.8%) | 1,773 (9.6%) | 101 (1.4%) | 561 (6.1%) | 84 (3.2%) | 1,212 (13.0%) |
| Chronic kidney disease | 376 (3.7%) | 2,558 (13.8%) | 156 (2.1%) | 504 (5.5%) | 220 (8.3%) | 2,054 (22.1%) |
| Chronic liver disease | 257 (2.5%) | 823 (4.5%) | 192 (2.6%) | 496 (5.4%) | 65 (2.5%) | 327 (3.5%) |
| COPD | 208 (2.1%) | 2,408 (13.0%) | 114 (1.5%) | 922 (10.0%) | 94 (3.5%) | 1,486 (16.0%) |
| Asthma | 810 (8.0%) | 3,703 (20.0%) | 653 (8.8%) | 2,056 (22.4%) | 57 (5.9%) | 1,617 (17.7%) |
| Sleep disorders | 707 (7.0%) | 2,847 (15.4%) | 566 (7.6%) | 1,604 (17.4%) | 141 (5.3%) | 1,243 (13.4%) |
| Depression | 1,993 (19.7 %) | 6,655 (36.0%) | 1,600 (21.5%) | 4,013 (43.6%) | 393 (14.8%) | 2,642 (28.4%) |
| Alzheimer/Dementia | 39 (0.4%) | 406 (2.2%) | 12 (0.2%) | 32 (0.3%) | 27 (1.0 %) | 374 (4.0 %) |
| Hypothyroidism | 426 (4.2%) | 2,224 (12.0%) | 281 (3.8%) | 978 (10.6%) | 145 (5.5%) | 1,246 (13.4%) |
| Osteoarthritis | 1,169 (11.6%) | 5,003 (27.1%) | 592 (7.9%) | 1,621 (17.6%) | 577 (21.7%) | 3,382 (36.4%) |
| Osteoporosis | 94 (0.9%) | 829 (4.5%) | 32 (0.4%) | 169 (1.8%) | 62 (2.3%) | 660 (7.1%) |
| **Lab values** |  |  |  |  |  |  |
| **Most recent HbA<sub>1c</sub> measurement six months prior to index date n (%)<sup>4</sup>** |  |  |  |  |  |  |
| <6.5% (48 mmol/mol) | 304 (3.0%) | 647 (3.5%) | 235 (3.2%) | 325 (3.5%) | 69 (2.6%) | 322 (3.5%) |
| 6.5 – 7.4% (48 – 57 mmol/mol) | 2,410 (23.8%) | 5,594 (30.2%) | 1,672 (22.4%) | 2,618 (28.5%) | 738 (27.8%) | 2,976 (32.0%) |
| 7.5 – 8.5% (58 – 69 mmol/mol) | 2,010 (19.9%) | 4,614 (24.9%) | 1,389 (18.6%) | 2,087 (22.7%) | 621 (23.4%) | 2,527 (27.2%) |
| >8.5% (69 mmol/mol) | 4,419 (43.7%) | 5,781 (31.3%) | 3,429 (46.0%) | 3,211 (34.9%) | 990 (37.3%) | 2,570 (27.6%) |
| Missing value | 967 (9.6%) | 1,858 (10.0%) | 732 (9.8%) | 956 (10.4%) | 235 (8.9%) | 902 (9.7%) |
| **Most recent eGFR measurement (mL/min/1.73 m<sup>2</sup>) six months prior to index date n (%)<sup>4</sup>** |  |  |  |  |  |  |
| < 30 | 8 (0.1%) | 73 (0.4%) | <7 | 13 (0.1%) | <7 | 60 (0.6%) |
| 30 – 59 | 452 (4.5%) | 2,406 (13.0%) | 184 (2.5%) | 499 (5.4%) | 268 (10.1%) | 1,907 (20.5%) |
| ≥60 | 6,886 (68.1%) | 11,188 (60.5%) | 5,190 (69.6%) | 6,067 (66.0%) | 1,696 (63.9%) | 5,121 (55.1%) |
| Missing value | 2,764 (27.3%) | 4,827 (27.6%) | 2,083 (27.9%) | 2,618 (28.5%) | 683 (25.7%) | 2,209 (23.8%) |

<sup>1</sup> The summed proportion value is above 100%, as a patient may receive more than one NIAD compound at the index date.
<sup>2</sup> Identified values closest to the index date (i.e., at or before index date).
<sup>3</sup> BMI was assessed using the most recently recorded value for weight (at or at any time before index date) and height (at or at any time before or after index date).
<sup>4</sup> The most recent value (at index date and previous six-months) registered in the database with a valid unit was considered.
**Abbreviations:** NIAD: non-insulin antidiabetic drug; SD: standard deviation; IQR: interquartile range; DPP-4: Dipeptidyl peptidase 4; SGLT-2: Sodium-glucose co-transporter 2; GLP-1: Glucagon-like peptide-1; BMI: body mass index; COPD: chronic obstructive pulmonary disease; HbA<sub>1c</sub>: hemoglobin A<sub>1c</sub>; eGFR: Estimated glomerular filtration rate;
**Note:** A history of comorbidities was assessed if ever registered in the database previous to or at index date. Numbers fewer than seven are suppressed according to data use agreement with IMRD.

Among older patients without polypharmacy, 6.5% received ≥1 PIP with a maximum of four PIPs per patient, while 39.6% of those with polypharmacy had ≥1 PIP with a maximum of 10 PIPs per patient (Table 2). Similarly, the prevalence of PIPs was higher among middle-aged adults with polypharmacy than in those without. Among middle-aged adults without polypharmacy, few PIPs were identified, with 98.5% of patients having no PIPs, while 22.7% of those with polypharmacy had ≥1 PIP with a maximum of four PIPs per patient (Table 2).

**Table 2.**
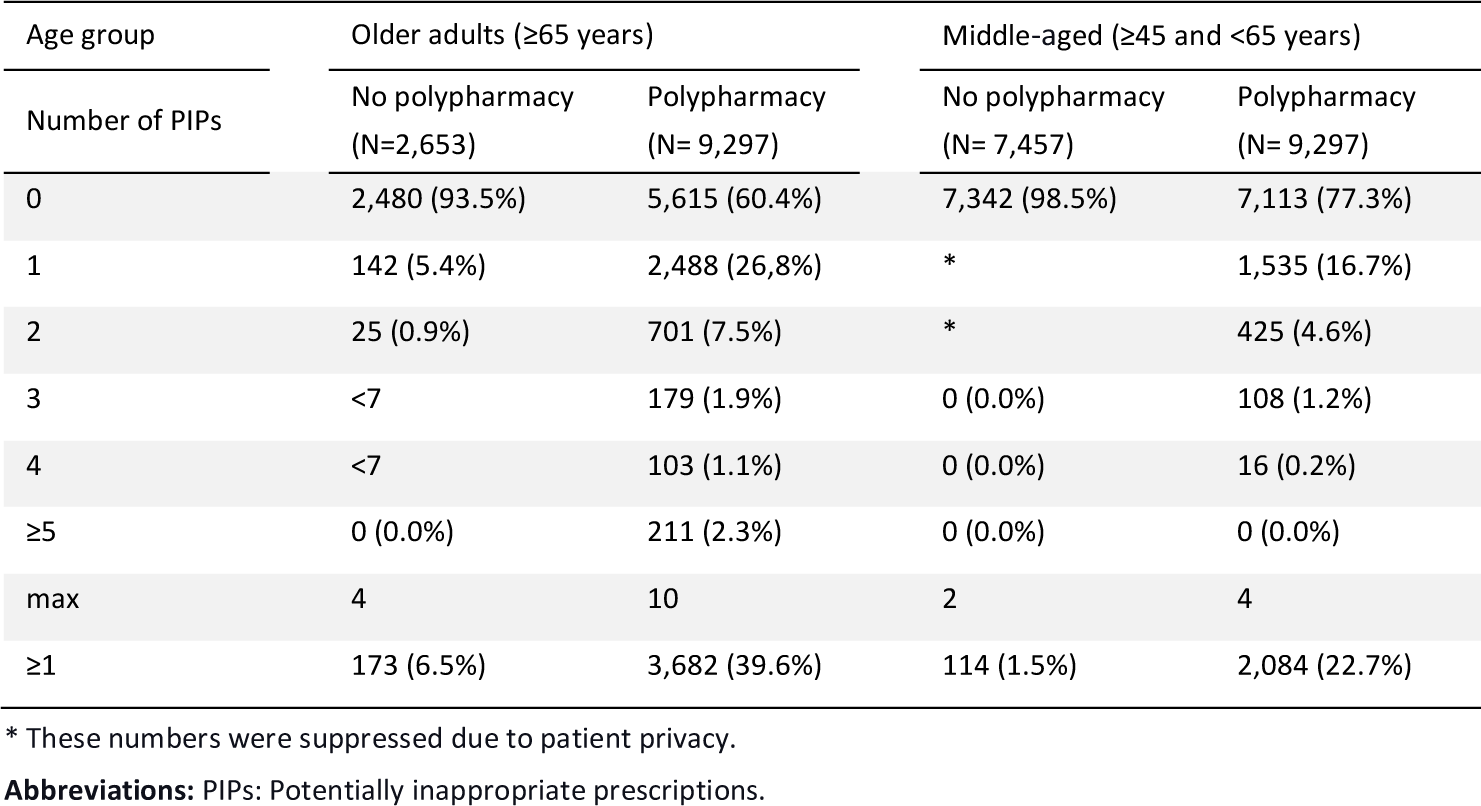
Prevalence of potentially inappropriate prescriptions (PIPs) in middle-aged and older adults according to the American Geriatrics Society (AGS) Beers criteria 2015 and the Prescribing Optimally in Middle-aged People’s Treatments (PROMPT) criteria, respectively, stratified by polypharmacy status.

Table 3 provides the prevalence of PIPs among older adults according to the Beers criteria. The most frequently occurring PIP was the use of PPIs for >8 weeks unless for high-risk patients (criteria 33 [n = 1,034, 11.1%]), followed by the use of high anticholinergic antidepressants monotherapy or in combination to other drugs for management of depression (criteria 16 [n=951, 10.2%]), the use of antipsychotics, benzodiazepines (BZD), non-BZD, BDZ receptor agonist hypnotics, tricyclic antidepressants (TCA) and selective serotonin reuptake inhibitors (SSRI) in patients with a history of falls or fractures (criteria 3 [n=911, 9.8%]), and the use of peripheral alpha-1 blockers in patients with hypertension (criteria 2 [n=495, 5.3%]). Among patients having no polypharmacy, the most frequent PIPs were PPI use >8 weeks unless for high-risk patients, erosive esophagitis, Barrett’s esophagitis, pathological hypersecretory condition, or demonstrated need for maintenance treatment (criteria 33 [n=46, 1.7%]) and the use of antidepressants monotherapy or in combination to other drugs for management of depression (criteria 16 [n=31, 1.2%]). The sensitivity analysis identifying the prevalence of individual PIPs among patients having complete information on dose or duration is shown in Table 3.

**Table 3.**
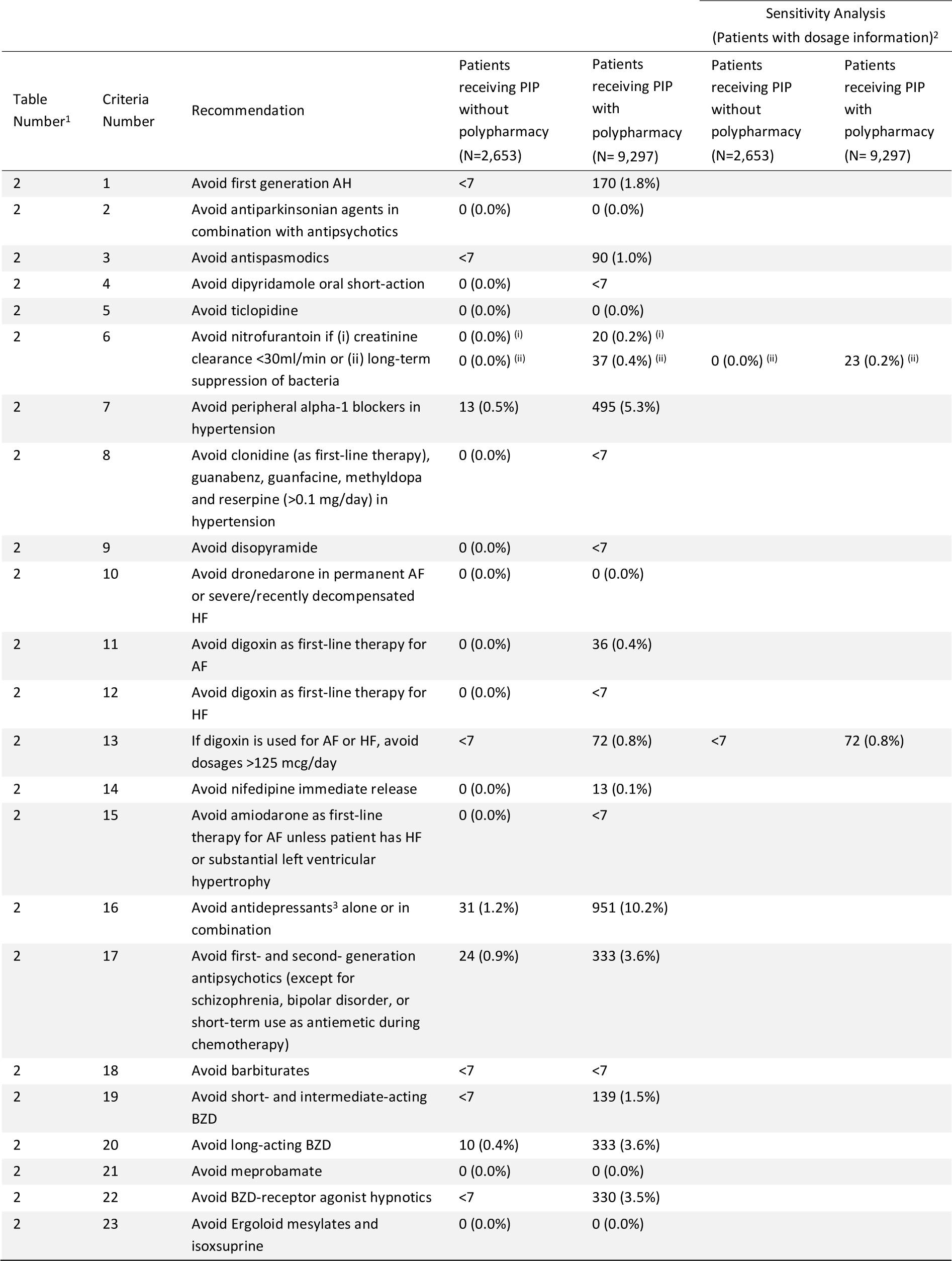
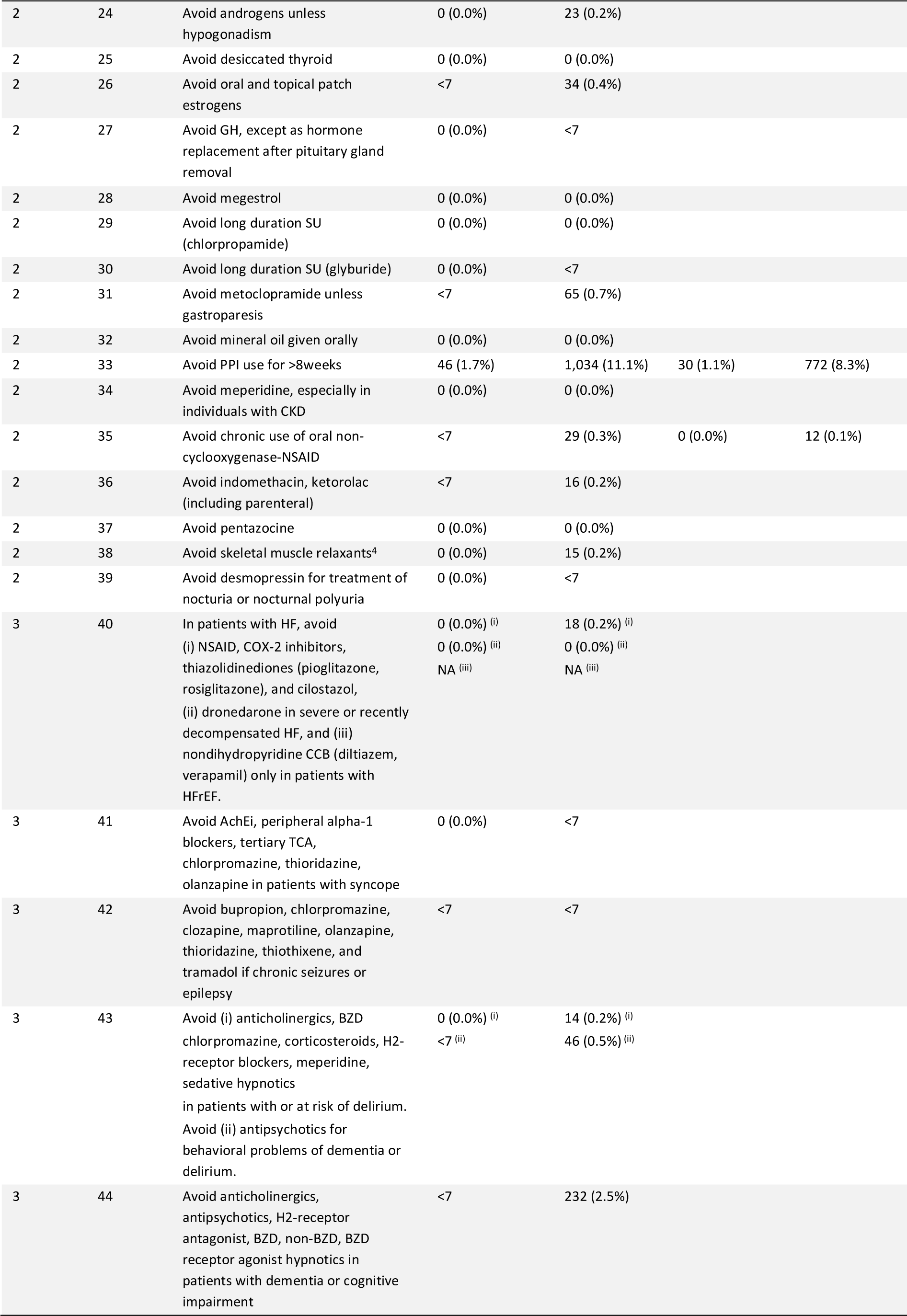
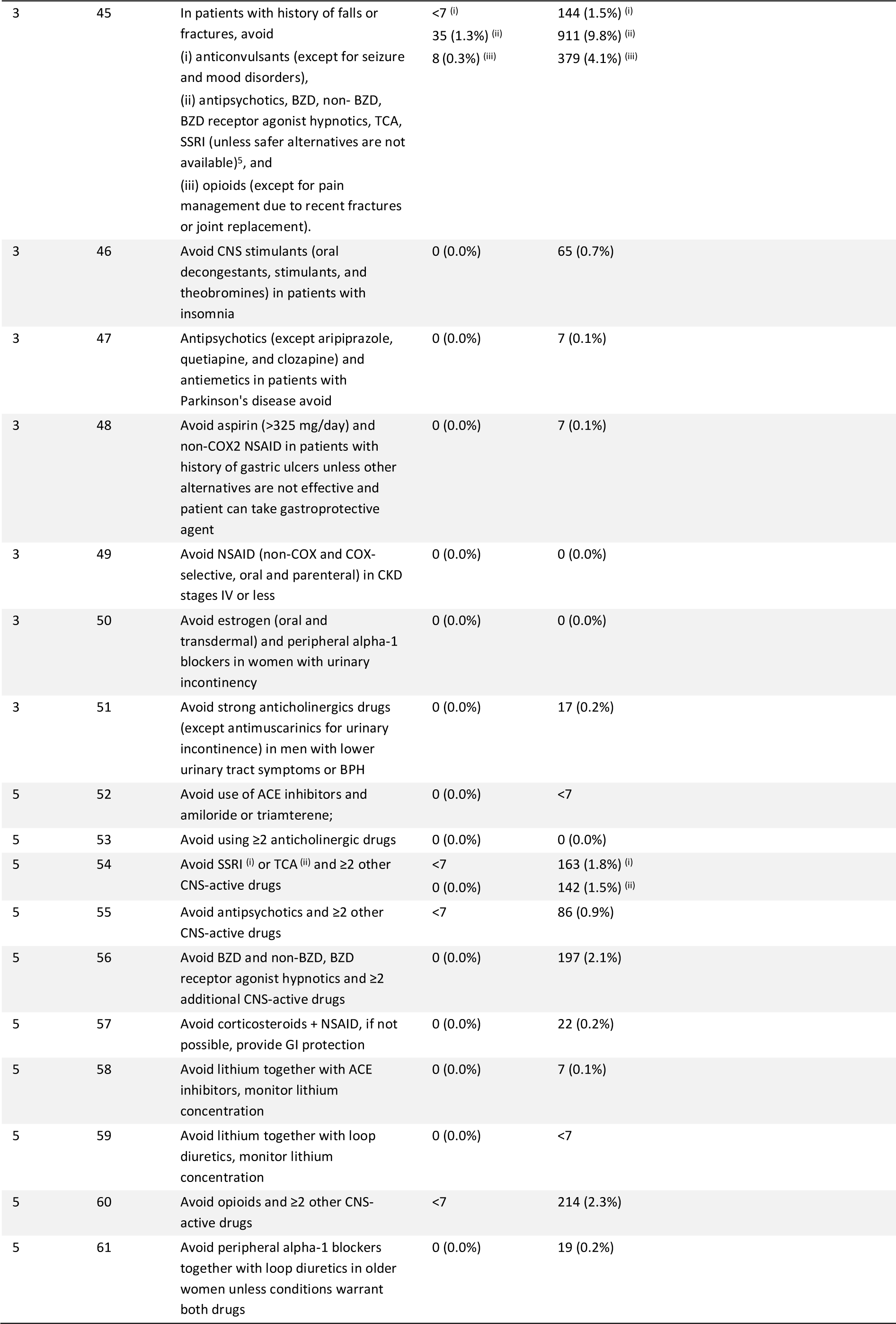
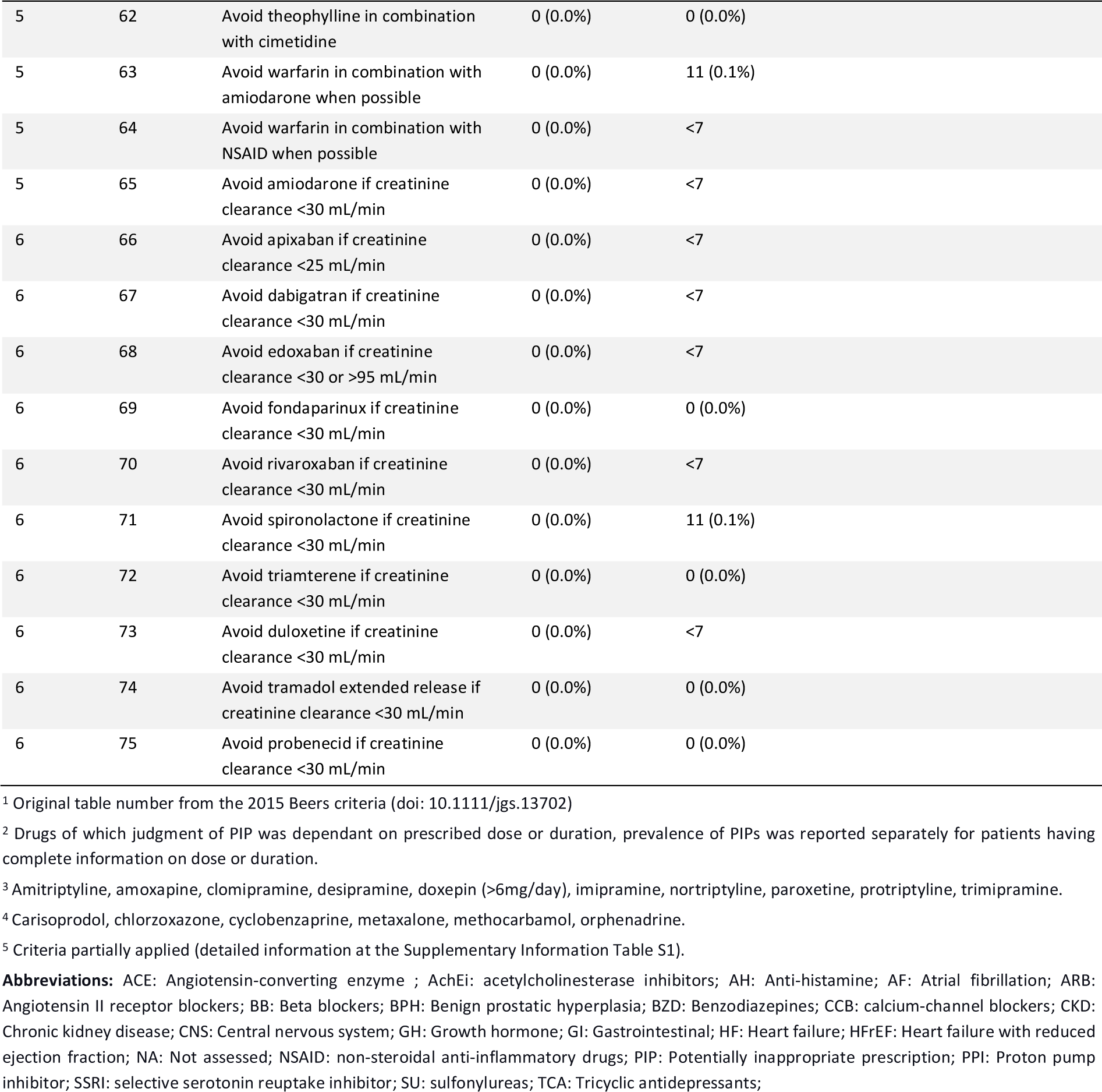
Prevalence of individual potentially inappropriate prescriptions (PIPs) in older adults with new-onset type 2 diabetes (T2DM) according to the American Geriatrics Society (AGS) Beers criteria 2015, stratified by polypharmacy status.

The prevalence of individual PIPs in middle-aged adults at the start of the first NIAD treatment is provided in Table 4. The highest prevalence in middle-aged patients with polypharmacy was observed for strong opioids without co-prescription of laxatives (criteria 17 [n=381, 4.1%]); followed by the combination of non-steroidal anti-inflammatory drugs (NSAIDs) and low-dose aspirin (acetylsalicylic acid) or SSRI without adequate gastrointestinal protection (criteria 21 [n=361, 3.9%]), long-term use of BZD (criteria 14 [n=293, 3.2%]), and PPI use above maintenance dosages for >8 weeks (criteria 2 [n=269,2.9%]). In contrast, in patients having no polypharmacy, the highest prevalence was observed for first-generation anti-histamines as first-line treatment >7 days (criteria 8 [n= 14, 0.2%]) and for PPI above maintenance dose for >8 weeks (criteria 2 [n=13, 0.2%]). The sensitivity analysis identifying the prevalence of individual PIPs among patients having complete information on dose or duration is shown in Table 4.

**Table 4.**
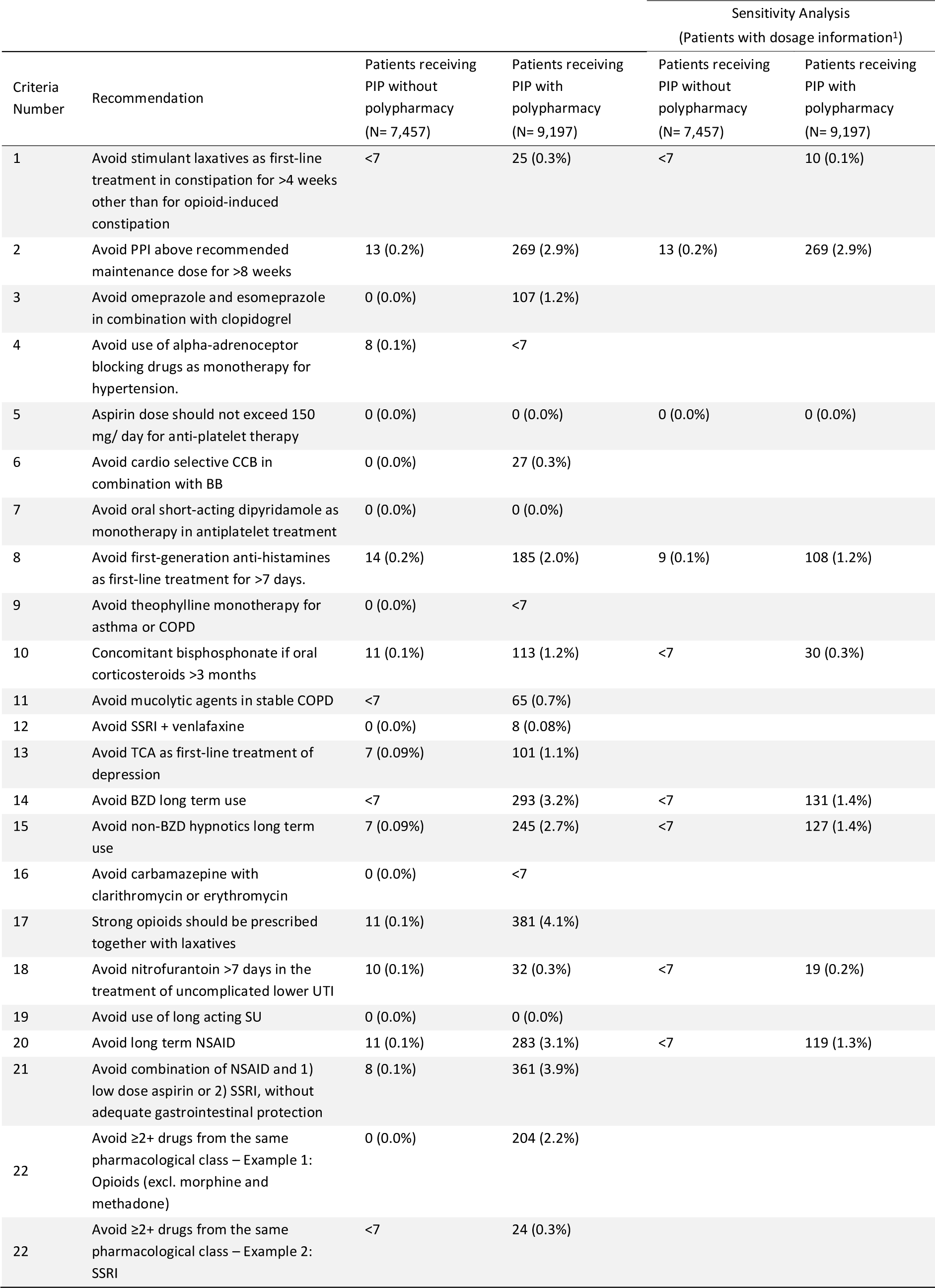
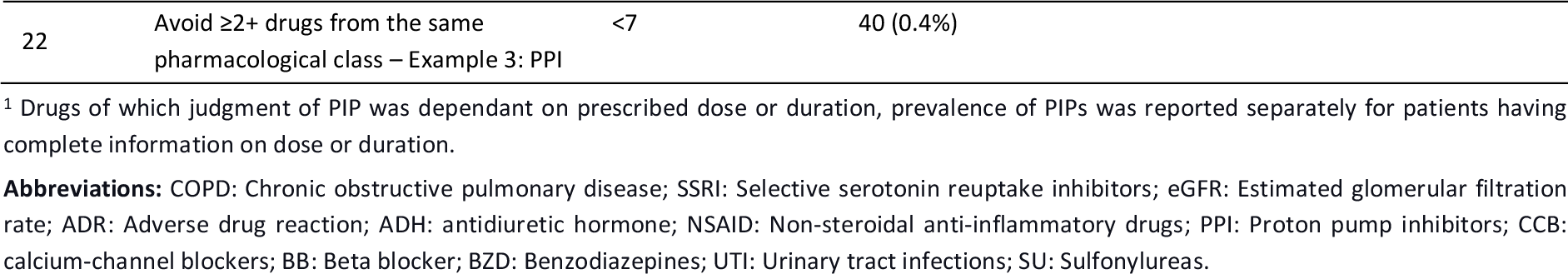
Prevalence of individual potentially inappropriate prescriptions (PIPs) in middle-aged adults with new-onset type 2 diabetes (T2DM) according to the Prescribing Optimally in Middle-aged People’s Treatments (PROMPT) criteria, stratified by polypharmacy status.

| Criteria Number | Recommendation | Patients receiving PIP without polypharmacy (N= 7,457) | Patients receiving PIP with polypharmacy (N= 9,197) | Sensitivity Analysis<br>(Patients with dosage information<sup>1</sup>) |  |
| --- | --- | --- | --- | --- | --- |
|  |  |  |  | Patients receiving PIP without polypharmacy (N= 7,457) | Patients receiving PIP with polypharmacy (N= 9,197) |
| 1 | Avoid stimulant laxatives as first-line treatment in constipation for >4 weeks other than for opioid-induced constipation | <7 | 25 (0.3%) | <7 | 10 (0.1%) |
| 2 | Avoid PPI above recommended maintenance dose for >8 weeks | 13 (0.2%) | 269 (2.9%) | 13 (0.2%) | 269 (2.9%) |
| 3 | Avoid omeprazole and esomeprazole in combination with clopidogrel | 0 (0.0%) | 107 (1.2%) |  |  |
| 4 | Avoid use of alpha-adrenoceptor blocking drugs as monotherapy for hypertension. | 8 (0.1%) | <7 |  |  |
| 5 | Aspirin dose should not exceed 150 mg/ day for anti-platelet therapy | 0 (0.0%) | 0 (0.0%) | 0 (0.0%) | 0 (0.0%) |
| 6 | Avoid cardio selective CCB in combination with BB | 0 (0.0%) | 27 (0.3%) |  |  |
| 7 | Avoid oral short-acting dipyridamole as monotherapy in antiplatelet treatment | 0 (0.0%) | 0 (0.0%) |  |  |
| 8 | Avoid first-generation anti-histamines as first-line treatment for >7 days. | 14 (0.2%) | 185 (2.0%) | 9 (0.1%) | 108 (1.2%) |
| 9 | Avoid theophylline monotherapy for asthma or COPD | 0 (0.0%) | <7 |  |  |
| 10 | Concomitant bisphosphonate if oral corticosteroids >3 months | 11 (0.1%) | 113 (1.2%) | <7 | 30 (0.3%) |
| 11 | Avoid mucolytic agents in stable COPD | <7 | 65 (0.7%) |  |  |
| 12 | Avoid SSRI + venlafaxine | 0 (0.0%) | 8 (0.08%) |  |  |
| 13 | Avoid TCA as first-line treatment of depression | 7 (0.09%) | 101 (1.1%) |  |  |
| 14 | Avoid BZD long term use | <7 | 293 (3.2%) | <7 | 131 (1.4%) |
| 15 | Avoid non-BZD hypnotics long term use | 7 (0.09%) | 245 (2.7%) | <7 | 127 (1.4%) |
| 16 | Avoid carbamazepine with clarithromycin or erythromycin | 0 (0.0%) | <7 |  |  |
| 17 | Strong opioids should be prescribed together with laxatives | 11 (0.1%) | 381 (4.1%) |  |  |
| 18 | Avoid nitrofurantoin >7 days in the treatment of uncomplicated lower UTI | 10 (0.1%) | 32 (0.3%) | <7 | 19 (0.2%) |
| 19 | Avoid use of long acting SU | 0 (0.0%) | 0 (0.0%) |  |  |
| 20 | Avoid long term NSAID | 11 (0.1%) | 283 (3.1%) | <7 | 119 (1.3%) |
| 21 | Avoid combination of NSAID and 1) low dose aspirin or 2) SSRI, without adequate gastrointestinal protection | 8 (0.1%) | 361 (3.9%) |  |  |
| 22 | Avoid ≥2+ drugs from the same pharmacological class – Example 1: Opioids (excl. morphine and methadone) | 0 (0.0%) | 204 (2.2%) |  |  |
| 22 | Avoid ≥2+ drugs from the same pharmacological class – Example 2: SSRI | <7 | 24 (0.3%) |  |  |

**Table 4.** Continued.
|  |  |  |  |
| --- | --- | --- | --- |
| 22 | Avoid ≥2+ drugs from the same<br>pharmacological class – Example 3: PPI | <7 | 40 (0.4%) |

<sup>1</sup> Drugs of which judgment of PIP was dependant on prescribed dose or duration, prevalence of PIPs was reported separately for patients having complete information on dose or duration.
**Abbreviations:** COPD: Chronic obstructive pulmonary disease; SSRI: Selective serotonin reuptake inhibitors; eGFR: Estimated glomerular filtration rate; ADR: Adverse drug reaction; ADH: antidiuretic hormone; NSAID: Non-steroidal anti-inflammatory drugs; PPI: Proton pump inhibitors; CCB: calcium-channel blockers; BB: Beta blocker; BZD: Benzodiazepines; UTI: Urinary tract infections; SU: Sulfonylureas.

## 4 Discussion

This is the first study to apply the Beers and the PROMPT and list to a large population-based primary care database to evaluate the prevalence of PIPs among both older and middle-aged adults initiating therapy for T2DM with and without polypharmacy. The prevalence of PIPs was overall higher in patients receiving polypharmacy compared to no polypharmacy. While the prevalence of PIPs was higher among older patients with polypharmacy (39.6%), we found that 22.7% of middle-aged patients with polypharmacy received ≥1 PIPs. At the individual PIP level, long-term use of PPIs and strong opioids without laxatives were the most frequently identified PIPs among older patients (criteria 33) and middle-aged patients (criteria 17), respectively, with polypharmacy (11.1% and 4.1%, respectively). The results of this study highlight the frequency of PIPs among patients with T2DM and the subsequent importance of medication reviews to avoid harmful adverse events due to PIPs.

While this is the first study to apply the Beers and PROMPT criteria to patients initiating their first NIAD treatment, our results align with the existing literature.[13,14,16,21–24] The only study to date, that has applied both Beers and PROMPT in patients with T2DM receiving polypharmacy to assess PIPs, observed higher PIPs prevalence in middle-aged patients (36.9% to 39.0%) than in older patients (24.9% to 28.0%).[14] Limitations included the absence of clinical information, and, therefore, fewer criteria were assessed. The latter is particularly important for the criteria related to medication with multiple indications. While our study was also limited by missing information, we could address more criteria due to the availability of clinical parameters and synchronization of data and criteria list through a multidisciplinary team, including a clinical pharmacologist and a clinical pharmacist. For example, in PROMPT criteria number 13, the lack of information on depression diagnosis implicates the overestimation of PIPs, particularly because TCA is used within different indications. While we acknowledge the diagnosis codes for depression are subject to misclassification and are an underestimation of true prevalence, we were able to combine our prescribing data with previous diagnosis and clinical patient records to overcome this limitation.

Another study using Beers criteria to assess PIPs in elderly with T2DM and polypharmacy identified that the prevalence of PIPs was 54.5%.[13] While this is substantially higher than the 39.6% found in our study, we expect the difference is largely due to differences in the study population. The study by Formiga et al. included patients age 75+ who were admitted to the hospital, while we included all older adults initiating a NIAD in general practice with a mean age of 73.7 years.[13] Thus, we expect that our results more likely reflect the prevalence of PIPs in the general population of older adults with diabetes.

As the number of studies assessing the Beers and PROMPT criteria in patients with T2DM is limited, we also note that our results are comparable to the reported prevalence of PIPs in the general population. Studies applying Beers and PROMPT showed PIPs prevalence ranging from 34.2% to 64.8% in older adults,[16,19,23,24] and 21.1% to 46.2% in middle-aged adults.[19,21,22] Nonetheless, it is challenging to draw an accurate one-to-one comparison with most studies, as very often, only part of the criteria was applied due to limitations on the dataset. Among the studies applying the Beers criteria, most observational studies implemented a subset of the criteria list.[25] Nevertheless, one study that implemented the entire list of criteria used administrative claims data from the USA and included patients 65+, with no stratification between polypharmacy status. The study found the prevalence of PIPs ranged between 34.2% to 37.6%. As the study did not stratify by polypharmacy status, it is difficult to directly compare our results. Overall, 3,682 patients (173 without polypharmacy and 3,682 with polypharmacy) had at least one PIP, which corresponds to a PIP prevalence of 32.3% among our older adults. While this is marginally lower, than the study by Jirón et al., the marginal disparities between these estimates may be due to differences between US and UK clinical settings and data sources used in each study.

At the individual PIP level in older adults, the long-term use of PPIs is of particular concern as it may increase the risk of adverse drug reactions, such as chronic kidney disease, particularly in high doses,[26] and B12 deficiency.[27] Studies in primary care and emergency settings indicate that PPIs are frequently prescribed for inappropriate indications or for indications where their use provides little to no benefit.[28] Moreover, patients may stay long-term on PPIs, often indefinitely, even without a clear clinical indication.[29,30] Therefore, our findings indicate the need for carefully reviewing indications of PPIs to minimise the risk of long-term PPI-related adverse outcomes in older patients with diabetes, particularly in those receiving polypharmacy.

The use of high anticholinergic antidepressants (criteria 16 [10.2%]), and SSRIs, BZD, non-BZD hypnotics, and TCA (criteria 45 [9.8%]) in patients with a history of falls and fractures were among the most frequent PIPs in older adults found this study as well as in previous work.[13,14] Anticholinergic antidepressants may cause ADR such as dry mouth, blurred vision, constipation, urinary dysfunction, sedation, orthostatic hypotension, and cognitive impairment, which all reduce the patient’s quality of life, may lead to increased risk of falls, substantial morbidity, and even mortality.[31] Older adults have a higher sensitivity to anticholinergic effects, with might worsen pre-existing symptoms.[32,33] Patients with dementia and depression may become more confused due to the anticholinergic side effects of some antidepressants as well as patients with orthostatic hypotension may collapse and fall.

These concerns become particularly important in patients with diabetes complications, such as neuropathy, retinopathy, and diabetic cardiomyopathy.[34,35] Although anticholinergic antidepressants are not the first-line pharmacological therapy for treating depression according to the clinical guidelines in the UK,[36] we found high prevalence of drug prescription in older patients. While further research on the appropriateness is needed, our results indicate that clinicians do not withhold from prescribing highly anticholinergic antidepressants to older patients with diabetes and polypharmacy, despite their potential to cause ADR and the availability of safer alternatives.

The use of SSRIs, BZD, non-BZD, and TCA is associated with an increased risk of falls and fractures,[37–39] a primary concern for elderly adults with T2DM. These drugs are often prescribed to the elderly for prolonged periods, without the repeat prescription being reviewed.[39,40] BZD have long been recognised as a contributory factor for falls. Non-BZD hypnotics are considered safer than BZD, however, they have an increased risk for falls when used in long-term.[38] Notably, in the UK, TCA drugs such as amitriptyline, prescribed in low doses for longer term, are considered appropriate to treat neuropathic pain.[41] Thus, although indications cannot be determined from our dataset, the high prevalence of TCA prescription in patients with T2DM may be, at least partially, for its use in pain management. Additionally, despite warnings on the clinical guideline for treating depression in the UK regarding the use of antidepressants in older people with increased risk of falls and fractures,[36] our study indicates that clinicians do not refrain from prescribing these drugs to older patients with a history of falls and fractures which are risk factors for new events. This is of particular concern in older patients with T2DM: besides the risk of falls and fractures increase with age,[42] complications of diabetes (e.g., neuropathy and vision loss) and NIAD treatment adverse effects (e.g., hypoglycaemia in patients receiving SU), can further increase this risk.[43]

At the individual PIP level in middle-aged adults, this study indicates that opioid-related constipation may not be adequately prevented by clinicians, despite being a significant concern and burden to patients.[44] Nonetheless, a possible reason for not adding a laxative drug to opioid treatment could be that patients already benefit from laxatives as over-the-counter (OTC) drugs, and thus, we may have missed those patients in the primary care dataset.

The combination of NSAIDs and SSRIs or low-dose aspirin without gastroprotection (3.9%) was also identified to be broadly used in middle-aged adults receiving polypharmacy. While drug combinations with NSAIDs, such as SSRIs and aspirin,[45–47] are risk factors for upper gastrointestinal bleeding (UGIB), the risk of outcomes associated with these two combinations is attenuated by concomitant use of a PPI.[47] Thus, this study indicates the need for clinical improvement and may help clinicians in tailoring therapy to minimise the risk of UGIB in middle-aged adults with T2DM starting their antidiabetic treatment.

Many of the most frequent PIP criteria in this study relate to the inappropriate duration of medication use. Although short-term use of medications may confide a different risk-benefit ratio than long-term use and may not be considered inappropriate in specific conditions, the high prevalence of the most frequent PIPs found in our study is not compatible with only appropriate drug prescriptions. To approach that, documenting and communicating the intended treatment duration or planned review dates in the medical records can provide valuable information to clinicians, facilitating regular review and discontinuation of inappropriate prescriptions.[48] Moreover, open communication with patients about prescription duration can also help manage expectations and reduce resistance to changing medication regimens.

While GPs may be less likely to discontinue drug prescriptions from secondary care, they play a critical role in conciliating prescriptions from different settings as well as evaluating the benefit of continuing drugs based on a risk-benefit assessment and their patient needs. The lack of evidence and the challenges in assessing treatment benefits and harms were previously identified as barriers to appropriate prescribing, particularly in older patients with multiple comorbidities.[48] To address that, evidence-based guidelines have been developed to support deprescribing of specific drugs, such as PPIs and BZD, which were among the most prevalent PIPs in this study.[49,50] These guidelines may offer valuable guidance to optimize medication regimens, enhance patient safety, and improve outcomes.

A key strength of this study is the detailed insights into the prevalence of PIPs in patients with new-onset T2DM, applying the 2015 AGS Beers criteria adapted to the UK to 65-year-olds and older and the PROMPT criteria to 45- to 64-year-old patients. The availability of primary care clinical data allowed for a more thorough assessment of PIPs specifying conditions in patients with diabetes compared to previous studies.[13,14] The IMRD allowed studying a large number of patients starting a NIAD treatment who are representative of the UK population. In the UK, GPs are central players managing chronic diseases, coordinating drug prescriptions that may be initially prescribed in secondary care, and thus have a favourable position to assess their patients’ medications across diseases. The sensitivity analysis confirmed the robustness of the results, where assumptions were necessary. Another strength is the availability of a clear and detailed definition of each criterion used in this study, which can be applied in future studies to assess PIPs using large population-based databases, facilitating interpretation and comparability between studies.

Our study also has some limitations. First, the Beers criteria is an extensively used and validated explicit process measure developed in the US with the intention of providing a valuable tool for assessing the quality of prescribing in older adults, regardless of their place of residence and their level of frailty. Thus, to accommodate the differences between the US and UK drug availability, we adapted the Beers criteria by including drugs from the British National Formulary, as previously published by Ble et al. We note that while several drugs in the Beers criteria are listed as PIPs, they are not inappropriate in older adults according to the National Institute for Health Care and Excellence (NICE) clinical guidelines in the UK (e.g., amitriptyline, clomipramine, desipramine, and nortriptyline are indicated for management of depression,[36] and nitrofurantoin long-term low dose prophylaxis of recurrent urinary tract infection[52]), whereas other non-recommended drugs (e.g., theophylline as monotherapy for asthma and COPD) are omitted. Similarly, the Beers criteria do not consider varying indications for certain drugs (e.g. amitriptyline for the management of neuropathic pain in the UK).[53] Thus, while Beers identifies those as PIPs, we acknowledge that this may overestimate inappropriate use in the UK. On the other hand, our estimation of PIP prevalence using Beers is likely to be more conservative, as Beers seems to underestimate PIP compared to explicit methods tailored to the context of Europe, such as Screening Tool of Older Persons’ Prescriptions (STOPP) and Screening Tool to Alert to Right Treatment (START) criteria.[13,54] Moreover, although Beers was not tailored to the context of Europe prescribing, clinically relevant issues associated with the inappropriate use of drugs identified using Beers remain relevant in Europe. Finally, Beers was readily adapted to our dataset compared to the STOPP/START criteria.

In addition to the above, we expect that we have likely underestimated medication use, and subsequently PIPs due to limitations with the IMRD. The IMRD only captures drug prescriptions made by GPs. Thus, we were not able to identify OTC drugs, such as NSAIDs and laxatives. Additionally, drugs given during hospitalisation and specialist prescriptions are not captured within the database.

Another limitation is that the Beers list was not created for use in observational data, and therefore, not all of the criteria could be applied due to limitations in our observational data. Failure to apply the full criteria list may have resulted in lower estimates of the overall PIPs prevalence compared to other studies, as well as overestimation of individual PIPs whenever it was partially applied. A complete overview of the limitations associated with each specific criterion are available in Supplementary Table S1 and Table S2. Similarly, due to missing data on some clinical parameters, not all patients had complete information. To overcome this limitation, we conducted a sensitivity analysis among only those patients with complete information, and results remained largely similar.

Finally, while the DDD by the ATC index does not necessarily correspond to the recommended daily dose, we had no access to the indication of drugs due to limitations on our dataset, and thus the exclusive use of BNF formulary as a source of standard daily dose to estimate CD was not feasible. Nevertheless, as the ATC index is freely available worldwide and allows for comparisons of drug consumption at an international level, the use of the DDD ATC index represents a valuable alternative to estimate CD.

## 5 Conclusion

This analysis using primary care data revealed opportunities for optimizing pharmacotherapy in patients newly treated with oral antidiabetics. While the burden of polypharmacy, and thus PIPs, increases with age, this problem is not exclusive to the elderly. Among patients with polypharmacy, 39.6% of older adults had at ≥1 PIP, compared to 22.7% of middle-aged adults with ≥1 PIP. Antidiabetic therapy carries its own challenges and risks, emphasizing the importance of minimizing risks associated with existing medications. Given the challenges of avoiding polypharmacy in adults aged ≥45 years with diabetes, it is crucial to implement measures that review indications, address drug interactions, and consider comorbidities, reducing the likelihood of inappropriate prescribing. Initiating treatment with NIADs in patients with polypharmacy should trigger a comprehensive medication review to optimize prescribing decisions.

## Data Availability

The data that support the findings of this study are available from IQVIA Medical Research Data (IMRD) incorporating THIN, a Cegedim database of anonymised electronic health records in the UK. Restrictions apply to the availability of these data, which were used under license for this study. For further information on how to access the data, contact IQVIA at.

## Acknowledgement

The authors thank Dr Adrian Martinez de la Torre and all members of the Swiss Data Science Center (SDSC) for their technical support and Prof Dr Theresa Shireman for the stimulating discussions on this topic.

## Authorship

AMB: Conceptualisation; Resources; Supervision; Interpretation of results. MLF: Conceptualisation; Methodology; Formal analysis; Investigation; Interpretation of results; Writing original draft; GF: Literature review and implementation of the study. DS: Methodology; Contributed to the design and implementation of the study; Interpretation of results. SW: Methodology; Contributed to the design and implementation of the study; Interpretation of results. All authors reviewed and contributed critically to the manuscript.

