## Supplementary Information for "Examining inappropriate medication in UK primary care for type 2 diabetes patients with polypharmacy"

#### Selection of drugs included in the polypharmacy analysis

**Table S1.** List of 2015 American Geriatrics Society Beers criteria used for the assessment of potential inappropriate prescription (PIP) in older adults.

**Table S2.** List of Prescribing Optimally in Middle-aged People's Treatments (PROMPT) criteria used for the assessment of potential inappropriate prescription (PIP) in middle-aged adults.

**Table S3.** List of Anatomical therapeutic chemical (ATC) classification codes and Read codes used to assess potential inappropriate prescription (PIP) according to the 2015 American Geriatrics Society Beers criteria.

**Table S4.** List of Anatomical therapeutic chemical (ATC) classification codes and Read codes used to assess potential inappropriate prescription (PIP) according to the Prescribing Optimally in Middle-aged People's Treatments (PROMPT) criteria.

### Supplementary Information

#### Selection of drugs included in the polypharmacy analysis:

The database contained 2,256 ATC codes. Of these, vaccines, insulin products, surgical dressings, other non-therapeutic products (e.g., cosmetics and disinfectants), general nutrients as well as records with invalid ATC codes were excluded from the calculation of polypharmacy. NIADs containing two active ingredients in the formulation were split and counted as exposure to two drug compounds (e.g., sitagliptin and metformin [A10BD07] was split in metformin [A10BA02] and sitagliptin [A10BH01]). We restricted our analysis to include drugs with a prescription frequency of 5+ in the database (624 unique ATC codes) during the study period.

**Table S1.** List of 2015 American Geriatrics Society Beers criteria used for the assessment of potential inappropriate prescription (PIP) in older adults.

| Table Number[1] | Criteria Number | Recommendation | Rationale | PIP definition | Limitations | Applicability |
|---|---|---|---|---|---|---|
| **2** | 1 | Avoid first-generation AH<br>Brompheniramine<br>Carbinoxamine<br>Chlorpheniramine<br>Clemastine<br>Cyproheptadine<br>Dexbrompheniramine<br>Dexchlorpheniramine<br>Dimenhydrinate<br>Diphenhydramine (except if oral for severe allergic reactions)<br>Doxylamine<br>Hydroxyzine<br>Meclizine<br>Promethazine<br>Triprolidine | Highly anticholinergic; clearance reduced with advanced age, confusion, dry mouth, constipation, and other anticholinergic effects or toxicity<br>Use of diphenhydramine in situations such as acute treatment of severe allergic reaction may be appropriate | PIP was defined as ≥1 prescription of first-generation AH at the index date or previous 90 days (except if diphenhydramine with a diagnosis of severe allergic reactions in the 14-days previous to drug prescription). | | Fully applied |
| **2** | 2 | Avoid antiparkinsonian agents (benztropine oral and trihexyphenidyl) in combination with antipsychotics | Not recommended for extrapyramidal symptoms with antipsychotics | PIP was defined as ≥1 prescription of antiparkinsonian agent at the index date or previous 90 days and ≥1 prescription of antipsychotic within the 90-days period. To accommodate differences between the US and UK drug availability, we adapted the Beers criteria by including orphenadrine and methocarbamol.[5] | Potential underestimation of PIP as some antipsychotics in the UK (e.g., clozapine) are only prescribed in secondary care and, therefore, were not included in the assessment of PIP. Potential overestimation of PIP, as we assumed that patients having a prescription for antiparkinsonian agent and antipsychotic medication within the 90 days interval were used concomitantly. | Fully applied |
| **2** | 3 | Avoid antispasmodics<br>Atropine<br>Belladona alkaloids<br>Clinidium-Chlordiazepoxide<br>Dicyclomine<br>Hyoscyamine<br>Propantheline<br>Scopolamine | Highly anticholinergic, uncertain effectiveness | PIP was defined as ≥1 prescription of antispasmodic agents at the index date or previous 90 days. To accommodate differences between the US and UK drug availability, we adapted the Beers criteria by including dicycloverine and hyoscine.[5] | | Fully applied |

**Table S1.** Continued.

| | | | | | | |
|---|---|---|---|---|---|---|
| **2** | 4 | Avoid dipyridamole oral short-action (does not apply to extended release combination with aspirin) | May cause orthostatic hypotension; IV formulation is acceptable for use in cardiac stress testing | PIP was defined as ≥1 prescription of dipyridamole (oral short-action formulation) at the index date or previous 90 days. | Potential underestimation of PIP, as we have not included IV formulations due to limitation of our data (i.e., patients receiving IV formulation other than for cardiac stress may have been misclassified as not receiving PIP). | Fully applied |
| **2** | 5 | Avoid ticlopidine | Safer, effective alternatives available | PIP was defined as ≥1 prescription of triclopidine at the index date or previous 90 days. | | Fully applied |
| **2** | 6 | Avoid nitrofurantoin if creatinine clearance < 30ml/min or for long-term suppression of bacteria. To accommodate differences between the US and UK, we set creatinine clearance <45 mL/min as per the recommendation of NHS upon toxicity assessment.[6] | Potential pulmonary toxicity, hepatoxicity, peripheral neuropathy, especially with long-term use | PIP was defined as ≥1 prescription of nitrofurantoin at the index date or previous 90 days in patients with (i) ≥2 measurements of eGFR <45 mL/min in the year previous to index date, (ii) a diagnosis code for CKD stages 3B, 4 ,or 5 ever prior index, or (iii) if long-term suppression of bacteria (i.e., ≥ 1 prescription of nitrofurantoin with total length >180 days, allowing for gap of maximum 14 days between prescriptions whenever patients had information on dosage, or minimal cumulative dose prescription (prescription of >9,000 mg) of nitrofurantoin within the aforementioned period whenever dosage information was missing. The minimal threshold of minimal cumulative dose prescription within 180 days was calculated based on the standard dosage recommended by NHS for long term nitrofurantoin use (i.e., 50 to 100 mg/day for long-term low dose prophylaxis of recurrent UTI).[6] | Potential overestimation of PIP as we used as a proxy for low creatinine clearance ≥2 eGFR measurements with value <45 mL/min in the year prior to index date or a diagnosis code for CKD stages 3B, 4, or 5 ever before the index. By using the minimal cumulative dose prescription approach, we may have misclassified patients receiving high doses of nitrofurantoin <6 months as PIP.<br>Although NHS recommends assessing toxicity every 3-6 months in patients receiving long-term nitrofurantoin, we have not assessed whether patients were checked on nitrofurantoin toxicity in the assessment of PIP due to the limitations of our dataset. | Fully applied |
| **2** | 7 | Avoid peripheral alpha-1 blockers (doxazosin, prazosin, and terazosin) as an antihypertensive | High risk of orthostatic hypotension; | PIP was defined as ≥1 prescription of alpha-1 blockers at the index date or previous 90 days in, in patients with prior diagnosis of hypertension. | Potential overestimation of PIP. We have not assessed the indication of alpha-1 blockers (i.e., we assumed it was used in the treatment of hypertension whenever prescribed to patients with hypertension) | Fully applied |
| **2** | 8 | Avoid central alpha blockers (clonidine as first-line anti-hypertensive. Avoid guanabenz, guanfacine, methyldopa and reserpine [>0.1 mg/ day]) | High risk of adverse CNS effects, bradycardia, orthostatic hypotension; Not recommended as routine treatment for hypertension | PIP was defined as ≥1 prescription of (i) guanabenz, guanfacine, methyldopa, or reserpine at index or prior 90 days in patients with hypertension, or (ii) clonidine at index or prior 90 days in patients with hypertension and no prior use of antihypertensive drug. | | Fully applied |
| **2** | 9 | Avoid disopyramide | Potent negative inotrope and therefore may induce HF in older adults; strongly anticholinergic | PIP was defined as ≥1 prescription of disopyramide at the index date or previous 90 days. | | Fully applied |

**Table S1.** Continued.

| | | | | | | |
|---|---|---|---|---|---|---|
| **2** | 10 | Avoid dronedarone in individuals with permanent atrial fibrillation or severe or recently decompensated heart failure | Worse outcomes have been reported in patients taking dronedarone having permanent AF or severe/decompensated HF | PIP was defined as ≥1 prescription of dronedarone at the index date or previous 90 days with no prior diagnosis of permanent AF or severe/decompensated HF. | | Fully applied |
| **2** | 11 | Avoid digoxin as first-line therapy for AF | It may be associated with increased mortality | PIP was defined as ≥1 prescription of digoxin at the index date or previous 90 days in patients with prior diagnosis of AF and with no previous use of a first-line agent for rate control in AF as recommended by the NICE guidelines on management of AF[7] (i.e., standard BB [excl. sotalol] or CCB [verapamil and diltiazem]. | We may have misclassified patients receiving digoxin as first-line treatment for AF who had used a BB or CCB prior to AF event for other indications as not receiving PIP. Moreover, in patients receiving PIP, we have not assessed the reason for prescribing digoxin as first-line therapy due to limitations of our dataset (i.e., according to the NICE guidelines, digoxin monotherapy may be appropriate in people with no or very little physical exercise or when other rate-limiting drug options are ruled out because of comorbidities or the person's preferences). | Fully applied |
| **2** | 12 | Avoid digoxin as first-line therapy for HF | Questionable effects on risk of hospitalization and may be associated with increased mortality in older adults with HF | PIP was defined as ≥1 prescription of digoxin at the index date or previous 90 days in patients with prior diagnosis of HF (except if severe HF with reduced ejection fraction) and no previous use of other antiarrhythmics as recommended by the NICE guidelines on management of chronic HF[8] (i.e., BB [bisoprolol, carvedilol, and nebivolol], ARB, mineralocorticoid receptor antagonists, or ACE inhibitor). | We may have misclassified patients receiving digoxin as first-line treatment for HF who had used other antiarrhythmics medication prior to AF event for other indications as not receiving PIP. Moreover, we have not assessed the reason why patient received digoxin as first-line treatment due to limitation of our data. Nevertheless, digoxin is recommended by the NICE guidelines for worsening or severe HF with reduced ejection fraction despite first-line treatment for HF. | Fully applied |
| **2** | 13 | If digoxin is used for AF or HF, avoid dosages >125 mcg/day | Decreased real clearance of digoxin may lead to increased risk of toxic effects; | PIP was defined as ≥1 prescription of digoxin with daily dose >125 mcg at the index date or previous 90 days in patients with a diagnosis of HF or AF. | Potential underestimation of PIP as only patients having complete dosage information were included in the analysis. | Fully applied |
| **2** | 14 | Avoid nifedipine immediate release | Potential for hypotension; risk of precipitating myocardial ischemia | PIP was defined as ≥1 prescription of nifedipine immediate release at the index date or previous 90 days. | | Fully applied |

**Table S1.** Continued.

| | | | | | | |
|---|---|---|---|---|---|---|
| **2** | 15 | Avoid amiodarone as first-line therapy AF unless patient has HF or substantial left ventricular hypertrophy | Amiodarone is effective for maintaining sinus rhythm but has greater toxicities than other antiarrhythmics used in AF; it may be reasonable first-line therapy in patients with concomitant HF or substantial left ventricular hypertrophy if rhythm control is preferred over rate control. | PIP was defined as ≥1 prescription of amiodarone at the index date or previous 90 days in patients with a previous diagnosis of AF and no prior use of beta blockers (except sotalol) or CCB (verapamil and diltiazem) - except in patients with additional diagnosis of HF or left ventricular hypertrophy previous to index date). | We may have misclassified patients receiving amiodarone as first-line treatment for AF who had used a BB or CCB prior to AF event for other indications as not receiving PIP. | Fully applied |
| **2** | 16 | Avoid the following antidepressants, alone or in combination<br>Amitriptyline<br>Amoxapine<br>Clomipramine<br>Desipramine<br>doxepin (>6mg/day)<br>Imipramine<br>Nortriptyline<br>Paroxetine<br>Protriptyline<br>Trimipramine | Highly anticholinergic, sedating, and cause orthostatic hypotension; | PIP was defined as ≥1 prescription of the antidepressants at the index date or previous 90 days. | Potential overestimation of PIP. First, the use of tricyclic antidepressants is recommended in the management of depression by the NICE guidelines[9], and thus the prescription of these drugs it might not be considered inappropriate.<br>Second, we have not assessed the indication of the antidepressant (i.e., the NICE guidelines for management of neuropathic pain in adults in non-specialists settings recommends amitriptyline as initial treatment for neuropathic pain [except trigeminal neuralgia], and thus it would be appropriate).[10] | Fully applied |
| **2** | 17 | Avoid first- (conventional) and second- (atypical) generation antipsychotics, unless non-pharmacological options have failed and patient is threatening substantial harm to self or others | Increased risk of stroke and greater rate of cognitive decline and mortality in persons with dementia. Avoid antipsychotics for behavioural problems of dementia or delirium unless nonpharmacological options have failed or are not possible and the older adult is threatening substantial harm to self and others | PIP was defined as ≥1 prescription of antipsychotics at the index date or previous 90 days (except in patients with diagnosis of schizophrenia or bipolar disorder ever prior to index date). | Patients with a cancer diagnosis were excluded due to the study design (i.e., exclusion criteria); thus, the short-term use of antipsychotics during chemotherapy described in the criteria was not assessed in our analysis.<br>Potential overestimation of PIP as we have not assessed if patients had previously failed nonpharmacological options and if an older adult is threatening substantial harm to themselves and others due to limitations on the dataset. Moreover, some antipsychotics in the UK (e.g., clozapine) are exclusively prescribed in secondary care; therefore, we have no access to their prescription data. | Partially applied |

**Table S1.** Continued.

| | | | | | | |
|---|---|---|---|---|---|---|
| **2** | 18 | Avoid barbiturates<br>Amobarbital<br>Butabarbital<br>Butalbital<br>Mephobarbital<br>Pentobarbital<br>Phenobarbital<br>Secobarbital | High rate of physical dependence, tolerance to sleep benefits, great risk of overdose at low dosages | PIP was defined as ≥1 prescription of barbiturates at the index date or previous 90 days. | | Fully applied |
| **2** | 19 | Avoid short- and intermediate-acting BZD<br>Alprazolam<br>Estazolam<br>Lorazepam<br>Oxazepam<br>Temazepam<br>Triazolam | Older adults have increased sensitivity to BZD and decreased metabolism of long-acting agents; In general, all BZD increase risk of cognitive impairment, delirium, falls, fractures, and motor vehicle crashes in older adults | PIP was defined as ≥1 prescription of short- and intermediate-acting BZD at the index date or previous 90 days. To accommodate differences between the US and UK drug availability, we adapted the Beers criteria by including clobazam, ketazolam, and medazepam.[5] | | Fully applied |
| **2** | 20 | Avoid long-acting BZD<br>Clorazepate<br>Chlordiazepoxide<br>Clonazepam<br>Diazepam<br>Flurazepam<br>Quazepam | Elderly have decreased metabolism of long-acting BZDs. Increased risk of cognitive impairment, delirium, falls, fractures, motor vehicle crashes. May be appropriate for seizure disorders, rapid eye movement sleep disorders, BZD withdrawal, ethanol withdrawal, severe generalized anxiety disorder, and periprocedural anaesthesia. | PIP was defined as ≥1 prescription of long-acting BZD at the index date or previous 90 days. To accommodate differences between the US and UK drug availability, we adapted the Beers criteria by including bromazepam, lormetazepam, and nitrazepam.[5] | Potential overestimation of PIP. We have not assessed the indication of the BZD (i.e., it may be appropriate in seizure disorders, rapid eye movement sleep disorders, BZD withdrawal, ethanol withdrawal, severe generalized anxiety disorder, and periprocedural anaesthesia) due to limitations in our dataset. | Fully applied |
| **2** | 21 | Avoid meprobamate | High rate of physical dependence; very sedating | PIP was defined as ≥1 prescription of meprobamate at the index date or previous 90 days. | | Fully applied |
| **2** | 22 | Avoid nonBZD, BZD receptor agonist hypnotics<br>Eszopiclone<br>Zolpidem<br>Zaleplon | ADR similar to BZDs in elderly, delirium, falls, fractures, increased emergency department visits, hospitalizations, motor vehicle crashes | PIP was defined as ≥1 prescription of BZD-receptor agonists hypnotics at the index date or previous 90 days. To accommodate differences between the US and UK drug availability, we adapted the Beers criteria by including zopiclone.[5] | | Fully applied |

**Table S1.** Continued.

| | | | | | | |
|---|---|---|---|---|---|---|
| **2** | 23 | Avoid Ergoloid mesylates (dehydrogenated ergot alkaloids) Isoxsuprine | Lack of efficacy | PIP was defined as ≥1 prescription of Ergoloid mesylates or isoxsuprine at the index date or previous 90 days. | | Fully applied |
| **2** | 24 | Avoid androgens unless confirmed hypogonadism with clinical symptoms<br>Methyltestosterone<br>Testosterone | Potential for cardiac problems; contraindicated in men with prostate cancer | PIP was defined as ≥1 prescription of androgen at the index date or previous 90 days in male except if previous diagnosis of hypogonadism. | Potential overestimation of PIP. We have not considered hypogonadism symptoms (only the previous diagnosis) to assess the PIP. However, the hypogonadism diagnosis is made upon low serum testosterone levels and the presence of clinical symptoms. Thus, we considered that if previous a diagnosis recorded in the database, then androgen is appropriate. | Fully applied |
| **2** | 25 | Avoid desiccated thyroid | Concerns about cardiac affects | PIP was defined as ≥1 prescription of desiccated thyroid at the index date or previous 90 days. | | Fully applied |
| **2** | 26 | Avoid oral and topical patch estrogens with or without progestins (vaginal cream or tablets are acceptable to use low-dose intravaginal estrogen for management of dyspareunia, lower UTI and other vaginal symptoms). | Carcinogenic potential (breast and endometrium); lack of cardioprotective effect and cognitive protection in older women; Evidence indicates that vaginal estrogens for the treatment of vaginal dryness are safe and effective. | PIP was defined as ≥1 prescription of estrogen at the index date or previous 90 days. | Potential underestimation of PIP as we excluded all vaginal cream tablets formulations, regardless the dose or indication. | Fully applied |
| **2** | 27 | Avoid GH, except as hormone replacement after pituitary gland removal | Impact on body composition is small and associated with edema, arthralgia, carpal tunnel syndrome, gynecomastia, impaired fasting glucose | PIP was defined as ≥1 prescription of GH at the index date or previous 90 days (except if pituitary gland removal diagnosis recorded ever before). | | Fully applied |
| **2** | 28 | Avoid megestrol | Minimal effect on weight; increases risk of thrombotic events and possibly death in older adults | PIP was defined as ≥1 prescription of megestrol at the index date or previous 90 days. | | Fully applied |
| **2** | 29 | Avoid sulfonylureas long duration - chlorpropamide | Prolonged half-life in older adults; can cause prolonged hypoglycemia; causes syndrome of inappropriate antidiuretic hormone secretion | PIP was defined as ≥1 prescription of chlorpropamide at the index date or previous 90 days. | | Fully applied |

**Table S1.** Continued.

| | | | | | | |
|---|---|---|---|---|---|---|
| **2** | 30 | Avoid sulfonylureas long duration - glyburide | Higher risk of severe prolonged hypoglycemia in older adults | PIP was defined as ≥1 prescription of glyburide at the index date or previous 90 days. To accommodate differences between the US and UK drug availability, we adapted the Beers criteria by including glibenclamide.[5] | | Fully applied |
| **2** | 31 | Avoid metoclopramide unless gastroparesis | Can cause extrapyramidal symptoms, including tardive dyskinesia; risk may be greater in frail older adults | PIP was defined as ≥1 prescription of metoclopramide at the index date or previous 90 days (except in patients with prior diagnosis of gastroparesis) | | Fully applied |
| **2** | 32 | Avoid mineral oil given orally | Potential for aspiration and adverse events | PIP was defined as ≥1 prescription of mineral oil (oral) at the index date or previous 90 days. | | Fully applied |
| **2** | 33 | Avoid PPI use for >8weeks, unless for high risk patients (e.g., oral corticosteroids or chronic NSAID use), erosive esophagitis, Barrett's esophagitis, pathological hypersecretory condition, or demonstrated need for maintenance treatment (e.g., due to failure of drug discontinuation trial or H2-receptor blockers) | Risk of *Clostridium difficile* infection and bone loss and fractures | PIP was defined as ≥1 prescription of PPI with a total length of >56 days at the index date or previous 90 days, allowing for a gap of maximum 7 days between prescriptions when dosage information is available, or (ii) minimal cumulative dose prescription (see below) of a PPI within the period above whenever dosage information was missing:<br><br>Omeprazole: 56 days*20 mg (>1,120 mg prescribed)<br>Lansoprazole: 56 days*30 mg (>1,680 mg prescribed)<br>Esomeprazole: 56 days*30 mg (>1,680 mg prescribed)<br>Pantoprazole: 56 days*40 mg (>2,240 mg prescribed)<br>Rabeprazole: 56 days*20 mg (>1,120 mg prescribed)<br>Except if patients fulfil ≥1 following condition(s):<br>(iii) concomitant use of oral corticosteroids (i.e., ≥1 prescription at index or previous 90 days, regardless of prescription length), (iv) prior use of H2-receptor blocker, (v) prior diagnosis of erosive esophagitis or Barrett's esophagitis prior to/at PPI prescription, (vii) pathological hypersecretory condition [i.e., Zollinger-Ellison syndrome], or (viii) concomitant use of NSAID (i.e., ≥1 prescription with total length >90 days allowing for a gap of maximum 7 days between prescriptions when dosage information is available, or minimal cumulative dose prescription (see below) of a NSAID within the period above whenever dosage information was missing:<br>(cont.) | We assumed that patients with ≥1 prescription of corticosteroids within 90 days had concomitant use with PPI. Also, we have only included corticosteroids and oral formulations of corticosteroids.<br>Due to data limitations, we cannot access the reason for drug discontinuation (e.g., treatment failure). Thus, we assumed that if a patient had previously used an H2-receptor blocker, it was discontinued due to treatment failure. Similarly, we have not assessed if failure in discontinuing PPIs previously due to limitations in our dataset.<br>Potential PIP overestimation. By using the minimal cumulative dose prescription approach (as a proxy for use of PPI >56 days) we may have misclassified patients receiving high doses of PPI for <8 weeks as receiving PIP. Moreover, we defined drug use >8 weeks as the prescription of >56 days of the drug, considering patients received a pack size for 56 days of treatment. Thus, patients receiving a prescription for 8 weeks with a pack size of 60 days may be misclassified as PIP (applicable for NSAID use >90 days). | Partially applied |

**Table S1.** Continued.

| | | | | | | |
|---|---|---|---|---|---|---|
| **2** | 33 | | | (cont.)<br>Ibuprofen: 90 days*. 1,2 g (>108 g prescribed)<br>Naproxen: 90 days* 500 mg (>45,000 mg prescribed)<br>Etodolac 600mg: 90 days* 400 mg (>36,000 mg prescribed)<br>Meloxican: 90 days*. 15 mg (>1,350 mg prescribed)<br>Diclofenac: 90days* 100 mg (>9,000 mg prescribed)<br>Etoricoxib: 90 days* 60 mg (>5,400 mg prescribed)<br>Mefenamic acid: 90 days* 1 g (>90 g prescribed)<br>Nabumetone: 90 days* 1 g (>90g prescribed)<br>Indometacin: 90 days*100 mg (>9,000 mg prescribed)<br>Tolfenamic acid: 90 days* 300 mg (>27,000 mg prescribed) | | |
| **2** | 34 | Avoid meperidine, especially in individuals with CKD | Not effective in dosages commonly used; may have higher risk of neurotoxicity than other opioids | PIP was defined as ≥1 prescription of meperidine at the index date or previous 90 days. | | Fully applied |
| **2** | 35 | Avoid NSAID chronic use unless other alternatives are not effective and patient can take gastroprotective agent.<br>Aspirin >325 mg/day<br>Diiclofenac<br>Diflunisal<br>Etodolac<br>Fenoprofen<br>Ibuprofen<br>Ketoprofen<br>Meclofenamate<br>Mefenamic acid<br>Meloxicam<br>Nabumetone<br>Naproxen<br>Oxaprozin<br>Piroxicam<br>Sulindac<br>Tolmetin | increased risk of GI bleeding or peptic ulcer disease in high risk groups, including those aged >75 or taking oral or parenteral corticosteroids anticoagulants, or antiplatelets; Use of PPI or misoprostol reduces but do not eliminate risk. Upper GI ulcers, gross bleeding, perforation caused by NSAID occur in ~ 1% of patients treated for 3-6 months and in 2-4% of patients treated for 1 year; these trends continue with longer duration of use. | PIP was defined as ≥1 prescription of NSAID with total length >90 days at the index date or previous 90 days allowing for a gap between prescription of up to 7 days when information on dosage is available, or minimal cumulative dose prescription (see below) of a NSAID within the period above whenever dosage information was missing, except if ≥1 prescription of PPI or misoprostol within 90-days interval.<br><br>Ibuprofen: 90 days* 1,2 g (>108g prescribed)<br>Naproxen: 90 days* 500 mg (>45,000 mg prescribed)<br>Etodolac: 90 days* 400 mg (>36,000 mg prescribed)<br>Meloxican: 90 days* 15 mg (>1,350 mg prescribed)<br>Diclofenac: 90 days* 100 mg (>9,000 mg prescribed)<br>Mefenamic acid: 90 days* 1 g (>90 g prescribed)<br>Nabumetone: 90 days*1 g (>90 g prescribed) | Chronic use of NSAID was defined as use >90 days. Potential overestimation of PIP, as we have not assessed whether other alternatives were effective due to limitations of the dataset. | Partially applied |

**Table S1.** Continued.

| | | | | | | |
|---|---|---|---|---|---|---|
| **2** | 36 | Avoid indomethacin, ketorolac (including parenteral) | Indomethacin is more likely than other NSAID to have adverse CNS effects. Increased risk of GI bleeding, peptic ulcer disease, and acute kidney injury in older adults | PIP was defined as ≥1 prescription of indomethacin or ketorolac at the index date or previous 90 days. | | Fully applied |
| **2** | 37 | Avoid pentazocine | Opioid analgesic that causes CNS adverse effects more commonly than other opioids analgesic drugs; is also a mixed agonist and antagonist | PIP was defined as ≥1 prescription of pentazocine at the index date or previous 90 days. | | Fully applied |
| **2** | 38 | Avoid skeletal muscle relaxants<br>Carisoprodol<br>Chlorzoxazone<br>Cyclobenzaprine<br>Metaxalone<br>Methocarbamol<br>Orphenadrine | Most muscle relaxants poorly tolerated by older adults because some have anticholinergic adverse effects, sedation, increased risk of fractures; effectiveness at dosages tolerated by older adults questionable | PIP was defined as ≥1 prescription of skeletal muscle relaxants at the index date or previous 90 days. | | Fully applied |
| **2** | 39 | Avoid desmopressin for treatment of nocturia or nocturnal polyuria | High risk of hyponatremia | PIP was defined as ≥1 prescription of desmopressin at the index date or previous 90 days in patients with a previous diagnosis of nocturia or nocturnal polyuria. | | Fully applied |
| **3** | 40 | In patients with HF, avoid<br>(i) NSAID, COX-2 inhibitors, thiazolidinediones (pioglitazone, rosiglitazone), and cilostazol,<br>(ii) dronedarone in severe or recently decompensated HF, and<br>(iii) nondihydropyridine CCB (diltiazem, verapamil) in patients with HFrEF. | Potential to promote fluid retention and exacerbate HF | PIP was defined as ≥1 prescription of the listed drugs at the index date or previous 90 days in patients with previous diagnosis of HF. Additionally, in (ii) patients should have a diagnosis of severe/ recently decompensated HF recorded at dronedarone prescription or prior 90 days (i.e., proxy for severe or recently decompensated HF). | Potential underestimation of PIP as we have not assessed (iii) due to limitations on the database (i.e., no diagnosis code for HFrEF in the dataset, as patients with HFrEF are managed by secondary care in the UK). | Partially applied |
| **3** | 41 | In patients with syncope, avoid<br>AchEi<br>Peripheral alpha-1 blockers<br>Doxazosin<br>Prazosin<br>Terazosin<br>(cont.) | Increases risk of orthostatic hypotension or bradycardia | PIP was defined as ≥1 prescription of AChEi, peripheral alpha-1 blockers (doxazosin, prazosin, terazosin), tertiary TCAs, chlorpromazine, thioridazine, or olanzapine at the index date or previous 90 days in patients with a diagnosis for syncope ever prior to drug prescription. | | Fully applied |

**Table S1.** Continued.

| | | | | | | |
|---|---|---|---|---|---|---|
| **3** | 41 | (cont.)<br>Tertiary TCA<br>Chlorpromazine<br>Thioridazine<br>Olanzapine | | | | |
| **3** | 42 | Avoid<br>Bupropion<br>Chlorpromazine<br>Clozapine<br>Maprotiline<br>Olanzapine<br>Thioridazine<br>Thiothixene<br>Tramadol<br>if chronic seizures or epilepsy | Lowers seizure threshold; may be acceptable in patients with will controlled seizures in whom alternative agents have not been effective. | PIP was defined as ≥1 prescription of at least one of the drugs listed at the index date or previous 90 days in patients with chronic seizures and epilepsy. Patients with well controlled seizures (i.e., no diagnosis of seizures in the previous 6 months to index as a proxy for no epileptic event and ≥1 prescription of anti-seizure medication within 90-days previous to index) were excluded. | Potential overestimation of PIP. We have not assessed if alternative agents failed previously due to limitation of the database and different indications (e.g., antidepressants, antipsychotics, and opioids). Moreover, we used a combination of absence of diagnosis record of seizure or epileptic event within 6 months prior to index and ≥1 prescription of anti-seizure medication at index or prior 90 days as a proxy for well controlled epilepsy. Thus, we assumed that if a patient had no diagnosis of epilepsy within 6 months prior to index and had co-medication for epilepsy, then drug prescription was not inappropriate.<br>Potential underestimation of PIP as some antipsychotics in the UK (e.g., clozapine) are exclusively prescribed in secondary care; therefore, we have no access to their prescription data. | Fully applied |
| **3** | 43 | In patients with or at risk of delirium avoid<br>(i) Anticholinergics<br>BZD<br>Chlorpromazine<br>Corticosteroids<br>H2-receptor blockers<br>Cimetidine<br>Famotidine<br>Nizatidine<br>Ranitidine<br>Meperidine<br>Sedative hypnotics (cont.) | Potential of these drugs of inducing or worsening delirium. Avoid antipsychotics for behavioural problems of dementia or delirium unless nonpharmacological options have failed or are not possible and the older adult is threatening substantial harm to self and others. Antipsychotics are associated with greater risk of stroke and mortality in persons with dementia. | PIP was defined as ≥1 prescription of ≥1 listed drug at the index date or previous 90 days in patients with diagnosis of delirium in the previous year. Additionally, patients receiving antipsychotics were defined as PIP if they had a previous diagnosis of dementia. | Potential underestimation of PIP as only patients having a diagnosis of delirium prior to index were included in the analysis due to limitations of our data on capturing the risk of delirium. Moreover, some antipsychotics in the UK (e.g., clozapine) are exclusively prescribed in secondary care; therefore, we have no access to their prescription data.<br>Potential overestimation of PIP as we have not assessed if other non-pharmacological options have failed previously or were not possible, as well as if older adults represented substantial harm to self or others (not well captured in the database). | Partially applied |

**Table S1.** Continued.

| | | | | | | |
|---|---|---|---|---|---|---|
| **3** | 43 | (cont.) avoid (ii) antipsychotics for behavioural problems of dementia or delirium. | | | | |
| **3** | 44 | In patients with dementia or cognitive impairment avoid<br>Anticholinergics<br>Antipsychotics<br>H2-receptor antagonist<br>BZD<br>Non-BZD, BZD receptor agonist hypnotics | Avoid because of CNS effects; Avoid antipsychotics for behavioural problems of dementia unless nonpharmacological options have failed or are not possible and the older adult is threatening substantial harm to self and others.<br>Antipsychotics are associated with greater risk of stroke and mortality in persons with dementia. | PIP was defined as ≥1 prescription of anticholinergics, BZD, H2-receptor antagonists, Non-BZDs BZD receptor agonist hypnotics or antipsychotics at the index date or previous 90 days in patients with diagnosis of dementia or cognitive impairment at or prior to index date. | Potential overestimation of PIP as we have not assessed if other non-pharmacological options have failed previously or were not possible, as well as if older adults represented substantial harm to self or others (not well captured in the database).<br>Potential underestimation of PIP as some antipsychotics in the UK (e.g., clozapine) are exclusively prescribed in secondary care; therefore, we have no access to their prescription data. | Fully applied |
| **3** | 45 | In patients with history of falls or fractures, avoid<br>(i) Anticonvulsants (except for seizure and mood disorders),<br>(ii) Antipsychotics, BZD, non-BZD, BZD receptor agonist hypnotics, TCA, SSRI unless safer alternatives are not available, and<br>(iii) Opioids (except for pain management due to recent fractures or joint replacement). | May cause ataxia, impaired psychomotor function, syncope, additional falls; shorter-acting BZDs are not safer than long acting ones. If one of the drugs must be used consider reducing use of other CSN-active medications that increase risk of falls and fractures (i.e., anticonvulsants, opioid-receptor agonists, anti-psychotics, antidepressants, BZDs-receptor agonists, other sedatives and hypnotics) and implement other strategies to reduce fall risk | PIP was defined as ≥1 prescription of ≥1 drug listed in patients with fracture and/or falls history at the index date or previous 90 days (except if patients with anticonvulsants had an ever-prior diagnosis of seizures or mood disorders, or patients with opioids had a diagnosis of fracture or joint replacement in the previous 30 days). | In patients receiving opioids or anticonvulsants, we could not assess if GPs considered reducing the use of other CSN-active medications that increase the risk of falls and fractures (i.e., antipsychotics, antidepressants, BZDs-receptor agonists, other sedatives and hypnotics) as well as if they implemented other strategies to reduce the risk of fall. Moreover, due to limitations on the database we could not assess if safer alternatives to antipsychotics, BZDs, non-BZDs/BZDs receptor agonist hypnotics, TCAs, or SSRI.<br>Potential underestimation of PIP as some antipsychotics in the UK (e.g., clozapine) are exclusively prescribed in secondary care; therefore, we have no access to their prescription data. | Partially applied |
| **3** | 46 | In patients with insomnia avoid<br>Oral decongestants<br>Pseudoephedrine<br>Phenylephrine (cont.) | CNS stimulants effect | PIP was defined as ≥1 prescription of oral decongestants, stimulants, or theobromines at the index date or previous 90 days in patients with a diagnosis of insomnia in the previous year. | | Fully applied |

**Table S1.** Continued.

| | | | | | | |
|---|---|---|---|---|---|---|
| **3** | 46 | (cont.)<br>Stimulants<br>Amphetamine<br>Armodafinil<br>Methylphenidate<br>Modafinil<br>Theobromines<br>Theophylline<br>Caffeine | | | | |
| **3** | 47 | In patients with Parkinson's disease avoid<br>Antipsychotics (except aripiprazole, quetiapine, and clozapine)<br>Antiemetics<br>Metoclopramide<br>Prochlopramide<br>Promethazine | Dopamine-receptor antagonists with potential to worsen parkinsonian symptoms;<br>Quetiapine, aripiprazole, clozapine, appear to be less likely to precipitate worsening of Parkinson disease | PIP was defined as ≥1 prescription of an antipsychotic or antiemetics listed at the index date or previous 90 days in patients with a previous diagnosis of Parkinson's disease. | Potential underestimation of PIP as some antipsychotics in the UK are exclusively prescribed in secondary care; therefore, we have no access to their prescription data. | Fully applied |
| **3** | 48 | Avoid aspirin (>325 mg/day) and non-COX2 NSAID in patients with history of gastric ulcers unless other alternatives are not effective and patient can take gastroprotective agent (PPI or misoprostol). | May exacerbate existing ulcers or cause new or additional ulcers | PIP was defined as ≥1 prescription of aspirin (>325 mg/day) and non-COX2 selective NSAID at the index date or previous 90 days in patients with a diagnosis for gastric ulcer ever prior to drug prescription without gastric protection (i.e., >1 prescription of PPI or misoprostol at index date or previous 90 days) | We assumed that if a patient could take a gastroprotective agent, it was co-prescribed together aspirin or non-COX2 selective NSAIDs. Moreover, we assumed that a prescription of PPI or misoprostol within the 90-days interval meant that patients were using concomitantly to aspirin or non-COX2 selective NSAIDs.<br>Potential overestimation of PIP as we could not assess the indication or weather previous alternatives were not effective; | Partially applied |
| **3** | 49 | Avoid NSAID (non-COX and COX-selective, oral and parenteral) in CKD stages IV or V (creatinine clearance <30 mL/min) | May increase risk of acute kidney injury and further decline of renal function | PIP was defined as ≥1 prescription of NSAID at the index date or previous 90 days in patients with a previous diagnosis of CKD stages IV or V (ever prior to index date) and/or patients with ≤2 eGFR measurements ≤30mL/min in the previous year. | | Fully applied |
| **3** | 50 | In women with urinary incontinency avoid<br>(cont.) | Aggravation of incontinence | PIP was defined as ≥1 prescription of estrogen or peripheral alpha-1 blockers at the index date or previous<br>(cont.) | | Fully applied |

**Table S1.** Continued.

| | | | | | | |
|---|---|---|---|---|---|---|
| **3** | 50 | (cont.) Estrogen oral and transdermal (exclude intravaginal)<br>Peripheral alpha-1 blockers<br>Doxazosin<br>Prazosin<br>Terazosin | | (cont.) 90 days in women with a diagnosis of urinary incontinency in the prior 5 years. | | |
| **3** | 51 | Avoid strong anticholinergics drugs (except antimuscarinics for urinary incontinence) in men with lower urinary tract symptoms or BPH | May decrease urinary flow and cause urinary retention | PIP was defined as ≥1 prescription of strong anticholinergics drugs at the index date or previous 90 days in men with ≥1 lower UT symptom diagnosis recorded in the last year, or diagnosis of BPH in the previous 5 years. | | Fully applied |
| **5** | 52 | Avoid use of ACE inhibitors and amiloride or triamterene; | Increased risk of hypercalcemia | PIP was defined as ≥1 prescription of ACE inhibitors and ≥1 prescription of amiloride or triamterene at the index date or previous 90 days in patients without a diagnosis code of hypocalcemia (or potassium measurement <3.5 mmol/L) at index or prior 6 months to index date. | Potential overestimation of PIP as we assumed that ACE inhibitors and amiloride or triamterene were concomitantly used as long as the two drugs were prescribed within the 90-days period. | Fully applied |
| **5** | 53 | Avoid using two or more anticholinergic drugs | Increased risk of cognitive decline | PIP was defined as concomitant use of ≥2 anticholinergic drugs at the index date or previous 90 days. Concomitant use was defined as ≥1-day prescription overlap of >2 drugs. | Potential overestimation of PIP as we assumed that two or more anticholinergic drugs were concomitantly used as long as the they were prescribed within the 90-days period. | Fully applied |
| **5** | 54 | Avoid antidepressants (i.e., SSRI or TCA) and at least two other CNS-active drugs | Increased risk of falls | PIP was defined as ≥1 prescription of SSRI or TCA at the index date or previous 90 days and ≥2 other CNS-active drugs (i.e., distinct ATC codes at 5th level) prescribed within the 90-days period. | Potential overestimation of PIP as we assumed that SSRI or TCA and the other CNS active drugs were concomitantly used as long as the drugs were prescribed within the 90-days period. | Fully applied |
| **5** | 55 | Avoid antipsychotics and two or more CNS-active drugs | Increased risk of falls | PIP was defined as ≥1 prescription of antipsychotics at the index date or previous 90 days with ≥2 other CNS-active drugs (i.e., distinct ATC codes at 5th level) prescribed within the 90-days period. | Potential overestimation of PIP as we assumed that antipsychotics and the other CNS active drugs were concomitantly used as long as the two drugs were prescribed within the 90-days period. | Fully applied |
| **5** | 56 | Avoid prescribing BZD and non-BZD, BZD receptor agonist hypnotics and at least two additional CNS-active drugs | Increased risk of falls and fractures | PIP was defined as ≥1 prescription of BZD and/or non-BZD, BZD hypnotics at the index date or previous 90 days with ≥2 other CNS-active drugs (i.e., distinct ATC codes at 5th level) prescribed within the 90-days period. | Potential overestimation of PIP as we assumed that two or more drugs were concomitantly used as long as the they were prescribed within the 90-days period. | Fully applied |

**Table S1.** Continued.

| | | | | | | |
|---|---|---|---|---|---|---|
| **5** | 57 | Avoid corticosteroids + NSAID, if not possible, provide GI protection (PPI or misoprostol) | Increased risk of peptic ulcer disease or GI bleeding | PIP was defined as ≥1 prescription of corticosteroids at the index date or previous 90 days with additional ≥1 prescription of NSAID within the 90-days period and without use of PPI, or misoprostol within the 90-days period. | Potential overestimation of PIP as we assumed that NSAIDs and corticosteroids were concomitantly used as long as the two drugs were prescribed within the 90-days period. | Fully applied |
| **5** | 58 | Avoid lithium + ACE inhibitors, monitor lithium concentration | Increased risk of lithium toxicity | PIP was defined as ≥1 prescription of lithium and ≥1 prescription of ACE inhibitor at index date or 90 days previous to index in patients without lithium monitoring. Lithium monitoring was defined by ≥1 record of a diagnosis code for lithium monitoring or a lab measurement (regardless result value) within 6-months previous to index date. | Potential overestimation of PIP as we assumed that lithium and ACE inhibitors were concomitantly used as long as the two drugs were prescribed within the 90-days period. | Fully applied |
| **5** | 59 | Avoid lithium + loop diuretics, monitor lithium concentration | Increased risk of lithium toxicity | PIP was defined as ≥1 prescription of lithium and ≥1 prescription of loop diuretic at index date or 90 days previous to index in patients without lithium monitoring. Lithium monitoring was defined by ≥1 record of a diagnosis code for lithium monitoring or a lab measurement (regardless result value) within 6-months previous to index date. | Potential overestimation of PIP as we assumed that lithium and loop diuretics were concomitantly used as long as the two drugs were prescribed within the 90-days period. | Fully applied |
| **5** | 60 | Avoid opioid receptor agonist analgesics + ≥2 other CNS-active drugs | Increased risk of falls | PIP was defined as ≥1 prescription of opioids at the index date or previous 90 days with ≥2 other CNS-active drugs (i.e., distinct ATC codes at 5th level) prescribed within the 90-days period. | Potential overestimation of PIP as we assumed that opioids and the additional CNS active drugs were concomitantly used as long as the drugs were prescribed within the 90-days period. | Fully applied |
| **5** | 61 | Avoid peripheral alpha-1 blockers in combination with loop diuretics in older women unless conditions warrant both drugs | Increased risk of urinary incontinence in older women | PIP was defined as ≥1 prescription of loop diuretics and ≥1 prescription alpha-1 blockers at index date or previous 90 days, except in patients with previous use of CCB + thiazide-like diuretic + ARB + ACE inhibitors (as per recommendation of NICE guidelines to manage adults with hypertension)[11] | Potential overestimation of PIP as we assumed that alpha-1 blockers and loop diuretics were concomitantly used as long as the two drugs were prescribed within 90 days.<br>Moreover, we assumed that the previous use of CCB + thiazide-like diuretic + ARB or ACE inhibitors (ever prior index) warranty a condition for appropriate prescription of loop diuretic and alpha-1-blocker. | Fully applied |
| **5** | 62 | Avoid theophylline in combination with cimetidine | Increased risk of theophylline toxicity | PIP was defined as ≥1 prescription of theophylline and ≥1 prescription of cimetidine at index date or 90 days previous to index. | Potential overestimation of PIP as we assumed that theophylline and cimetidine were concomitantly used as long as the two drugs were prescribed within the 90-days period | Fully applied |

**Table S1.** Continued.

| | | | | | | |
|---|---|---|---|---|---|---|
| **5** | 63 | Avoid warfarin in combination with amiodarone when possible; monitor international normalized ratio closely | Increased risk of bleeding | PIP was defined as ≥1 prescription of warfarin and ≥1 prescription amiodarone at index date or previous 90 days without diagnosis code for warfarin monitoring or INR monitoring, or ≥1 INR measurement (regardless the value measured) recorded within the 90-days period. | Potential overestimation of PIP as we assumed that warfarin and amiodarone were concomitantly used as long as the two drugs were prescribed within the 90-days period. Moreover, we assumed that PIP occurs if no INR monitoring during a 90-days period when using the combination of drugs. | Fully applied |
| **5** | 64 | Avoid warfarin in combination with non-steroidal anti-inflammatory drugs when possible; if used together, monitor bleeding closely | Increased risk of bleeding | PIP was defined as ≥1 prescription of warfarin and ≥1 prescription NSAID at index date or previous 90 days without diagnosis code for warfarin monitoring or INR monitoring, or ≥1 INR measurement (regardless the value measured) recorded within the 90-days period. | Potential overestimation of PIP as we assumed that warfarin NSAID were concomitantly used as long as the two drugs were prescribed within the 90-days period. | Fully applied |
| **6** | 65 | Avoid amiloride if creatinine clearance <30 mL/min | Increased potassium and decreased sodium | PIP was defined as ≥1 prescription of amiloride at the index date or previous 90 days in patients with ≥2 measurements of eGFR <30 mL/min in the year previous to index date or a diagnosis code for CKD stages 4 or 5 ever prior index. | | Fully applied |
| **6** | 66 | Avoid apixaban if creatinine clearance <25 mL/min | Increased risk of bleeding | PIP was defined as ≥1 prescription of apixaban at the index date or previous 90 days in patients with ≥2 eGFR measurement <25 mL/min within 1-year period prior to the index date. | | Fully applied |
| **6** | 67 | Avoid dabigatran if creatinine clearance <30 mL/min | Increased risk of bleeding | PIP was defined as ≥1 prescription of dabigatran at the index date or previous 90 days in patients with ≥2 eGFR measurement <30 mL/min in the year previous to index date or a diagnosis code for CKD stage 5 ever prior index. | | Fully applied |
| **6** | 68 | Avoid edoxaban if creatinine clearance <30 or >95 mL/min | Increased risk of bleeding | PIP was defined as ≥1 prescription of edoxaban at the index date or previous 90 days in patients with ≥2 eGFR measurement <30 or >95 mL/min within 1-year period prior to the index date or a diagnosis code for CKD stage 5 ever prior index. | | Fully applied |
| **6** | 69 | Avoid fondaparinux if creatinine clearance <30 mL/min | Increased risk of bleeding | PIP was defined as ≥1 prescription of fondaparinux at the index date or previous 90 days in patients with ≥2 eGFR measurement <30 mL/min in the year previous to index date or a diagnosis code for CKD stage 5 ever prior index. | | Fully applied |

**Table S1.** Continued.

| | | | | | |
|---|---|---|---|---|---|
| 6 | 70 | Avoid rivaroxaban if creatinine clearance <30 mL/min | Increased risk of bleeding | PIP was defined as ≥1 prescription of rivaroxaban at the index date or previous 90 days in patients with ≥2 eGFR measurement <30 mL/min in the year previous to index date or a diagnosis code for CKD stage 5 ever prior index. | Fully applied |
| **6** | 71 | Avoid spironolactone if creatinine clearance <30 mL/min | increased potassium | PIP was defined as ≥1 prescription of spironolactone at the index date or previous 90 days in patients with ≥2 eGFR measurement <30 mL/min in the year previous to index date or a diagnosis code for CKD stage 5 ever prior index. | Fully applied |
| **6** | 72 | Avoid triamterene if creatinine clearance <30 mL/min | increased potassium and decreased sodium | PIP was defined as ≥1 prescription of triamterene at the index date or previous 90 days in patients with ≥2 eGFR measurement <30 mL/min in the year previous to index date or a diagnosis code for CKD stage 5 ever prior index. | Fully applied |
| **6** | 73 | Avoid duloxetine if creatinine clearance <30 mL/min | increased GI adverse effects (nausea, diarrhea) | PIP was defined as ≥1 prescription of duloxetine at the index date or previous 90 days in patients with ≥2 eGFR measurement <30 mL/min in the year previous to index date or a diagnosis code for CKD stage 5 ever prior index. | Fully applied |
| **6** | 74 | Avoid tramadol extended release if creatinine clearance <30 mL/min | CNS adverse effects | PIP was defined as ≥1 prescription of tramadol modified-release formulations at the index date or previous 90 days in patients with ≥2 eGFR measurement <30 mL/min in the year previous to index date or a diagnosis code for CKD stage 5 ever prior index. | Fully applied |
| **6** | 75 | Avoid probenecid if creatinine clearance <30 mL/min | Loss of effectiveness | PIP was defined as ≥1 prescription of probenecid at the index date or previous 90 days in patients with ≥2 eGFR measurement <30 mL/min in the year previous to index date or a diagnosis code for CKD stage 5 ever prior index. | Fully applied |

[1] Original table number from the 2015 Beers criteria (doi: 10.1111/jgs.13702).

**Abbreviations:** ACE: Angiotensin converting enzyme; AchEi: acetylcholinesterase inhibitors; AF: Atrial fibrillation; AH: Anti-histamine; ARB: Angiotensin II receptor blockers; BB: Beta blockers; BPH: Benign prostatic hyperplasia; BZD: Benzodiazepines; CCB: Calcium-channel blockers; CKD: Chronic kidney disease; CNS: Central nervous system; HFrEF: Heart failure with reduced ejection fraction; eGFR: Estimated glomerular filtration rate; GH: Growth hormone; GI: Gastrointestinal; GP: General practitioner; HF: Heart failure; INR: International normalized ratio; IV: Intravenous; NHS: National Health Service; NICE:  National Institute for the Heath and Care Excellence; NSAID: Non-steroidal anti-inflammatory drugs; OTC: Over the counter; PIP: Potential inappropriate prescription; PPI: Proton pump inhibitors; SSRI: Selective serotonin reuptake inhibitor; TCA: Tricyclic antidepressants; UK: United Kingdom; UTI: Urinary tract infections;

**Table S2.** List of Prescribing Optimally in Middle-aged People's Treatments (PROMPT) criteria used for the assessment of potential inappropriate prescription (PIP) in middle-aged adults.

| Criteria Number | Recommendation | Rationale | PIP definition | Limitations | Criteria Applicability |
|---|---|---|---|---|---|
| **1** | Other than for opioid-induced constipation, stimulant laxatives should not be prescribed as first-line treatment in constipation for >4 weeks. | Stimulant laxatives are not suitable for continuous long-term use, other than for opioid induced constipation. | ≥1 prescription of stimulant laxatives at or within 90-days to index date, without a prescription of opioid drug and no previous use of other laxative drug prior 5 years to the index date (proxy for first line treatment). Use of stimulant laxatives ≥4 weeks was defined as (i) ≥1 prescription with cumulative length >28 days, allowing for a gap of up to 7 days between prescriptions, whenever information on dosage for all prescriptions was available, or (ii) minimal cumulative dose prescription (see below) of a stimulant laxative drug for constipation within the aforementioned period when dosage information was missing:<br>▪ Senna all formulations: 28 days* 7.5 mg (prescription of >210 mg)<br>▪ Senna fruit 12.4%: 28 days* 4 g sachet (prescription of >112 g)<br>▪ Bisacodyl oral formulations: 28 days* 10 mg (prescription of >280 mg)<br>▪ Bisacodyl suppositories: 28 days* 10 mg (prescription of >280 mg)<br>▪ Sodium picosulfate all formulations: 28 days* 5 mg (prescription of >140 mg) | Potential underestimation of PIP, as we look back 5 years prior to the index date for assessing previous use of other laxative drugs.<br>Potential overestimation of PIP by using a minimal cumulative dose prescription approach in patients with missing information on dosage (e.g., patients using high dose of laxatives for short time may be misclassified as having PIP). Moreover, PIP may be overestimated as we defined the drug use >4 weeks equal to prescription of >28 days of drug assuming that pack size is equal to 28 days of treatment (i.e., patients receiving a prescription for 4 weeks with a pack size of 30 days may be misclassified as PIP). | Fully applied |
| **2** | PPI should not be prescribed above recommended maintenance dose for >8 weeks, unless treatment is indicated for rare conditions (e.g., Zollinger-Ellison syndrome) | PPI can lead to adverse reactions such as headaches, dizziness, skin rash, abdominal pain, diarrhoea, back pain, and upper respiratory infections | PIP was defined as use of PPI in high doses (see below) for >8 weeks in absence of a rare condition (i.e., ≥1 prescription of a PPI with cumulative length of >56 days allowing for a gap of maximum 7 days between prescriptions). Only included patients having complete information on dosage.<br>High dose per PPI drug:<br>▪ omeprazole >20 mg/day<br>▪ lansoprazole >30 mg/day<br>▪ esomeprazole >20 mg/day<br>▪ pantoprazole >40 mg/day<br>▪ rabeprazole >20 mg/day | Potential underestimation of PIP due to inclusion of only complete cases in the analysis. Moreover, patients starting at normal dose and increasing the value above threshold over the 8 weeks period were not classified as PIP (i.e., only classified as PIP patients at high dose for at least >56 days).<br>PIP may be overestimated as we defined the drug use >8 weeks equal to prescription of >56 days of drug assuming (pack size for 56 days of treatment). Thus, patients receiving a prescription for 8 weeks with a pack size of 60 days may be misclassified as PIP. | Fully applied |
| **3** | Avoid omeprazole and esomeprazole in combination with clopidogrel | Reduction of antiplatelet effect that may lead to thrombosis | PIP was defined as the concomitant use of clopidogrel and esomeprazole or omeprazole. Concomitant use was defined as ≥1-day overlap prescription between the two drugs. | Potential underestimation of PIP as only patients with complete dosage information for at least one of the drugs were included. Potential overestimation of PIP as whenever the two drugs were not prescribed at the same day but had ≥1-day overlap, we assumed the first drug was not advised to be stopped at the prescription of the second drug. | Fully applied |

**Table S2.** Continued.

| | | | | | |
|---|---|---|---|---|---|
| **4** | Avoid use of AAB drugs as monotherapy for hypertension. | Increased risk of orthostatic hypotension | PIP was defined as AAB monotherapy (i.e., ≥1 prescription of AAB at or within 90-days to index date and no prescription of additional hypertensive drug in the 90-days period) in patients previously diagnosed with hypertension. | Potential overestimation of PIP as we assumed that a prescription of AAB and another anti-hypertensive drug within the 90-days period meant that patients were using the drugs concomitantly. | Fully applied |
| **5** | Aspirin doses should not exceed 150 mg/day for anti-platelet therapy | Risk of bleeding | PIP was defined as ≥1 prescription of aspirin with antiplatelet indication (ATC B01AC06) with dose >150 mg/ day. | Potential underestimation of PIP as only patients having complete dosage information were included in the analysis. Moreover, we used the ATC code as a proxy to ascertain the indication (we have not included aspirin formulations with other indication), and we assumed that all patients receiving aspirin ≤150 mg was for antiplatelet therapy. | Fully applied |
| **6** | Avoid cardio-selective CCB in combination with BB | Risk of atrioventricular block and risk of myocardial depression | PIP was defined as the concomitant use of CCB and BB. Concomitant use of drugs was defined as ≥1 day of use of both drugs. | Potential underestimation of PIP as only patients with complete dosage information on the two drugs prescribed on the same date were included. Potential overestimation of PIP as we assumed that patients having a prescription for CCB and BB with ≥1-day overlap received concomitantly the two drugs (we have not considered that physicians could have advised patients to stop one drug before initiating the other). | Fully applied |
| **7** | Avoid oral short-acting dipyridamole as monotherapy in antiplatelet treatment | Risk of orthostatic hypotension | PIP was defined as use of short-acting dipyridamole with no additional antiplatelet medication use. | Potential underestimation of PIP as we assumed that whenever patients received a prescription of dipyridamole with another antiplatelet drug within 90-days period the two drugs were used concomitantly. | Fully applied |
| **8** | Avoid first-generation AH drug as first-line treatment for >7 days. | May cause addiction and/or exert anticholinergic effects causing unwanted side effects (e.g., constipation, drowsiness, psychomotor impairment) | PIP was defined as ≥1 prescription of first-generation oral AH for >7 days, with no use of other AH drug within 1-year before index date. Only oral formulations were included in the analysis. We defined >7 days drug use as (i) ≥1 prescription with cumulative length >7 days, allowing for a gap of ≤1 day between prescriptions (when information on dosage is available), or (ii) minimal cumulative dose prescription (see below) of a first-generation AH drug within the aforementioned period whenever dosage information was missing:<br>Minimal threshold of cumulative drug prescribed (in mg per drug) within 90-day period previous to index.<br>▪ Promethazine hydrochloride all formulations: 7 days * 25 mg (prescription of >175 mg)<br>▪ Diphenhydramine 25 mg tablets: 7days * 200 mg (prescription of >1400 mg)<br>▪ Chlorphenamine all formulations: 7 days * 12 mg (prescription of >84 mg)<br>(cont.) | Potential overestimation of PIP as we assumed that first-generation AH was only used for short-term periods. Moreover, PIP may be overestimated as we defined the drug use >7 days (i.e., prescription of >7 days of drug) assuming that pack size is equal to a number that is multiple of 7. | Fully applied |

**Table S2.** Continued.

| | | | | | |
|---|---|---|---|---|---|
| **8** | | | (cont.)<br>▪ Alimemazine 10mg tablets: 7 days * 30 mg (prescription of ≥210 mg)<br>▪ Hydroxyzine all formulations: 7 days * 75 mg (prescription of ≥525 mg) | | |
| **9** | Avoid theophylline as monotherapy for asthma or COPD | Increased risk of arrhythmias | PIP was defined as ≥1 prescription of theophylline at index date or previous 90-days with no additional medication for asthma/COPD was prescribed in the 90-days period. | PIP potential overestimation as we assumed patients having a prescription for theophylline and other medication for asthma or COPD within the 90 days interval were used concomitantly. | Fully applied |
| **10** | Concomitant bisphosphonate should be prescribed if oral corticosteroids are used >3 months | Long-term use of an oral corticosteroids increases the risk of osteoporosis and subsequent bone fracture | The use of corticosteroids for >90 days was defined as (i) ≥1 prescription of corticosteroids with a total length of >90 days in patients with dosage information, or (ii) minimal cumulative dose prescription in mg (see below) of an oral corticosteroid drug within the period above whenever dosage information was missing. The cumulative dose of oral corticosteroids was calculated among current users and expressed as prednisone equivalents using an online tool.[1] The minimal threshold in prednisone equivalent was 900 mg (10 mg * 90 days) in patients with no dosage information. We have not included budesonide as it is a glucocorticoid with limited systemic bioavailability due to extensive (90%) first-pass hepatic metabolism by the cytochrome p-450 enzyme system.[2,3] These properties limit systemic adverse effects. | As we used the minimal cumulative dose prescription of prednisone equivalents in patients with missingness of dosage information, patients receiving high dosages for shorter time may be misclassified as having PIP. Potential overestimation of PIP, as we have not accounted for bisphosphonates administered in the hospital (e.g., zoledronic acid) due to limitations of the database. | Fully applied |
| **11** | Avoid mucolytic agents in stable COPD | There is little benefit from the use of mucolytic agents in stable COPD | PIP was defined as ≥1 prescription for mucolytic at index or previous 90 days in patients with stable COPD. Stable COPD was defined as no diagnosis record for exacerbations within 1-year previous to the mucolytic agent prescription. | Potential overestimation of PIP as we assumed that patients with no diagnosis code for exacerbation registered by the GP in the previous year had no exacerbation event, and therefore, were considered stable. | Fully applied |
| **12** | Avoid SSRI in combination with venlafaxine | Serotonin syndrome | PIP was defined as ≥1 prescription of SSRI and ≥1 prescription of venlafaxine with ≥1 day overlap treatment. | Potential overestimation of PIP as we assume patients with prescriptions overlapping at least one day were using the two drugs concomitantly (we have not considered that physicians could have advised patients to stop one drug before initiating another). Also, potential underestimation of PIP, as only patients with complete dosage information for at least one of the two drugs were included in the analysis. | Fully applied |
| **13** | Avoid TCA as first-line treatment of depression | Anticholinergic effects; constipation; dry mouth; drowsiness | PIP was defined as ≥1 TCA prescription at index date or previous 90 days in patients with a previous diagnosis of depression who had not used other AD after diagnosis of depression (i.e., proxy for TCA as first line therapy for depression). | Potential underestimation of PIP. In the UK, antidepressants such as TCA are also used as treatment for chronic primary pain in patients over 16 years old.[4] Therefore, we have only included in the analysis patients with a previous diagnosis of depression in the database. If multiple diagnosis were recorded in the database, we used the first one. Moreover, we assumed that patients receiving TCA or other antidepressant before the diagnosis had a different (cont.) | Fully applied |

**Table S2.** Continued.

| | | | | | |
|---|---|---|---|---|---|
| **13** | | | | (cont.) indication than treating depression (e.g., pain management), and thus, should not rule out the potential for misclassification of patients that received other antidepressants with indication to treat this condition before the registry of depression diagnosis in the database. | |
| **14** | Avoid BZD drugs long term use (>4 weeks) | Risk of dependency; daytime sedation; cognitive impairment; agitation; irritability | PIP was defined as (i) ≥1 prescription of BZD with total length >30 days, allowing for a gap between prescriptions of ≤7 days in patients with complete dosage information, or (ii) minimal cumulative dose prescription (see below) of a BZD drug within the aforementioned period whenever dosage information was missing:<br><br>Diazepam all formulations: 28 days *10 mg (prescription of >280 mg)<br>Clonazepam all formulations: 28 days *8 mg (prescription of >224 mg)<br>Temazepam all formulations: 28 days *20 mg (prescription of >560 mg)<br>Chlordiazepoxide all formulations: 28 days *30 mg (prescription of >840 mg)<br>Lorazepam 1 mg tablets: 28 days *2.5 mg (prescription of >70 mg)<br>Loprazolam 1 mg tablets: 28 days *1 mg (prescription of >28 mg)<br>Oxazepam all formulations: 28 days* 50 mg (prescription of >1,400 mg)<br>Nitrazepam 5 mg tablets: 28 days * 5 mg (prescription of >140 mg)<br>Clobazam 10 mg tablets: 28 days* 20 mg (prescription of >560 mg)<br>Alprazolam 250 mcg tablets: 28 days * 1 mg (prescription of >28 mg) | Potential PIP overestimation. By using the minimal cumulative dose prescription approach, we may have misclassified patients receiving high doses of BZD for <4 weeks as receiving PIP. Moreover, we defined drug use >4 weeks as the prescription of >28 days of the drug, considering patients received a pack size for 28 days of treatment. Thus, patients receiving a prescription for 4 weeks with a pack size of 30 days may be misclassified as PIP. | Fully applied |
| **15** | Avoid non-BZD hypnotics (zolpidem, zaleplon and zopiclone) use >4 weeks | Non-BZD hypnotics have ADR similar to those of BZD (e.g., daytime sedation, cognitive impairment, agitation, and irritability) | PIP was defined as (i) ≥1 prescription of non-BZD drugs with total length >30 days, allowing for a gap between prescriptions of ≤7 days in patients with complete dosage information, or (ii) minimal cumulative dose prescription (see below) of a non-BZD drug within the aforementioned period whenever dosage information was missing:<br>Zopiclone all formulations: 28 days * 7.5 mg (prescription of >210 mg)<br>Zolpidem all formulations: 28 days * 10 mg (prescription of >280 mg) | Potential PIP underestimation. By using the minimal cumulative dose prescription approach, we may have misclassified patients receiving high doses of non-BZD hypnotics for <4 weeks as receiving PIP. Moreover, we defined drug use >4 weeks as the prescription of >28 days of the drug, considering patients received a pack size for 28 days of treatment. Thus, patients receiving a prescription for 4 weeks with a pack size of 30 days may be misclassified as PIP. | Fully applied |
| **16** | Avoid carbamazepine in combination with clarithromycin or erythromycin | Inhibition of carbamazepine metabolism; headache; drowsiness; nausea; | PIP was defined as the concomitant use of carbamazepine and clarithromycin or erythromycin. Concomitant use was defined as ≥1 day of prescription overlap between the two drugs. | PIP potential overestimation as we assumed that patients with prescriptions overlapping ≥1 day received the two drugs concomitantly (we have not considered that physicians could have advised patients to stop one drug before initiating another). | Fully applied |

**Table S2.** Continued.

| | | | | | |
|---|---|---|---|---|---|
| **17** | Strong opioids should not be prescribed without laxatives | Constipation | PIP was defined as ≥1 prescription of strong opioids at index date or previous 90 days, with no prescription for laxatives within the 90 days period. | Potential overestimation of PIP. Laxatives are OTC drugs; thus, patients may have used laxatives without a prescription. | Fully applied |
| **18** | Nitrofurantoin should not be prescribed >7 days for uncomplicated UTI | Potential for pulmonary toxicity | PIP was defined ≥1 prescription of nitrofurantoin with total length >7 days, allowing for gap of ≤1 day between prescriptions for patients with information on dosage, or (ii) minimal cumulative dose prescription of >1,400 mg of nitrofurantoin (7 days * 200 mg) within the aforementioned period whenever dosage information was missing | Potential PIP overestimation as we consider that nitrofurantoin was only indicated for the management of UTI. | Fully applied |
| **19** | Avoid use of long acting SU for management of diabetes | Oral long-acting SU can cause prolonged hypoglycaemia or syndrome of inappropriate ADH secretion; | PIP was defined as ≥1 prescription of long acting SU at index date or previous 90 days. | | Fully applied |
| **20** | Avoid long-term NSAID treatment | Long-term NSAID treatment should be reviewed periodically due to increased risk of thrombotic effects, and the lowest effective dose should be prescribed for the shortest period. | PIP was defined as (i) ≥1 prescription NSAID with total length >90 days at index date or previous 90 days, allowing a gap of ≤7 days between prescriptions for patients with information on dosage, or (ii) minimal cumulative dose prescription (see below) of a NSAID within the aforementioned period whenever dosage information was missing:<br>Ibuprofen all formulations: 90 days * 1,2 g (prescription of >108 g)<br>Naproxen all formulations: 90 days * 500 mg (prescription of >45,000 mg)<br>Etodolac 600 mg: 90 days * 400 mg (prescription of >36,000 mg)<br>Meloxican all formulations: 90 days * 15 mg (prescription of >1,350 mg)<br>Diclofenac all formulations: 90 days * 100 mg (prescription of >9,000 mg)<br>Etoricoxib all formulations: 90 days * 60 mg (prescription of >5,400 mg)<br>Mefenamic acid all formulations: 90 days * 1 g (prescription of >90 g)<br>Celecoxib all formulations: 90 days * 200 mg (prescription of >18,000 mg)<br>Aceclofenac 100mg tablets: 90 days * 200 mg (prescription of >18,000 mg)<br>Nabumetone 500mg tablets: 90 days * 1 g (prescription of >90 g)<br>Indometacin 50mg capsules: 90 days * 100 mg (prescription of >9,000 mg)<br>Piroxicam all formulations 90 days * 20 mg (prescription of >1,800 mg) | Potential PIP underestimation. By using the minimal cumulative dose prescription approach, we may have misclassified patients receiving high doses of non-BZD hypnotics for <4 weeks as receiving PIP. Moreover, we may have underestimated the use of NSAID (over-the counter drugs). | Fully applied |
| **21** | Avoid combination of NSAID with low dose aspirin or SSRI, without adequate GI protection | Risk of gastro-intestinal bleeding | PIP was defined as ≥1 prescription of NSAID and ≥1 prescription of SSRI or aspirin, with ≥1-day prescription overlap, at or in the prior to 90-days, with no additional prescription of PPI or H2-receptor antagonist within the 90-days period. | Potential underestimation of PIP. We assumed patients with a prescription for PPI or H2-receptors within 90 days had the gastroprotective medication while using the combination of NSAID + SSRI or aspirin. | Fully applied |

**Table S2.** Continued.

| | | | | | |
|---|---|---|---|---|---|
| **22** | Avoid using 2+ drugs from the same pharmacological class, unless used for additive effects in line with current clinical guidelines – Example 1: duplication of opioids (excluding methadone and morphine) | Possible unwanted duplication of effect, increasing risk of side effects and adverse events | PIP was defined as ≥1 prescription of two different opioids (i.e., different ATC codes at 5$^{th}$ level) at index date or within 90 days previous to index date, with ≥1 day overlap between prescriptions. | We assumed that patients with prescriptions overlapping at least one day were using the two drugs concomitantly. We have not considered that physicians could have advised patients to stop one drug before initiating another). Also, potential underestimation of PIP, as we considered only patients with information on dosage for at least one drug in the analysis. | Partially applied |
| **22** | Avoid using 2+ drugs from the same pharmacological class, unless used for additive effects in line with current clinical guidelines – Example 2: duplication of SSRI | Possible unwanted duplication of effect, increasing risk of side effects and adverse events | PIP was defined by ≥1 prescription of two different SSRI (i.e., different ATC codes at 5$^{th}$ level) at index date or within 90 days previous to index date, with ≥1 day overlap between prescriptions. | Potential overestimation of PIP as the co-prescription of two SSRI may be due to medication transition. We have not considered that physicians could have advised patients to stop one drug before initiating another). Also, potential underestimation of PIP, as we considered only patients with information on dosage for at least one drug in the analysis. | Partially applied |
| **22** | Avoid using 2+ drugs from the same pharmacological class, unless used for additive effects in line with current clinical guidelines – Example 3: duplication of PPI | Possible unwanted duplication of effect, increasing risk of side effects and adverse events | PIP was defined by ≥1 prescription of two different PPI (i.e., different ATC codes at 5$^{th}$ level) at index date or within 90 days previous to index date, with ≥1 day overlap between prescriptions. | We have not considered that physicians could have advised patients to stop one drug before initiating another). Also, potential underestimation of PIP, as we considered only patients with information on dosage for at least one drug in the analysis. | Partially applied |

**Abbreviations:** AAB: alpha-adrenoceptor blocking drugs; ADH: antidiuretic hormone; ADR: adverse drug reaction; AH: antihistamine; ATC: Anatomical therapeutic chemical classification ; BB: beta blockers; BZD: benzodiazepine; CCB: calcium channel blockers; COPD: chronic obstructive pulmonary disease; GP: General practitioner; NSAID: non-steroidal anti-inflammatory drugs; PIP: Potentially inappropriate prescription; PPI: proton pump inhibitors; SSRI: selective serotonin reuptake inhibitor; SU: sulfonylureas; TCA: Tricyclic antidepressant; UTI: Urinary tract infections.

**Table S3.** List of Anatomical therapeutic chemical (ATC) classification codes and Read codes used to assess potential inappropriate prescription (PIP) according to the 2015 American Geriatrics Society Beers criteria.

| Table Number[1] | Criteria Number | ATC codes used | Read codes used |
|---|---|---|---|
| **2** | 1 | **First-generation anti-histamines**: "R06AB01" "R05X" "R01BA52" "R01BA51" "R06AB51" "R06AB04" "R06AB54" "R06AA54" "R06AA04" "R06AX02" "N07CA52" "R06AA02" "N02BE71" "R06AA52" "N07XX59" "R01BA02" "R05DA20" "R05FB02" "R01BA52" "R06AA09" "R05CA03" "R01AA04" "N05B B01" "R06AE05" "R06AE55" "R06AD02" "N02AB52" "R06AX07" | **Severe allergy:** "2227.11" "3359.00" "3368.00" "66G..00" "66G3.00" "66GZ.00" "7Q08400" "8Hld.00" "8HVK000" "8T0C.00" "9b99.00" "9b9W.00" "9NlX.00" "AB63300" "D310.00" "D310011" "D310z00" "D403300" "F401400" "F4C0611" "F4D3100" "F4D3111" "M280.00" "SN50.11" "SN50000" "SN50100" "SN59000" "SN59100" "SN59200" "SN59300" "SN59400" "SN5A.00" "ZL18200" "ZL5A600" "ZL9A200" "ZLD3200" "ZLE6100" |
| **2** | 2 | **Antiparkinsonian agents:** "N04AC01" "N04AA01" "N04AB02" "M03BA03"<br>**Antipsychotics:** "N05A" "N06C" "A03CA01" "N01AH51" | |
| **2** | 3 | **Antispasmodics (exclude ophthalmic):** "A03BA01" "A04A D04" "A03B A01" "G04BE02 in combination with atropine" "A03CA02" "A03AA07" "A02AA02" "A03BB01" "A04AD01" "A03AB05" | |
| **2** | 4 | **Dipyridamole (oral short-acting):** "B01AC07" | |
| **2** | 5 | **Triclopidine:** "B01AC05" | |
| **2** | 6 | **Nitrofurantoin:** "J01XE01" | **Chronic Kidney disease stages 3B, 4 and 5:** "1Z13.00" "1Z14.00" "1Z16.00" "1Z1F.00" "1Z1G.00" "1Z1H.00" "1Z1J.00" "1Z1K.00" "1Z1L.00" "K054.00" "K055.00" |
| **2** | 7 | **Alpha-1 blockers (doxazosin, prazosin, and terazosin):** "C02CA04" "C02CA01" "G04CA03" | **Hypertension:** "14A2.00" "661M600" "661N600" "662..12" "6627.00" "6628.00" "662b.00" "662c.00" "662d.00" "662F.00" "662G.00" "662O.00" "662P.00" "662P000" "662P100" "662q.00" "8B26.00" "8BL0.00" "8CR4.00" "8HT5.00" "8I3N.00" "8IA6.00" "9OI1.00" "9OI3.00" "9OI4.00" "9OI5.00" "9OI6.00" "9OI7.00" "9OI8.00" "9OIA.11" "9OIZ.00" "G2...00" "G2...11" "G20..00" "G20..12" "G200.00" "G201.00" "G202.00" "G203.00" "G20z.00" "G20z.11" "G21..00" "G210.00" "G210000" "G210100" "G210z00" "G211.00" "G211000" "G211100" "G211z00" "G21z.00" "G21z000" "G21z011" "G21z100" "G21zz00" "G22..00" "G220.00" "G221.00" "G222.00" "G22z.00" "G22z.11" "G23..00" "G230.00" "G231.00" "G232.00" "G233.00" "G234.00" "G23z.00" "G24..00" "G240.00" "G240000" "G240z00" "G241.00" "G241000" "G241z00" "G244.00" "G24z.00" "G24z000" "G24z100" "G24zz00" "G25..00" "G25..11" "G250.00" "G251.00" "G26..00" "G26..11" "G27..00" "G28..00" "G2y..00" "G2z..00" "G672.11" "G8y3.00" "Gyu2.00" "Gyu2000" "Gyu2100" "L12..00" "L122.00" "L122000" "L122100" "L122300" "L122400" "L122z00" "L127.00" "L127000" "L127100" "L127200" "L127300" "L127400" "L127z00" "L128.00" "L128000" "L128100" "L128200" "L12B.00" "Lyu1.00" |
| **2** | 8 | **Clonidine, guanabenz, guanfacine, methyldopa, and reserpine:** "C02AC02" "C02AB01" "C02LB01" "C02AA02" "C03AA01 (whenever the formulation combines reserpine)" "C03AA03 (whenever the formulation combines reserpine)" "R03DA07 (whenever the formulation combines reserpine)" "N02CX02" "C02AC01"<br>**Other antihypertensive drugs:** "C02 except C02CA" "C09AA" "C09C" "C09D" "C08C" "C08D" "C03A" "C03B" "C03C" "C03D" "C03E" "C03XA" "C07" "C09XA02" "C02CA" | **Hypertension**: see criteria number 7. |

**Table S3.** Continued.

| | | | |
|---|---|---|---|
| **2** | 9 | **Disopyramide:** "C01BA03" | |
| **2** | 10 | **Dronedarone:** "C01BD07" | **Persistent atrial fibrillation:** "G573500" "G573400"<br>**Decompensated heart failure:** "G580200" |
| **2** | 11 | **Digoxin:** "C01AA05"<br>**Beta blockers:** "C07" (except C07AA07)<br>**Calcium-channel blockers:** "C08DB01" "C08DA01" "C08DA51" "C09BB10""C08DB01" | **Atrial fibrillation:** "14AD.00" "14AN.00" "2JS..00" "3272" "3283" "662S.00" "6A9..00" "9Os..00" "9Os0.00" "9Os1.00" "9Os2.00" "9Os3.00" "9Os4.00" "G573.00" "G573000" "G573200" "G573300" "G573400" "G573500" "G573700" "G573z00" "G574.00" "G574000" "G574011" "G574z00" |
| **2** | 12 | **Digoxin:** see criteria number 11<br>**Beta blockers:** "C07AB07" "C07AB12" "C07AG02"<br>**Calcium-channel blockers:** see criteria 11<br>**Angiotensin receptor blockers:** "C09C" "C09D"<br>**Angiotensin-converting enzyme inhibitors:** "C09AA" "C09B"<br>**Mineralocorticoid receptor antagonists:** "H02AA02" | **Heart failure:** "14A6.00" "14AM.00" "1O1..00" "2JZ..00" "661M500" "661N500" "662p.00" "662T.00" "662W.00" "679W100" "679X.00" "67D4.00" "8B29.00" "8CL3.00" "8CMK.00" "8H2S.00" "8HBE.00" "8HHz.00" "8Hk0.00" "8HTL000" "8IE0.00" "8IE1.00" "9Or..00" "9Or0.00" "9Or1.00" "9Or2.00" "9Or3.00" "9Or4.00" "9Or5.00" "G232.00" "G58..00" "G58..11" "G580.00" "G580.11" "G580.12" "G580000" "G580100" "G580200" "G580300" "G580400" "G582.00" "G583.00" "G583.11" "G583.12" "G58z.00" "G58z.12" "G5y4z00" "L09y200" "Q48y100" "SP11100" "SP11111" |
| **2** | 13 | **Digoxin:** see criteria number 11 | **Atrial fibrillation:** see criteria number 11<br>**Heart failure:** see criteria number 12 |
| **2** | 14 | **Nifedipine:** "C08CA05" | |
| **2** | 15 | **Amiodarone:** "C01BD01"<br>**Other antiarrhythmic drugs:** "C07" (except C07AA07) "C08DB01" "C08DA01" "C08DA51" "C09BB10" "C08DB01" | **Atrial fibrillation:** see criteria number 11<br>**Heart failure:** see criteria number 12<br>**Left ventricular hypertrophy:** "G5y3411" |
| **2** | 16 | **Amitriptyline, amoxapine, clomipramine, desipramine, doxepin (>6mg/day), imipramine, nortriptyline, paroxetine, protriptyline, trimipramine:** "N06AA09" "N06CA01" "N06AA17" "N06AA04" "N06AA01" "N06AA12" "N06AA02" "N06AA10" "N06AB05" "N06AA11" "N06AA06" | |
| **2** | 17 | **Antipsychotics:** "N05AA" "N06CA01" "N05AB03" "N05AB04" "N05AB06" "N05AB02" "A03CA01" "N05AC02" "N05AC04" "N05AD01" "N05AD02" "N05AD07" "N05AD08" "N01AH51 whenever formulation combined with antipsychotics" "N05AE01" "N05AF01" "N05AF03" "N05AF05" "N05AG02" "N05AH" "N05AL01" "N05AL04" "N05AL05" "N05A X08" "N05AB02" "N05AE05" "N05AE03" "N05AG01" "N05AN01" "N05AX11" "N05AX13" | **Schizophrenia:** "13L3.12" "13Y2.00" "1464.00" "212T.00" "212W.00" "E10..00" "E100.00" "E100.11" "E100000" "E100100" "E100200" "E100300" "E100400" "E100500" "E100z00" "E101.00" "E101000" "E101100" "E101200" "E101300" "E101400" "E101500" "E101z00" "E102.00" "E102000" "E102100" "E102200" "E102300" "E102400" "E102500" "E102z00" "E103.00" "E103000" "E103100" "E103200" "E103300" "E103400" "E103500" "E103z00" "E104.00" "E105.00" "E105000" "E105100" "E105200" "E105300" "E105400" "E105500" "E105z00" "E106.00" "E106.11" "E107.00" "E107.11" "E107000" "E107100" "E107200" "E107300" "E107400" "E107500" "E107z00" "E10y.00" "E10y.11" "E10y000" "E10y100" "E10yz00" "E10z.00" "E122.00" "E14z.11" "E212.00" "E212000" "E212200" "E212z00" "Eu05200" "Eu05212" "Eu2..00" "Eu20.00" "Eu20000" "Eu20011" "Eu20100" "Eu20111" "Eu20200" "Eu20212" "Eu20213" "Eu20214" "Eu20300" "Eu20311" "Eu20400" "Eu20500" "Eu20511" "Eu20512" "Eu20600" "Eu20y00" "Eu20y11" "Eu20y12" "Eu20y13" "Eu20z00" "Eu21.00" "Eu21.11" "Eu21.12" "Eu21.13" "Eu21.14" "Eu21.15" "Eu21.16" "Eu21.17" "Eu21.18" "Eu22013" "Eu23000" "Eu23100" "Eu23111" "Eu23112" "Eu23200" "Eu23211" "Eu23212" "Eu23214" "Eu25.00" "Eu25000" "Eu25011" "Eu25012" "Eu25100" "Eu25111" "Eu25112" "Eu25200" "Eu25211" "Eu25212" "Eu25y00" "Eu25z00" "Eu25z11" "Eu60100" "Eu84512" "ZRhi.00" "ZS7C611" "ZV11000" (cont.) |

**Table S3.** Continued.

| | | | |
|---|---|---|---|
| **2** | 17 | | (cont.)<br>**Bipolar disorder:** "E11..11" "E114.00" "E114000" "E114100" "E114200" "E114300" "E114400" "E114500" "E114600" "E114z00" "E115.00" "E115000" "E115100" "E115200" "E115300" "E115400" "E115500" "E115600" "E115z00" "E116.00" "E116000" "E116100" "E116200" "E116300" "E116400" "E116500" "E116600" "E116z00" "E117.00" "E117000" "E117100" "E117200" "E117300" "E117400" "E117500" "E117600" "E117z00" "Eu30.11" "Eu31.00" "Eu31000" "Eu31100" "Eu31200" "Eu31300" "Eu31500" "Eu31600" "Eu31700" "Eu31800" "Eu31900" "Eu31911" "Eu31y00" "Eu31y11" "Eu31z00" "ZRby100" |
| **2** | 18 | **Barbiturates:** "N05CB01" "N05CA02" "N03AA01" "N03AA02" "R03DA74" "N05CA06" | |
| **2** | 19 | **Short- and intermediate-acting benzodiazepines:** "N05BA12" "N05BA06" "N05BA04" "N05CD07" "N05CD05" "N05BA09" "N05BA10" "N05BA03" | |
| **2** | 20 | **Long-acting benzodiazepines:** "N05BA05" "N05BA02" "N06CA01" "A03CA02" "N03AE01" "N05BA01" "N05CD01" "N05CD02" "N05CD06" "N05BA08" | |
| **2** | 21 | **Meprobamate:** "N05BC01" "N02BA71" "C03AA01 whenever formulation combined with meprobamate" | |
| **2** | 22 | **BZD-receptor agonist hypnotics**: "N05CF01" "N05CF02" "N05CF03" | |
| **2** | 23 | **Ergoloid mesylates (dehydrogenated ergot alkaloids), Isoxsuprine:** "C04AA01" "C04AE01" | |
| **2** | 24 | **Androgens:** "G03BA02" "G03BA03" | **Hypogonadism:** "C139.00" "C163.11" "C163200" "C163300" "C172.11" "C172.12" "F146.00" |
| **2** | 25 | **Desiccated thyroid:** "H03AA05" | |
| **2** | 26 | **Estrogens:** "G03FA14" "G03FA11" "G03CA03" "G03AA14" "G03FA17" "G03FA01" "G03FB06" "G03FB08" "G03FB05" "G03FB09" "G03FB01" "G03CA53" "G03AA11" "G03AA09" "G03AA10" "G03AA12" "G03AA07" "G03AB04" "G03AB06" "G03CA01" "G03AA05" "G03AA01" "G03DC02" "G03BA02" "G03CA04" "L02A A01" "G03C X01" | **Transphenoidal hypophysectomy**: 7100300 |
| **2** | 27 | **Somatotropin:** "H01AC01 | |
| **2** | 28 | **Megestrol:** "L02AB01" | |
| **2** | 29 | **Chlorpropamide:** "A10BB02" | |
| **2** | 30 | **Glyburide / Glibenclamide:** "A10BB01" | |
| **2** | 31 | **Metoclopramide:** "A03FA01" "N02BE51 whenever formulation combined with metoclopramide" | **Gastroparesis:** "C10FR00" "C10FR11" "J16y900" |
| **2** | 32 | **Mineral oil:** "A06AA01" "A06AA51" | |
| **2** | 33 | **Proton pump inhibitors:** "A02BC" "M01AE53" "A02BD" "M01AE52"<br>**Oral corticosteroids:** "H02AB01" "H02AB02" "H02AB04" "H02AB06" "H02AB07" "H02AB08" "H02AB09" "A01AC03" "H02AB10" "H02AB13" "A07EA07" "A07EA06"<br>**Non-steroidal anti-inflammatory drugs:** "M01A" (excluding topical formulations, suppositories, eye drops, and plasters)<br>**H2-receptor antagonist:** "A02BA" | |

**Table S3.** Continued.

| | | | |
|---|---|---|---|
| **2** | 34 | **Meperidine:** "N02AB02" | |
| **2** | 35 | **Non-cyclooxygenase-selective non-steroidal anti-inflammatory drugs:** "N02BA01" "N02AJ07" "N02BA51" "N06BC01" "M01AB05" "M01AB55" "N02BA11" "M01AB08" "M01AE04" "M01AE51" "M01AE01" "N02BE51 whenever in combination with ibuprofen" "N02AJ08" "N02BE71 whenever in combination with ibuprofen" "M01AE03" "M01AE53" "M01AG01" "M01AC06" "M01AX01" "M01AE02" "M01AE52" "M01AC01" "M01AB02" "M01AB03"<br>**Proton pump inhibitor:** "A02BC" "M01AE53" "A02BD" "M01AE52"<br>**Misoprostol:** "A02BB01" | |
| **2** | 36 | **Indomethacin, ketorolac:** "M01AB01" "M01AB15" | |
| **2** | 37 | **Pentazocine:** "N02AD01 " | |
| **2** | 38 | **Skeletal muscle relaxants:** "M03BA02" "M03BA03" "M03BA53" "N04AB02" "M03BC01" " M03BC51" | |
| **2** | 39 | **Desmopressin:** "H01BA02" | **Nocturia or nocturnal polyuria:** "1A13.00" "1A22011" "R084200" |
| **3** | 40 | **Non-steroidal anti-inflammatory drugs:** M01A" (excluding M01AH, topical formulations, suppositories, eye drops, and plasters)<br>**COX-2 inhibitors:** "L01XX33" "M01AH01" "M01AH02" "M01AH03" "M01AH04" "M01AH05" "M01AH06"<br>**Thiazides**: "A10BG03" "A10BD06" "A10BD03" "A10BG02"<br>**Cilostozol:** "B01AC23"<br>**Dronedarone:** "C01BD07"<br>**Nondihydropyridine CCBs:** "C08DB01" "C08DA01" "C09BB10" | **Heart failure:** see criteria 12<br>**Severe/ decompensated heart failure**: "G580200" |
| **3** | 41 | **Alpha-1 blockers (doxazosin, prazosin, and terazosin):** "C02CA04" "C02CA01" "G04CA03"<br>**Acetylcholinesterase inhibitors:** "N06DA" "N07AA"<br>**Tertiary tricyclic antidepressant drugs:** "N06AA09" "N06CA01" "N06AA02" "N06AA06" "N06AA12" "N06AA04"<br>**Other antipsychotics (chlorpromazine, thioridazine, olanzapine):** "N05AA01" "N05AC02" "N05AH03" | **Syncope:** "147A.00" "147B.00" "1B6..11" "1B6..12" "1B62.00" "1B68.00" "2244.00" "Eu46y16" "G33z200" "R002.00" "R002.11" "R002100" "R002400" "R002500" "R002600" "R002z00" "R062000" "SN21.00" "SN21.12" |
| **3** | 42 | **Bupropion, chlorpromazine, clozapine, mapritiline, olanzapine, thioridazine, thiothixene, and tramadol:** "N06AX12" "N05AA01" "N05AH02" "N06AA21" "N05AH03" "N05AC02" "N02AX02" "N02AJ13" "N02BE71" | **Seizures:** "1473.00" "14On.00" "1B26.00" "1B27.00" "1O30.00" "282..13" "6110.00" "667..00" "6671.00" "6672.00" "6674.00" "6677.00" "6678.00" "6679.00" "667A.00" "667B.00" "667C.00" "667D.00" "667E.00" "667F.00" "667G.00" "667H.00" "667J.00" "667K.00" "667L.00" "667M.00" "667N.00" "667Q.00" "667R.00" "667S.00" "667T.00" "667V.00" "667W.00" "667Z.00" "67AF.00" "67IJ000" "8BIF.00" "8CE7.00" "8Hlp.00" "8IAg.00" "8IAh.00" "8IAi.00" "8IB2.00" "8IB3.00" "8IB4.00" "8T0L.00" "9N0r.00" "9N4V.00" "9Of..00" "9Of0.00" "9Of1.00" "9Of2.00" "9Of3.00" "9Of4.00" "9Of5.00" "9Of6.00" "9Of7.00" "E201500" "Eu05212" "Eu06013" "Eu10800" "Eu44511" "Eu80300" "F132100" "F132z12" "F25..00" "F250.00" "F250000" "F250100" "F250200" "F250300" "F250400" "F250y00" (cont.) |

**Table S3.** Continued.

| | | | |
|---|---|---|---|
| **3** | 42 | | (cont.) "F250z00" "F251.00" "F251000" "F251011" "F251100" "F251200" "F251300" "F251400" "F251500" "F251600" "F251y00" "F251z00" "F254.00" "F254000" "F254100" "F254200" "F254300" "F254500" "F254z00" "F255.00" "F255000" "F255011" "F255012" "F255100" "F255200" "F255300" "F255311" "F255400" "F255500" "F255600" "F255y00" "F255z00" "F257.00" "F25A.00" "F25B.00" "F25C.00" "F25D.00" "F25E.00" "F25F.00" "F25G.00" "F25H.00" "F25y.00" "F25y000" "F25y100" "F25y400" "F25yz00" "F25z.00" "Fyu5000" "Fyu5100" "Q480.12" "R003300" "R003400" "R003z11" "SC20000" |
| **3** | 43 | **Anticholinergics:** "R06AB01" "R06AA52" "R06AD52" "R01BA52" "R01BA51" "R06AB51" "R06AB04" "R06AA54" "R06AA04" "R06AX02" "N07CA52" "R06AA02" "N02BE71" "N07XX59" "R01BA02" "R05FB02" "R06AA09" "R05CA03" "R01AA04" "N05BB01" "R06AX07" "N04AA01" "N04AB02" "M03B C01" "N02B E01" "N06A A09" "N06CA01" "N06AA17" "N06AA04" "N06AA01" "N06AA12" "N06AA02" "N06AA10" "N06AB05" "N06AA11" "N06AA06" "N05AA01" "N05AH02" "N05AH01" "N05AH03" "N05AB03" "N05AC02" "N05AB06" "A03CB04" "C01BA03" "G04BD10" "G04BD11" "G04BD02" "G04BD04" "G04BD08" "G04CA53" "G04BD07" "G04BD09" "A07DA52" "A03CB02" "A03CB" "A06AB04" "A03BA04" "C02AA03" "A03CA02" "A03BA01" "A07DA01" "A04AD04" "A03BA01" "N02AA51" "G04BE02" "A03AB05" "N05AB04" "R06AD02" " "N02AB52"<br>**Antipsychotics (incl. chlorpromazine):** "N05AA" "N06CA01" "N05AB03" "N05AB04" "N05AB06" "N05AB02" "A03CA01" "N05AC02" "N05AC04" "N05AD01" "N05AD02" "N05AD07" "N05AD08" "N01AH51 whenever formulation combined with antipsychotics" "N05AE01" "N05AF01" "N05AF03" "N05AF05" "N05AG02" "N05AH" "N05AL01" "N05AL04" "N05AL05" "N05A X08" "N05AB02" "N05AE05" "N05AE03" "N05AG01" "N05AN01" "N05AX11" "N05AX13"<br>**Benzodiazepines:** "N05BA12" "N05CD11" "N05BA06" "N05CD06" "N05BA04" "N05CD07" "N05CD05" "N05BA08" "N05BA09" "N05BA05" "N05BA02" "N06CA01" "A03CA02" "N03AE01" "N05BA01" "N05CD03" "N05CD01" "N05CD02" "N05BA11" "N05CD08" "N05BA03" "N05BA10"<br>**Corticosteroids:** "H02AB01" "H02AB02" "H02AB04" "H02AB06" "H02AB07" "H02AB08" "H02AB09" "A01AC03" "H02AB10" "H02AB13" "A07EA07" "H02BX01"<br>**H2-receptor blockers:** "A02BA01" "A02BA51" "A02BA02" "A02BA07" "A02BA53" "A02BA03" "A02BA04"<br>**Meperidine:** "N02AB02"<br>**Sedative hypnotics:** "N05CF01" "N05CF02" "N05CF03" "N05CB01" "N05CA02" "N05CA03" "N03AA01" "N03AA02" " C02LA71" "N05CA06" "N05CC01" "N05CE05" "N05CM02" | **Delirium:** "E001100" "E003.00" "E004100" "E010.00" "E010.11" "E010.12" "E02y000" "E030.11" "E031.11" "E133.11" "Eu04.00" "Eu04000" "Eu04100" "Eu04y00" "Eu04y11" "Eu04z00" "Eu10411" "Eu23011" "Eu23111"<br>**Dementia:** "1461" "66h..00" "6AB..00" "8BM0200" "8BPa.00" "8CET.00" "8CMe000" "8CMG200" "8CMZ.00" "8CMZ000" "8CMZ100" "8CMZ300" "8CSA.00" "8IAe000" "8IAe200" "9Ou..00" "9Ou1.00" "9Ou2.00" "9Ou3.00" "9Ou4.00" "9Ou5.00" "E00..11" "E00..12" "E000.00" "E001.00" "E001000" "E001100" "E001200" "E001300" "E001z00" "E002.00" "E002000" "E002100" "E002z00" "E003.00" "E004.00" "E004.11" "E004000" "E004100" "E004200" "E004300" "E004z00" "E012.00" "E012.11" "E02y100" "E041.00" "Eu00.00" "Eu00000" "Eu00011" "Eu00012" "Eu00013" "Eu00100" "Eu00111" "Eu00112" "Eu00113" "Eu00200" "Eu00z00" "Eu00z11" "Eu01.00" "Eu01.11" "Eu01000" "Eu01100" "Eu01111" "Eu01200" "Eu01300" "Eu01y00" "Eu01z00" "Eu02.00" "Eu02000" "Eu02100" "Eu02200" "Eu02300" "Eu02400" "Eu02500" "Eu02y00" "Eu02z00" "Eu02z11" "Eu02z13" "Eu02z14" "Eu02z16" "Eu04000" "Eu04100" "Eu10711" "Eu84311" "F110.00" "F110000" "F110100" "F118100" "Fyu3000" "ZR1T.00" "ZS7C500" "E00z.00" "Eu02z12" "Eu02z15" "F111.00" "F112.00" |
| **3** | 44 | **Anticholinergics:** see criteria 43<br>(cont.) | **Dementia and cognitive impairment:** "1461" "66h..00" "6AB..00" "8BM0200" "8BPa.00" "8CET.00" "8CMe000" "8CMG200" "8CMZ.00" "8CMZ000" "8CMZ100" "8CMZ300" "8CSA.00" "8IAe000" "8IAe200" "9Ou..00" "9Ou1.00" "9Ou2.00" "9Ou3.00" "9Ou4.00" "9Ou5.00" "E00..11" "E00..12"(cont.) |

**Table S3.** Continued.

| | | | |
|---|---|---|---|
| **3** | 44 | (cont.)<br>**Benzodiazepines:** see criteria 43<br>**H2-receptor blockers:** see criteria 43<br>**Non-benzodiazepine, benzodiazepine receptor agonist hypnotics:** "N05CF01" "N05CF02" "N05CF03" | (cont.) "E000.00" "E001.00" "E001000" "E001100" "E001200" "E001300" "E001z00" "E002.00" "E002000" "E002100" "E002z00" "E003.00" "E004.00" "E004.11" "E004000" "E004100" "E004200" "E004300" "E004z00" "E012.00" "E012.11" "E02y100" "E041.00" "Eu00.00" "Eu00000" "Eu00011" "Eu00012" "Eu00013" "Eu00100" "Eu00111" "Eu00112" "Eu00113" "Eu00200" "Eu00z00" "Eu00z11" "Eu01.00" "Eu01.11" "Eu01000" "Eu01100" "Eu01111" "Eu01200" "Eu01300" "Eu01y00" "Eu01z00" "Eu02.00" "Eu02000" "Eu02100" "Eu02200" "Eu02300" "Eu02400" "Eu02500" "Eu02y00" "Eu02z00" "Eu02z11" "Eu02z13" "Eu02z14" "Eu02z16" "Eu04000" "Eu04100" "Eu10711" "Eu84311" "F110.00" "F110000" "F110100" "F118100" "Fyu3000" "ZR1T.00" "ZS7C500" "E00z.00" "Eu02z12" "Eu02z15" "F111.00" "F112.00" "28E..00" "28E0.00" "28E1.00" "28E2.00" "28E3.00" "311B.00" "38Dv.00" "38Dv000" "38Qr.00" "38Qv.00" "38Qv.11" "3AD3.00" "3AE1.00" "3AE2.00" "3AE3.00" "3AE4.00" "3AE5.00" "3AE6.00" "3AF..00" "6AQ..00" "9OqG.11" "Eu05700" "Eu05800" "Z7C..00" "Z7C1.00" "Z7C2.00" "ZR1H.00" "ZR1I.00" "ZR3b.00" "ZR3b.11" "ZRa2.00" "ZRd..00" "ZRh6.00" "ZRh6.11" "ZRkL.00" "ZRLfE00" "ZRLfF00" "ZRV9.11" "ZRVa.00" "ZRVa.11" "ZRVt.00" "ZRVt.11" "ZS3..00" |
| **3** | 45 | **Benzodiazepines:** see criteria 43<br>**Non-benzodiazepine, benzodiazepine receptor agonist hypnotics:** see criteria 44<br>**Antipsychotics:** see criteria 43<br>**Anticonvulsants:** "N05BA01" "N03A" "R03DA07 whenever combined with phenobarbital"<br>**Tricyclic antidepressants:** "N06AA09" "N06CA01" "N06AA02" "N06AA06" "N06AA12" "N06AA07" "N06AA17" "N06AA01" "N06AA10" "N06C" "N06AA11" "N06AA04" "N06AA13" "N06AA15" "N06AA16" "N06AA21"<br>**Selective serotonin reuptake inhibitors:** "N06AB"<br>**Opioids:** "N02AA01" "N02AA51" "N01AB02" "N02AA02" "N02AA03" "N02AA05" "N02AA55" "N02AJ" "N02AA08" "N02AA10" "N02AB02" "N02AB52" "N01AH" "N02AB03" "N02AD01" "N02BE01" "N02AC54" "N02AC01" "N02AC04" "N02AD02" "N07BC01" "N02AE01" "N07BC51" "N02AF02" "N02AX02" "N02AX" "R05FA02" "A07DA52" "A07DA02" "R06AA09 whenever in combination with opioids" "N02AA59" "N02AB52" "N07BC02" "N02AC52" | **Falls:** "16D..00" "16D1.00" "16D5.00" "16D6.00" "1Bb0.00" "38A..00" "38A0.00" "38A1.00" "66aF.00" "67IC.00" "67ID.00" "67IE.00" "8BIG.00" "8CM7.00" "8CMW400" "8Hk1.00" "8HTl.00" "R01z600" "R200.12" "TCy..00" "U10..00"<br>**Fractures:** "14G6.00" "14G7.00" "14G8.00" "14G9.00" "14GA.00" "7J41.00" "7J41000" "7J41100" "7J41200" "7J41300" "7J41400" "7J41500" "7J41y00" "7J41z00" "7J42300" "7J42400" "7J42500" "7J42600" "7J42700" "7J42800" "7J42900" "7J42A00" "7J42a00" "7J42b00" "7J42B00" "7J42c00" "7J42C00" "7J42d00" "7J42D00" "7J42e00" "7J42E00" "7J42F00" "7J42f00" "7J42G00" "7J42g00" "7J42H00" "7J42h00" "7J42J00" "7J42j00" "7J42K00" "7J42k00" "7J42L00" "7J42l00" "7J42M00" "7J42m00" "7J42N00" "7J42P00" "7J42Q00" "7J42R00" "7J42S00" "7J42T00" "7J42U00" "7J42V00" "7J42W00" "7J42X00" "7J42Y00" "7J42Z00" "7J43.00" "7J43.11" "7J43000" "7J43100" "7J43200" "7J43211" "7J43212" "7J43300" "7J43400" "7J43700" "7J43800" "7J43900" "7J43A00" "7J43C00" "7J43D00" "7J43y00" "7J43z00" "7K14.00" "7K14y00" "7K14z00" "7K15.00" "7K15y00" "7K15z00" "7K1D.00" "7K1D511" "7K1Dy00" "7K1Dz00" "7K1Ez00" "7K1F.00" "7K1F300" "7K1F400" "7K1F500" "7K1Fy00""7K1Fz00" "7K1G200" "7K1Gy11" "7K1H200" "7K1H300" "7K1H400" "7K1J300" "7K1JH00" "7K1Jy00" "7K1Jz00" "7K1K000" "7K1K200" "7K1K700" "7K1K800" "7K1K900""7K1KA00" "7K1KB00" "7K1KC00" "7K1Ky00" "7K1Kz00" "7K1L011" "7K1L100" "7K1L211" "7K1L300" "7K1La00" "7K1Lb00" "7K1Le00" "7K1Lg00" "7K1LZ00" "7K1N900" "7K1NA00" "7K1T100" "7K6GN00" "7K6H200" "7K6H411" "7K6H700" "7K6Hh00" "7K6HX00" "7P20100" "8HTo.00" "B585000" "N1y1.00" "N1y2.00" "N331.00" "N331.13" "N331000" "N331100" "N331200" "N331300" "N331400" "N331500" "N331600" "N331700" "N331800" "N331900" "N331A00" "N331B00" "N331C00" "N331M00" "N331M11" "N331N00" "N331y00" "N331z00" "N338.00" "N338000" "N338100" "N338111" "N338200" "N338300" "N338400" "N338500" "N338600" "N338z00" "NyuB000" "NyuB800" "NyuCE00" "S1...00" "S10..00" "S10..11" "S10..12" "S100.00" "S100.11" "S100.12" "S100000" "S100100" "S100111" "S100200" "S100211" "S100300" "S100311" "S100400" "S100411" "S100500" "S100511" "S100600" "S100611" "S100700" "S100711" "S100800" "S100900" "S100A00" "S100B00" "S100C00" "S100D00" "S100E00" "S100F00" "S100G00" "S100H00" "S100J00" "S100K00" "S100L00" "S100M00" "S100N00""S100x00" (cont.) |

**Table S3.** Continued.

| | | |
|---|---|---|
| **3** | 45 | (cont.) "S100z00" "S101.00" "S101.11" "S101.12" "S101000" "S101100" "S101111" "S101200" "S101211"<br>"S101300" "S101311" "S101400" "S101411" "S101500" "S101511" "S101600" "S101611" "S101700"<br>"S101711" "S101800" "S101900" "S101A00" "S101B00" "S101C00" "S101D00" "S101E00" "S101F00"<br>"S101G00" "S101H00" "S101J00" "S101K00" "S101L00" "S101M00" "S101N00" "S101x00" "S101z00"<br>"S102.00" "S102000" "S102100" "S102200" "S102300" "S102400" "S102500" "S102600" "S102y00"<br>"S102z00" "S103.00" "S103000" "S103100" "S103200" "S103300" "S103400" "S103500" "S103600"<br>"S104.00" "S104000" "S104100" "S104200" "S104300" "S104400" "S104500" "S104600" "S105.00"<br>"S105000" "S105100" "S105200" "S105300" "S105400" "S105500" "S105600" "S106.00" "S106000"<br>"S106100" "S107.00" "S107000" "S107100" "S108.00" "S109.00" "S10A.00" "S10A000" "S10A100"<br>"S10A200" "S10B.00" "S10B000" "S10B100" "S10B200" "S10B300" "S10B400" "S10B500" "S10B600"<br>"S10x.00" "S10y.00" "S10z.00" "S11..00" "S11..11" "S11..12" "S110.00" "S110000" "S110100" "S110200"<br>"S110300" "S110400" "S110600" "S110700" "S110800" "S110900" "S110A00" "S110z00" "S111.00"<br>"S111000" "S111100" "S111200" "S111300" "S111400" "S111600" "S111700" "S111800" "S111900"<br>"S111A00" "S111z00" "S112.00" "S112000" "S112100" "S112200" "S112300" "S112400" "S112600"<br>"S112700" "S112800" "S112900" "S112A00" "S112z00" "S113.00" "S113000" "S113100" "S113200"<br>"S113300" "S113400" "S113600" "S113700" "S113800" "S113900" "S113A00" "S113z00" "S114.00"<br>"S114000" "S114100" "S114200" "S114300" "S114400" "S114500" "S115.00" "S115000" "S115100"<br>"S115200" "S115300" "S115400" "S115500" "S115z00" "S116.00" "S116000" "S116100" "S116200"<br>"S116300" "S116z00" "S117.00" "S117000" "S117100" "S117200" "S117300" "S117z00" "S118.00"<br>"S118000" "S118100" "S118200" "S118300" "S118z00" "S119.00" "S119000" "S119100" "S119200"<br>"S119300" "S119z00" "S11x.00" "S11y.00" "S11z.00""S120.00" "S120000" "S120100" "S120200"<br>"S120300" "S120400" "S120500" "S120600" "S120700" "S120800" "S120900" "S120A00" "S120z00"<br>"S121.00" "S121000" "S121100" "S121200" "S121300" "S121400" "S121500" "S121600" "S121700"<br>"S121800" "S121900" "S121z00" "S122.00" "S123.00" "S125200" "S126200" "S127.00""S127000"<br>"S127100" "S128.00" "S12X.00" "S12X000" "S12X100" "S12y.00" "S12y000" "S12y100" "S12z.11"<br>"S12z.12" "S13..00" "S130.00" "S130000" "S130100" "S130200" "S130300" "S130400" "S130500"<br>"S130600" "S130y00" "S130z00" "S131.00" "S131000" "S131100" "S131200" "S131300" "S131400"<br>"S131500" "S131600" "S131y00" "S131z00" "S132.00" "S132000" "S132100" "S132200" "S132y00"<br>"S132z00" "S133.00" "S133000" "S133100" "S133200" "S133y00" "S133z00" "S134.00" "S134000"<br>"S134100" "S134300" "S134400" "S134500" "S134600" "S134700" "S134800" "S134z00" "S135.00"<br>"S135000" "S135100" "S135300" "S135400" "S135500" "S135600" "S135700" "S135800" "S135y00"<br>"S135z00" "S13y.00" "S13z.00" "S14..00" "S140.00" "S141.00" "S14z.00" "S15..00" "S150.00" "S150000"<br>"S150100" "S1z..00" "S2...00" "S2...11" "S20..00" "S20..11" "S200.00" "S200000" "S200100" "S200200"<br>"S200300" "S200z00" "S201.00" "S201000" "S201100" "S201200" "S201300" "S201z00" "S20z.00"<br>"S21..00" "S21..11" "S210.00" "S210000" "S210100" "S210200" "S210300" "S210400" "S210500"<br>"S210600" "S210z00" "S211.00" "S211000" "S211100" "S211200" "S211300" "S211400" "S211500"<br>"S211600" "S211z00" "S21z.00" "S22..00" "S220.00" "S220000" "S220100" "S220200" "S220300"<br>"S220400" "S220500" "S220600" "S220700" "S220z00" "S221.00" "S221.11" "S221000 "S221100"<br>"S221200" "S221300" "S221400" "S221500" "S221600" "S221700" "S221z00" "S222.00" "S222000"<br>"S222100" "S222z00" "S223.00" "S223000" "S223100" "S223z00" "S224.00" "S224.11" "S224000" (cont.) |

**Table S3.** Continued.

| | | |
|---|---|---|
| **3** | 45 | (cont.) "S224100" "S224200" "S224300" "S224400" "S224500" "S224600" "S224700" "S224800" "S224900" "S224x00" "S224z00" "S225.00" "S225.11" "S225000" "S225100" "S225200" "S225300" "S225400" "S225500" "S225600" "S225700" "S225800" "S225900" "S225x00" "S225z00" "S226.00" "S227.00" "S228.00" "S22z.00" "S23..00" "S23..11" "S230.00" "S230000" "S230100" "S230200" "S230300" "S230400" "S230500" "S230600" "S230700" "S230800" "S230900" "S230A00" "S230B00" "S230z00" "S231.00" "S231000" "S231100" "S231200" "S231300" "S231400" "S231500" "S231600" "S231700" "S231800" "S231900" "S231A00" "S231B00" "S231z00" "S232.00" "S232000" "S232100" "S232200" "S232300" "S232z00" "S233.00" "S233000" "S233100" "S233200" "S233300" "S233z00" "S234.00" "S234.11" "S234000" "S234100" "S234111" "S234200" "S234211" "S234300" "S234400" "S234500" "S234600" "S234700" "S234800" "S234900" "S234911" "S234912" "S234A00" "S234A11" "S234A12" "S234B00" "S234C00" "S234D00" "S234E00" "S234F00" "S234G00" "S234z00" "S235.00" "S235.11" "S235000" "S235100" "S235111" "S235200" "S235211" "S235300" "S235400" "S235500" "S235600" "S235700" "S235800" "S235900" "S235911" "S235912" "S235A00" "S235A11" "S235A12" "S235B00" "S235C00" "S235D00" "S235E00" "S235F00" "S235z00" "S236.00" "S237.00" "S238.00" "S239.00" "S23A.00" "S23B.00" "S23C.00" "S23x.00" "S23x000" "S23x100" "S23x111" "S23x200" "S23x211" "S23x300" "S23xz00" "S23y.00" "S23y000" "S23y100" "S23y200" "S23y300" "S23yz00" "S23z.00" "S24..00" "S24..11" "S240.00" "S240000" "S240100" "S240200" "S240300" "S240400" "S240500" "S240600" "S240700" "S240800" "S240900" "S240A00" "S240B00" "S240C00" "S240D00" "S240E00" "S240F00" "S240y00" "S240z00" "S241.00" "S241000" "S241100" "S241200" "S241300" "S241400" "S241500" "S241600" "S241700" "S241800" "S241900" "S241A00" "S241B00" "S241C00" "S241D00" "S241E00" "S241F00" "S241y00" "S241z00" "S242.00" "S242000" "S242100" "S242200" "S242300" "S24z.00" "S25..00" "S25..11" "S250.00" "S250000" "S250200" "S250300" "S250400" "S250500" "S250600" "S250700" "S250800" "S250A00" "S250B00" "S250C00" "S250x00" "S250z00" "S251.00" "S251000" "S251200" "S251300" "S251400" "S251500" "S251600" "S251700" "S251800" "S251A00" "S251B00" "S251C00" "S251x00" "S251z00" "S252.00" "S253.00" "S26..00" "S26..11" "S26..12" "S260.00" "S260000" "S260300" "S260400" "S260500" "S260600" "S260700" "S260800" "S260900" "S260A00" "S260B00" "S260C00" "S260D00" "S260E00" "S260F00" "S260G00" "S260H00" "S260J00" "S260K00" "S260L00" "S260M00" "S260N00" "S260P00" "S260Q00" "S260R00" "S260S00" "S260T00" "S260U00" "S260V00" "S260W00" "S260x00" "S260z00" "S261.00" "S261000" "S261300" "S261400" "S261500" "S261600" "S261700" "S261800" "S261900" "S261A00" "S261B00" "S261C00" "S261D00" "S261E00" "S261F00" "S261G00" "S261H00" "S261J00" "S261K00" "S261L00" "S261M00" "S261N00" "S261P00" "S261Q00" "S261R00" "S261S00" "S261T00" "S261U00" "S261V00" "S261W00" "S261x00" "S261z00" "S262.00" "S263.00" "S264.00" "S26z.00" "S27..00" "S270.00" "S271.00" "S27z.00" "S28..00" "S28..11" "S280.00" "S281.00" "S28z.00" "S29..11" "S29..12" "S29..13" "S292.00" "S292000" "S292100" "S293.00" "S294.00" "S294000" "S294100" "S2A..00" "S2B..00" "S2z..00" "S3...00" "S3...11" "S30..00" "S30..11" "S300.00" "S300100" "S300200" "S300300" "S300311" "S300400" "S300600" "S300700" "S300800" "S300900" "S300A00" "S300y00" "S300y11" "S300z00" "S301.00" "S301100" "S301200" "S301300" "S301311" "S301400" "S301500" "S301600" "S301700" "S301800" "S301900" "S301A00" "S301y00" "S301y11" "S301z00" "S302.00" "S302011" "S302012" "S302100" "S302200" "S302400" "S303.00" "S303011" "S303012" "S303100" "S303200" "S303300" "S303400" "S303z00" "S304.00" (cont.) |

**Table S3.** Continued.

| | | |
|---|---|---|
| **3** | 45 | (cont.) "S305.00" "S30w.00" "S30x.00" "S30y.00" "S30y.11" "S30z.00" "S31..00" "S310.00" "S310000" "S310011" "S310012" "S310100" "S310z00" "S311.00" "S311000" "S311100" "S311z00" "S312.00" "S312.11" "S312000" "S312100" "S312200" "S312300" "S312400" "S312500" "S312600" "S312x00" "S312z00" "S313.00" "S313.11" "S313000" "S313100" "S313200" "S313300" "S313400" "S313500" "S313600" "S313x00" "S313z00" "S314.00" "S315.00" "S31z.00" "S32..00" "S320.00" "S320000" "S320100" "S320200" "S320300" "S320400" "S321.00" "S321000" "S321100" "S321200" "S321300" "S321400" "S32z.00" "S33..00" "S330.00" "S330000" "S330011" "S330012" "S330100" "S330200" "S330300" "S330400" "S330500" "S330600" "S330700" "S330800" "S330900" "S330z00" "S331.00" "S331000" "S331011" "S331012" "S331100" "S331200" "S331300" "S331400" "S331500" "S331600" "S331700" "S331800" "S331900""S331A00" "S331z00" "S332.00" "S332000" "S332100" "S332200" "S332z00" "S333.00" "S333000" "S333100" "S333200" "S333z00" "S334.00" "S334000" "S334100" "S335.00" "S335000" "S335100" "S336.00" "S336000" "S337.00" "S338.00" "S339.00" "S339000" "S339100" "S33A.00" "S33B.00" "S33C.00" "S33x.00" "S33x.11" "S33x000" "S33x100" "S33x200" "S33xz00" "S33y.00" "S33y000" "S33y100" "S33y200" "S33yz00" "S33z.00" "S34..00" "S340.00" "S341.00" "S342.00" "S342000" "S342100" "S343.00" "S343000" "S343100" "S344.00" "S344.11" "S344.12" "S344000" "S344100" "S345.00" "S345000" "S345100" "S346.00" "S346000" "S346100" "S347.00" "S347000" "S347100" "S348.00" "S349.00" "S34x.00" "S34y.00" "S34z.00" "S35..00" "S35..11" "S35..12" "S350.00" "S350.11" "S350.12" "S350000" "S350100" "S351.00" "S351000" "S351100" "S352.00" "S352.11" "S352000" "S352100" "S352111" "S352200" "S352300" "S352400" "S352500" "S352600" "S352700" "S352800" "S352900" "S352A00" "S352B00" "S352C00" "S352D00" "S352E00" "S352F00" "S352G00" "S352H00" "S352J00" "S352z00" "S353.00" "S353000" "S353100" "S353111" "S353200" "S353300" "S353400" "S353500" "S353600" "S353700" "S353800" "S353900" "S353A00" "S353B00" "S353C00" "S353D00" "S353E00" "S353F00" "S353G00" "S353H00" "S353J00" "S353z00" "S354.00" "S355.00" "S356.00" "S35z.00" "S36..00" "S36..11" "S360.00" "S360000" "S360100" "S360200" "S360300" "S361.00" "S361000" "S361100" "S361200" "S361300" "S362.00" "S362000" "S362100" "S363.00" "S36z.00" "S37..00" "S370.00" "S371.00" "S3X..00" "S3x..00" "S3x0.00" "S3x1.00" "S3x2.00" "S3x3.00" "S3x4.00" "S3xz.00" "S3z..00" "S3z..11" "S3z0.00" "S3z0000" "S3z1.00" "S3z2.00" "S3zz.00" "S4...13" "S4A..00" "S4A0.00" "S4A0000" "S4A0100" "S4A1.00" "S4A1000" "S4A1100" "S4A2.00" "S4A2000" "S4A2100" "S4A3.00" "S4A3000" "S4A3100" "S4B..00" "S4B0.00" "S4B0000" "S4B0100" "S4B1.00" "S4B1000" "S4B1100" "S4B2.00" "S4B2000" "S4B2100" "S4B3.00" "S4B3000" "S4B3100" "S4C..00" "S4C0.00" "S4C0000" "S4C0100" "S4C0200" "S4C0300" "S4C0400" "S4C0500" "S4C0600" "S4C0y00" "S4C1.00" "S4C1000" "S4C1100" "S4C1200" "S4C1300" "S4C1400" "S4C1500" "S4C1600" "S4C1y00" "S4C2.00" "S4C2000" "S4C2100" "S4C2200" "S4C2300" "S4C2400" "S4C2500" "S4C2600" "S4C2y00" "S4C3.00" "S4C3000" "S4C3100" "S4C3200" "S4C3300" "S4C3400" "S4C3500" "S4C3600" "S4C3y00" "S4D..00" "S4D0.00" "S4D0000" "S4D0100" "S4D0200" "S4D0300" "S4D0400" "S4D0500" "S4D0600" "S4D1.00" "S4D1000" "S4D1100" "S4D1200" "S4D1300" "S4D1400" "S4D1500" "S4D1600" "S4D2.00" "S4D2000" "S4D2100" "S4D2200" "S4D2300" "S4D2400" "S4D2500" "S4D2600" "S4D3.00" "S4D3000" "S4D3100" "S4D3200" "S4D3300" "S4D3400" "S4D3500" "S4D3600" "S4E..00" "S4E0.00" "S4E1.00" "S4E2.00" "S4E3.00" "S4F..00" "S4F0.00" "S4F1.00" "S4F2.00" "S4F3.00" "S4F4.00" "S4F5.00" "S4F6.00" "S4F7.00" "S4G..00" "S4G0.00" "S4G1.00" "S4G2.00" "S4G3.00" "S4H..00" "S4H0.00" "S4H0000" (cont.) |

**Table S3.** Continued.

| | | |
|---|---|---|
| **3** | 45 | (cont.) "S4H0100" "S4H0200" "S4H0400" "S4H0600" "S4H1.00" "S4H1000" "S4H1100" "S4H1200" "S4H1300" "S4H1400" "S4H1600" "S4H2.00" "S4H2000" "S4H2100" "S4H2200" "S4H2400" "S4H2600" "S4H3.00" "S4H3000" "S4H3100" "S4H3200" "S4H3300" "S4H3400" "S4H3600" "S4J..00" "S4J0.00" "S4J0000" "S4J0100" "S4J1.00" "S4J1000" "S4J1100" "S4J1200" "S4J1300" "S4J2.00" "S4J2000" "S4J2100" "S4J3.00" "S4J3000" "S4J3100" "S4J3200" "S4J3300" "SC0X.00" "SC0z.11" "SC20.00" "SC3C000" "SC3D400" "SD92000" "SP04A00" "SP04C00" "SP22400" "SR1..00" "SR10.00" "SR10000" "SR10100" "SR11.00" "SR12.00" "SR12000" "SR12100" "SR13.00" "SR14.00" "SR14000" "SR14100" "SR15000" "SR15100" "SR16000" "SR16100" "SR1z.00" "SR1z000" "SR1z100" "Syu1500" "Syu1600" "Syu2700" "Syu2800" "Syu4200" "Syu4300" "Syu4400" "Syu5300" "Syu5400" "Syu6300" "Syu6400" "Syu6500" "Syu7200" "Syu8300" "Syu8D00" "Syu9400" "SyuA200" "SyuBB00" "SyuL100" "SyuL400" "TC7..00" "Z6G1900" "Zw01.00" "Zw02.00" "Zw02400" "Zw02500" "Zw02D00" "Zw02E00" "Zw02F00" "Zw02G00" "Zw02H00" "Zw02J00"<br>**Seizure:** "1473.00" "14On.00" "1B26.00" "1B27.00" "1O30.00" "282..13" "6110.00" "667..00" "6671.00" "6672.00" "6674.00" "6677.00" "6678.00" "6679.00" "667A.00" "667B.00" "667C.00" "667D.00" "667E.00" "667F.00" "667G.00" "667H.00" "667J.00" "667K.00" "667L.00" "667M.00" "667N.00" "667Q.00" "667R.00" "667S.00" "667T.00" "667V.00" "667W.00" "667Z.00" "67AF.00" "67IJ000" "8BIF.00" "8CE7.00" "8Hlp.00" "8IAg.00" "8IAh.00" "8IAi.00" "8IB2.00" "8IB3.00" "8IB4.00" "8T0L.00" "9N0r.00" "9N4V.00" "9Of..00" "9Of0.00" "9Of1.00" "9Of2.00" "9Of3.00" "9Of4.00" "9Of5.00" "9Of6.00" "9Of7.00" "E201500" "Eu05212" "Eu06013" "Eu10800" "Eu44511" "Eu80300" "F132100" "F132z12" "F25..00" "F250.00" "F250000" "F250100" "F250200" "F250300" "F250400" "F250y00" "F250z00" "F251.00" "F251000" "F251011" "F251100" "F251200" "F251300" "F251400" "F251500" "F251600" "F251y00" "F251z00" "F254.00" "F254000" "F254100" "F254200" "F254300" "F254500" "F254z00" "F255.00" "F255000" "F255011" "F255012" "F255100" "F255200" "F255300" "F255311" "F255400" "F255500" "F255600" "F255y00" "F255z00" "F257.00" "F25A.00" "F25B.00" "F25C.00" "F25D.00" "F25E.00" "F25F.00" "F25G.00" "F25H.00" "F25y.00" "F25y000" "F25y100" "F25y400" "F25yz00" "F25z.00" "Fyu5000" "Fyu5100" "Q480.12" "R003300" "R003400" "R003z11" "SC20000"<br>**Bipolar disorder:** "E11..11" "E114.00" "E114000" "E114100" "E114200" "E114300" "E114400" "E114500" "E114600" "E114z00" "E115.00" "E115000" "E115100" "E115200" "E115300" "E115400" "E115500" "E115600" "E115z00" "E116.00" "E116000" "E116100" "E116200" "E116300" "E116400" "E116500" "E116600" "E116z00" "E117.00" "E117000" "E117100" "E117200" "E117300" "E117400" "E117500" "E117600" "E117z00" "Eu30.11" "Eu31.00" "Eu31000" "Eu31100" "Eu31200" "Eu31300" "Eu31500" "Eu31600" "Eu31700" "Eu31800" "Eu31900" "Eu31911" "Eu31y00" "Eu31y11" "Eu31z00" "Eu34100" "ZRby100"<br>**Depression:** "12K8.00" "1465.00" "1B17.00" "1B17.11" "1B1U.00" "1B1U.11" "1BT..00" "1JJ..00" "1TC..00" "1TC0.00" "1TC1.00" "1TC2.00" "1TC3.00" "2257.00" "62T1.00" "6658000" "6659000" "6G00.00" "8BK0.00" "8CAa.00" "8HHq.00" "8HHq000" "9H90.00" "9H91.00" "9H92.00" "9HA0.00" "9k4..00" "9k40.00" "9kQ..00" "9kQ..11" "9Ov..00" "9Ov0.00" "9Ov1.00" "9Ov2.00" "9Ov3.00" "9Ov4.00" "E001300" "E002.00" "E002100" "E002z00" "E004300" "E02y300" "E11..12" "E112.00" "E112.11" (cont.) |

**Table S3.** Continued.

| | | | |
|---|---|---|---|
| **3** | 45 |  | (cont.) "E112.12" "E112.13" "E112.14" "E112000" "E112100" "E112200" "E112300" "E112400" "E112500" "E112600" "E112z00" "E113.00" "E113.11" "E113000" "E113100" "E113200" "E113300" "E113400" "E113500" "E113600" "E113700" "E113z00" "E11y200" "E11z200" "E130.00" "E130.11" "E135.00" "E200300" "E204.00" "E204.11" "E211200" "E291.00" "E2B..00" "E2B0.00" "E2B1.00" "Eu02z16" "Eu32.00" "Eu32.11" "Eu32.12" "Eu32.13" "Eu32000" "Eu32100" "Eu32200" "Eu32211" "Eu32212" "Eu32213" "Eu32300" "Eu32311" "Eu32312" "Eu32313" "Eu32314" "Eu32400" "Eu32500" "Eu32600" "Eu32700" "Eu32800" "Eu32B00" "Eu32y00" "Eu32y11" "Eu32y12" "Eu32z00" "Eu32z11" "Eu32z12" "Eu32z13" "Eu32z14" "Eu33.00" "Eu33.11" "Eu33.12" "Eu33.13" "Eu33.14" "Eu33000" "Eu33100" "Eu33200" "Eu33211" "Eu33212" "Eu33213" "Eu33214" "Eu33300" "Eu33311" "Eu33313" "Eu33314" "Eu33315" "Eu33316" "Eu33400" "Eu33y00" "Eu33z00" "Eu34111" "Eu34112" "Eu34113" "Eu34114" "Eu3y111" "Eu41200" "Eu41211" "Eu53011" "Eu53012" "Eu92000" "Eu33z11" |
| **3** | 46 | **CNS stimulants:** "R01BA02" "R01BA52" "R01BA51" "N02BE71" "R05DA20" "R06AB51" "N02BE01" "R01BA03" " "R01AA04" "C01CA06" "R01AB01" "N06BA04" "N06BA07" "R03DA04" "C01CA26" "R03DB04" "N06BC01" "N02BA01 (whenever in combination with caffeine)" "R06AA09" "N06BC01" "R06AA02" "R05DA04" "N02CA52 whenever in combination with caffeine" "N02AC54 whenever in combination with caffeine" | **Insomnia:** "1B1B.00" "1B1B.11" "1B1B000" "1B1B100" "1B1B200" "38D1.00" "8Q0..00" "9Ngt.00" "E274.00" "E274.12" "E274100" "E274111" "E274200" "Eu51000" "Fy00.00" "K5A2100" "R005.11" "R005100" "R005200" |
| **3** | 47 | **Antipsychotics:** see criteria 43 (except "N05AH02" "N05AH04" "N05AX12")<br>**Antiemetics:** "A03FA01" "N05AB04" "R06AD02" "R05DA20 whenever in combination with antiemetics" "R06AD52" "N02AB52 whenever in combination with promethazine" "N02BE71 whenever in combination with promethazine " | **Parkinson's disease:** "147F.00" "297A.00" "2987" "2987.11" "2994" "2994.11" "38GM.00" "8Hx0.00" "8T06.00" "8T06000" "9Nle.00" "A94y100" "Eu02300" "F11x900" "F12..00" "F121.00" "F121.11" "F123.00" "F124.00" "F12W.00" "F12X.00" "F12z.00" "F130300" "Fyu2000" "Fyu2100" "Fyu2200" "Fyu2900" "Fyu2B00" |
| **3** | 48 | **Aspirin:** "N02BA01" "N02AJ07" "N02BA51" "N06BC01 whenever in combination with aspirin"<br>**Non-COX2 non-steroidal anti-inflammatory drugs:** "M01AA01" "M01AB01" "M01AB02" "M01AB03" "M01AB05" "M01AB55" "M01AB08" "M01AB11" "M01AB15" "M01AB16" "M01AC01" "M01AC02" "M01AC05" "M01AC06" "M01AE01" "M01AE14" "M01AE02" "M01AE52" "M01AE03" "M01AE53" "M01AE17" "M01AE04" "M01AE05" "M01AE09" "M01AE11" "M01AG01" "M01AG02" "M01AX01" "M01AX04"<br>**Proton pump inhibitors:** "A02BC" "M01AE53" "A02BD" "M01AE52"<br>**Misoprostol:** "A02BB01" | **Gastric ulcers:** "14C1.00" "14C1.11" "14C1.12" "1956.00" "7612111" "7612500" "761D500" "761D600" "761J.00" "761J.11" "761J000" "761J100" "761J111" "761Jy00" "761Jz00" "7627.00" "7627000" "7627100" "7627200" "7627y00" "7627z00" "J080000" "J102000" "J11..00" "J110.00" "J110000" "J110100" "J110111" "J110200" "J110300" "J110400" "J110y00" "J110z00" "J111.00" "J111000" "J111100" "J111111" "J111200" "J111211" "J111300" "J111400" "J111y00" "J111z00" "J112.00" "J112z00" "J113.00" "J11y.00" "J11y000" "J11y100" "J11y200" "J11y300" "J11y400" "J11yy00" "J11yz00" "J11z.00" "J11z.12" "J12..00" "J120.00" "J120000" "J120100" "J120200" "J120300" "J120400" "J120y00" "J120z00" "J121.00" "J121000" "J121100" "J121111" "J121200" "J121211" "J121300" "J121400" "J121y00" "J121z00" "J122.00" "J124.00" "J125.00" "J125z00" "J126.00" "J12y.00" "J12y000" "J12y100" "J12y200" "J12y300" "J12y400" "J12yy00" "J12yz00" "J12z.00" "J13..00" "J130.00" "J130000" "J130100" "J130200" "J130300" "J130400" "J130y00" "J130z00" "J131.00" "J131000" "J131100" "J131200" "J131300" "J131400" "J131y00" "J131z00" "J13y.00" "J13y000" "J13y100" "J13y200" "J13y300" "J13y400" "J13yy00" "J13yz00" "J13z.00" "J14..00" "J14..12" "J14..13" "J14..15" "J140.00" "J140000" "J140100" "J140200" "J140300" "J140400" "J140y00" "J140z00" "J141.00" "J141000" "J141100" "J141200" "J141300" "J141400" "J141y00" "J141z00" "J14y.00" "J14y000" "J14y100" "J14y200" "J14y300" "J14y400" "J14yy00" "J14yz00" "J14z.00" "J17y800" "ZV12711" "ZV12712" "ZV12C00" |

**Table S3.** Continued.

| | | | |
|---|---|---|---|
| **3** | 49 | **Non-steroidal anti-inflammatory drugs:** "N02BA01" "N02AJ07" "N02BA51" "N06BC01 whenever in combination with aspirin" M01A" (excluding topical formulations, suppositories, eye drops, and plasters) | **Chronic kidney disease stages 4 and 5:** "1Z13.00" "1Z14.00" "1Z1H.00" "1Z1J.00" "1Z1K.00" "1Z1L.00" "K054.00" "K055.00" |
| **3** | 50 | **Alpha-1 blockers:** see criteria 7<br>**Estrogens:** "G03AA" "G03AB" "G03C" "G03F" "L02AA01" | |
| **3** | 51 | **Anticholinergic drugs to be avoided in men:** "R06AB01" "R06AA52" "R06AD52" "R01BA52" "R01BA51" "R06AB51" "R06AB04" "R06AA54" "R06AA04" "R06AX02" "N07CA52" "R06AA02" "N02BE71" "N07XX59" "R01BA02" "R05FB02" "R06AA09" "R05CA03" "R01AA04" "N05BB01" "R06AX07" "N04AA01" "N04AB02" "M03B C01" "N02B E01" "N06A A09" "N06CA01" "N06AA17" "N06AA04" "N06AA01" "N06AA12" "N06AA02" "N06AA10" "N06AB05" "N06AA11" "N06AA06" "N05AA01" "N05AH02" "N05AH01" "N05AH03" "N05AB03" "N05AC02" "N05AB06" "A03CB04" "C01BA03" "A07DA52" "A03CB02" "A03CB" "A06AB04" "A03BA04" "C02AA03" "A03CA02" "A03BA01" "A07DA01" "A04AD04" "A03BA01" "N02AA51" "A03AB05" "N05AB04" "R06AD02" "N02AB52" | **Urinary tract symptoms:** "14D4.00" "A32y300" "A981100" "A983100" "K15..00" "K150.00" "K151.00" "K151200" "K151z00" "K152.00" "K152000" "K152y00" "K152z00" "K153.11" "K154.00" "K154000" "K154100" "K154200" "K154300" "K154400" "K154500" "K154600" "K154700" "K154800" "K154z00" "K155.00" "K15y.00" "K15y000" "K15y100" "K15yz00" "K15z.00" "K213.00" "Kyu5000" "Kyu5100" "R081000" "R086.00" "R086z00" "1AZ6.00" "1AZ6000" "1AZ6100" "1AZ6200"<br>**Benign prostatic hyperplasia:** "25Q2.11" "K20..00" "K20..14" "K20..15" "K200.00" |
| **5** | 52 | **ACE inhibitors:** "C09AA01" "C09BA01" "C09AA02" "C09BA02" "C09AA03" "C09BA03" "C09AA04" "C09BA04" "C09AA05" "C09BB05" "C09AA06" "C09BA06" "C09AA08" "C09AA09" "C09AA10" "C09BB10" "C09AA13" "C09AA16"<br>**Amiloride and triamterene:** "C03DB01" "C03EB02"<br>Whenever in combination with amiloride or triamterene: "C03EA02" "C07DB01" "C03EA01" "C07DA06" "C03EB01" "C03DB02" | **Hypokalemia:** "44I4200" "C368.11" |
| **5** | 53 | **Anticholinergics:** see criteria 43 | |
| **5** | 54 | **Tricyclic antidepressants:** "N06AA09" "N06CA01" "N06AA02" "N06AA06" "N06AA12" "N06AA07" "N06AA17" "N06AA01" "N06AA10" "N06C" "N06AA11" "N06AA04" "N06AA13" "N06AA15" "N06AA16" "N06AA21"<br>**Selective serotonin reuptake inhibitors:** "N06AB"<br>**Additional central nervous system active drugs:** Opioids (see criteria 45), benzodiazepines (see criteria 43), non-benzodiazepine / benzodiazepine receptor agonist hypnotics (see criteria 22), antipsychotics (see criteria 17). | |
| **5** | 55 | **Antipsychotics:** see criteria 17<br>**Additional central nervous system active drugs:** opioids (see criteria 45), benzodiazepines (see criteria 43), non-benzodiazepine / benzodiazepine receptor agonist hypnotics (see criteria 22), selective serotonin reuptake inhibitors (see criteria 54), tricyclic antidepressants (see criteria 55). | |
| **5** | 56 | **Benzodiazepines**: see criteria 43<br>**Non-benzodiazepine / benzodiazepine receptor agonist hypnotics:** see criteria 22<br>(cont.) | |

**Table S3.** Continued.

| | | | |
|---|---|---|---|
| **5** | 56 | (cont.)<br>**Additional central nervous system active drugs:** opioids (see criteria 45), selective serotonin reuptake inhibitors (see criteria 54), tricyclic antidepressants (see criteria 55), antipsychotics (see criteria 17). | |
| **5** | 57 | **Corticosteroids:** see criteria 43<br>**Non-steroid anti-inflammatory drugs:** see criteria 49<br>**PPI:** see criteria 48<br>**Misoprostol:** see criteria 48 | |
| **5** | 58 | **Lithium:** "N05AN01"<br>**Angiotensin-converting enzyme:** see criteria 12 | **Lithium monitoring:** "44vE.00" "44W8.00" "44W8.11" "44W8000" "44W8100" "44W8200" "6657.11" "665J.00" "R105300" |
| **5** | 59 | **Lithium:** see criteria 58<br>**Loop diuretics:** "C03CA" "C03EB02" "C03DB01 whenever in combination with loop diuretic" "C03CB02" "C03CC01" "C03EB01" "C07CA23 whenever in combination with loop diuretic " "C03CB01" | **Lithium monitoring:** see criteria 58 |
| **5** | 60 | **Opioids:** see criteria 45<br>**Additional central nervous system active drugs:** selective serotonin reuptake inhibitors (see criteria 54), tricyclic antidepressants (see criteria 55), antipsychotics (see criteria 17), benzodiazepines (see criteria 43), non-benzodiazepine / benzodiazepine receptor agonist hypnotics (see criteria 22) | |
| **5** | 61 | **Loop diuretics:** see criteria 59<br>**Peripheral alpha-1 blockers:** see criteria 7<br>**Angiotensin-converting enzyme:** see criteria 12<br>**Angiotensin receptor blockers:** see criteria 12<br>**Calcium-channel blockers:** see criteria 11<br>**Thiazide-like diuretics:** "C03A" | |
| **5** | 62 | **Theophylline:** "R03DA04" "C01CA26 whenever in combination with theophylline" "R03DB04"<br>**Cimetidine:** "A02BA01" "A02BA51" | |
| **5** | 63 | **Warfarin:** "B01AA03"<br>**Amiodarone:** "C01BD01" | **Warfarin monitoring and International normalized ratio monitoring:** "4130.00" "41C5.00" "42jr.00" "42QE.00" "42QE000" "42QE100" "42QE200" "66Q..00" "66Q7.00" "66Q7000" "66Q8.00" "66Q8000" "66QE.00" "66QG.00" "8HHW.00" "9k21.00" "9k22.00" "9k22.11" "9k25.00" "9k25.11" |
| **5** | 64 | **Warfarin:** see criteria 63<br>**Non-steroidal anti-inflammatory drugs:** M01A" (excluding topical formulations, suppositories, eye drops, and plasters) | **Warfarin monitoring and International normalized ratio monitoring:** see criteria 63 |
| **6** | 65 | **Amiodarone:** see criteria 63 | **Chronic kidney disease stages 4 and 5:** see criteria 49 |
| **6** | 66 | **Apixaban:** "B01AF02" | **Chronic kidney disease stage 5**: "1Z14.00" "1Z1K.00" "1Z1L.00" "K055.00" |
| **6** | 67 | **Dabigatran:** "B01AE07" | **Chronic kidney disease stages 4 and 5:** see criteria 49 |

**Table S3.** Continued.

| | | | |
|---|---|---|---|
| **6** | 68 | **Edoxaban:** "B01AF03" | **Chronic kidney disease stages 4 and 5:** see criteria 49 |
| **6** | 69 | **Fondaparinux:** "B01AX05" | **Chronic kidney disease stages 4 and 5:** see criteria 49 |
| **6** | 70 | **Rivaroxaban:** "B01AF01" | **Chronic kidney disease stages 4 and 5:** see criteria 49 |
| **6** | 71 | **Spironolactone:** "C03DA01" "C03EB01" | **Chronic kidney disease stages 4 and 5:** see criteria 49 |
| **6** | 72 | **Triamterene:** "C03DB02" "C03EA06 whenever in combination with triamterene" "C03EB01 whenever in combination with triamterene " | **Chronic kidney disease stages 4 and 5:** see criteria 49 |
| **6** | 73 | **Duloxetine:** "N06AX21" | **Chronic kidney disease stages 4 and 5:** see criteria 49 |
| **6** | 74 | **Tramadol:** "N02AX20" | **Chronic kidney disease stages 4 and 5:** see criteria 49 |
| **6** | 75 | **Probenecid:** "M04AB01" | **Chronic kidney disease stages 4 and 5:** see criteria 49 |

[1] Original table number from the 2015 Beers criteria (doi: 10.1111/jgs.13702)

**Table S4.** List of Anatomical therapeutic chemical (ATC) classification codes and Read codes used to assess potential inappropriate prescription (PIP) according to the Prescribing Optimally in Middle-aged People's Treatments (PROMPT) criteria.

| Criteria Number | ATC codes used | Read codes used |
|---|---|---|
| 1 | **Opioids:** "N02AA01" "N02AA51" "N01AB02" "N02AA02" "N02AA03" "N02AA05" "N02AA55" "N02AJ" "N02AA08" "N02AA10" "N02AB02" "N02AB52" "N01AH01" "N02AB03" "N01AH51" "N02AD01" "N02BE01" "N02AC54" "N02AC01" "N02AC04" "N02AD02" "N07BC01" "N02AE01" "N07BC51" "N02AF02" "N02AX02" "N02AX"<br>**Stimulant laxatives:** "A06AB02" "A06AA02 (whenever in combination with a stimulant laxative)" "A06 C01" "A06AB06" "P02CB01" "A06A B58" "A06AB08" "A06AB05" "A06AB07" "A06AB53" "A06AA51" "A06AB04" "A06AD04"<br>**Other laxative drugs:** "A06AC01" "A03AA04" "A06AC03" "A06AC53" "A06AC06" "A06AD15" "A06AD" "A11G (whenever in combination with macrogol)" "A06A G06" "A06AA02" "A06AG10 " "A06AA51" "A06AA01" "A06AX" | **Zollinger Ellison Syndrome:** C115.11 |
| 2 | **Proton pump inhibitors:** "A02BC" "M01AE53" "A02BD" "M01AE52" | |
| 3 | **Esomeprazole and omeprazole:** "A02BC01" "M01AE53" "A02BC05" "M01AE52"<br>**Clopidogrel:** "B01AC04" | |
| 4 | **Alpha-adrenoreceptor antagonists:** "C02CA04" "C02CA01" "G04CA03"<br>**Other antihypertensive drugs:** "C02 except C02CA" "C09AA" "C09C" "C09D" "C08C" "C08D" "C03A" "C03B" "C03C" "C03D" "C03E" "C03XA" "C07" "C09XA02" | **Hypertension:** "14A2.00" "661M600" "661N600" "662..12" "6627.00" "6628.00" "662b.00" "662c.00" "662d.00" "662F.00" "662G.00" "662O.00" "662P.00" "662P000" "662P100" "662q.00" "8B26.00" "8BL0.00" "8CR4.00" "8HT5.00" "8I3N.00" "8IA6.00" "9OI1.00" "9OI3.00" "9OI4.00" "9OI5.00" "9OI6.00" "9OI7.00" "9OI8.00" "9OIA.11" "9OIZ.00" "G2...00" "G2...11" "G20..00" "G20..12" "G200.00" "G201.00" "G202.00" "G203.00" "G20z.00" "G20z.11" "G21..00" "G210.00" "G210000" "G210100" "G210z00" "G211.00" "G211000" "G211100" "G211z00" "G21z.00" "G21z000" "G21z011" "G21z100" "G21zz00" "G22..00" "G220.00" "G221.00" "G222.00" "G22z.00" "G22z.11" "G23..00" "G230.00" "G231.00" "G232.00" "G233.00" "G234.00" "G23z.00" "G24..00" "G240.00" "G240000" "G240z00" "G241.00" "G241000" "G241z00" "G244.00" "G24z.00" "G24z000" "G24z100" "G24zz00" "G25..00" "G25..11" "G250.00" "G251.00" "G26..00" "G26..11" "G27..00" "G28..00" "G2y..00" "G2z..00" "G672.11" "G8y3.00" "Gyu2.00" "Gyu2000" "Gyu2100" "L12..00" "L122.00" "L122000" "L122100" "L122300" "L122400" "L122z00" "L127.00" "L127000" "L127100" "L127200" "L127300" "L127400" "L127z00" "L128.00" "L128000" "L128100" "L128200" "L12B.00" "Lyu1.00" |
| 5 | **Aspirin:** "B06AC06" | |
| 6 | **Calcium-channel blockers:** "C08DB01" "C08DA01" "C09BB10"<br>**Beta blockers:** "C07AA" "C07AB" "C07AG" "C07BA" "C07BB" "C07C" "C07D" | |
| 7 | **Dipyridamole:** "B01AC07"<br>**Other antiplatelet medication:** "B01AC04" "B01AC05" "B01AC24" "B01AC22" "B01AC23" "B01AC06" |  |

**Table S4.** Continued.

| | | |
|---|---|---|
| **8** | **First generation anti-histamines (oral formulations):** "R06AB01" "R06AB51" "R06AB04" "R06AA54" "R06AA52" "R06AA04" "R06AX02" "R01BA52" "R06A A02" "R06AA52" "R01BA02" "R05DA20" "R05FB02" "R06AA09" "R05CA03" "R01AA04" "N05BB01" "R06AE05" "R06AD02" "R06AD52" "R06AX07" "R06AD01" "R06AD07" "R01BA01" "R01BA51" "R06AB05" "R06AC01"<br>**Other oral anti-histamine drugs:** "R06AX13" "R06AX27" "R06AX26" "R06AE07" "R06AE09" "R06AX18" "R06AX25" | |
| **9** | **Theophylline:** "R03DA04" "R03DA02" "R03DB04" "R03DA54" "R03DA74" | |
| **10** | **Oral corticosteroids:** "H02AB01" "H02AB02" "H02AB04" "H02AB06" "H02AB07" "H02AB08" "H02AB09" "A01AC03" "H02AB10" "H02AB13" "A07EA07"<br>**Bisphosphonates:** "M05BA01" "M05BB01" "M05BA02" "M05BA03" "M05BB03" "M05BA04" "M05BA05" "M05BA06" "M05BA08" "M05BA07" "M05BB04" "M05BB02" | |
| **11** | **Mucolytic agents:** "R05CB" | **Chronic obstructive pulmonary disease:** "66YB.00" "66Yg.00" "66YL.00" "66YL.11" "679V.00" "8CR1.00" "9Oi3.00" "9Oi4.00" "H3...00" "H3...11" "H32..00" "H322.00" "H36..00" "H37..00" "H38..00" "H3y..00" "H3z..00"<br>**Exacerbations:** "8BP8.00" "H312200" "H3y1.00" |
| **12** | **Venlafaxine:** "N06AX16"<br>**Selective serotonin reuptake inhibitors:** "N06AB" | |
| **13** | **Tricyclic antidepressants:** "N06AA09" "N06CA01" "N06AA02" "N06AA06" "N06AA12" "N06AA07" "N06AA17" "N06AA01" "N06AA10" "N06C" "N06AA11" "N06AA04" "N06AA13" "N06AA15" "N06AA16" "N06AA21"<br>**Other antidepressants:** "N06AX" "N06AB" "N06AG" "N06AF" | **Depression:** "12K8.00" "1465.00" "1B17.00" "1B17.11" "1B1U.00" "1B1U.11" "1BT..00" "1JJ..00" "1TC..00" "1TC0.00" "1TC1.00" "1TC2.00" "1TC3.00" "2257.00" "62T1.00" "6658000" "6659000" "6G00.00" "8BK0.00" "8CAa.00" "8HHq.00" "8HHq000" "9H90.00" "9H91.00" "9H92.00" "9HA0.00" "9k4..00" "9k40.00" "9kQ..00" "9kQ..11" "9Ov..00" "9Ov0.00" "9Ov1.00" "9Ov2.00" "9Ov3.00" "9Ov4.00" "E001300" "E002.00" "E002100" "E002z00" "E004300" "E02y300" "E11..12" "E112.00" "E112.11" "E112.12" "E112.13" "E112.14" "E112000" "E112100" "E112200" "E112300" "E112400" "E112500" "E112600" "E112z00" "E113.00" "E113.11" "E113000" "E113100" "E113200" "E113300" "E113400" "E113500" "E113600" "E113700" "E113z00" "E11y200" "E11z200" "E130.00" "E130.11" "E135.00" "E200300" "E204.00" "E204.11" "E211200" "E291.00" "E2B..00" "E2B0.00" "E2B1.00" "Eu02z16" "Eu32.00" "Eu32.11" "Eu32.12" "Eu32.13" "Eu32000" "Eu32100" "Eu32200" "Eu32211" "Eu32212" "Eu32213" "Eu32300" "Eu32311" "Eu32312" "Eu32313" "Eu32314" "Eu32400" "Eu32500" "Eu32600" "Eu32700" "Eu32800" "Eu32B00" "Eu32y00" "Eu32y11" "Eu32y12" "Eu32z00" "Eu32z11" "Eu32z12" "Eu32z13" "Eu32z14" "Eu33.00" "Eu33.11" "Eu33.12" "Eu33.13" "Eu33.14" "Eu33000" "Eu33100" "Eu33200" "Eu33211" "Eu33212" "Eu33213" "Eu33214" "Eu33300" "Eu33311" "Eu33313" "Eu33314" "Eu33315" "Eu33316" "Eu33400" "Eu33y00""Eu33z00" "Eu34111" "Eu34112" "Eu34113" "Eu34114" "Eu3y111" "Eu41200" "Eu41211" "Eu53011" "Eu53012" "Eu92000" "Eu33z11" |
| **14** | **Benzodiazepines:** "N05BA12" "N05CD11" "N05BA06" "N05CD06" "N05BA04" "N05CD07" "N05CD05" "N05BA08" "N05BA09" "N05BA05" "N05BA02" "N06CA01" "A03CA02" "N03AE01" "N05BA01" "N05CD03" "N05CD01" "N05CD02" "N05BA11" "N05CD08" "N05BA03" "N05BA10" | |
| **15** | **Non-benzodiazepine hypnotics:** "N05CF01" "N05CF02" "N05CF03" | |

**Table S4.** Continued.

| | |
|---|---|
| **16** | **Carbamazepine:** "N03AF01"<br>**Erythromycin:** "J01FA01"<br>**Clarithromycin:** "J01FA09" "A02BD14" "A02BD12" "A02BD06" "A02BD07" "A02BD05" "A02BD07" "A02BD09" "A02BD11" |
| **17** | **Strong opioids:** "N02AA01" "R05FA02" "N02AA51" "N01AB02 (whenever formulation has a combination with morphine)" "N02AA03" "N02AA05" "N02AA55" "N02AB02" "N02AB52" "N01AH01" "N02AB03" "N01AH51" "N02AD01" "N02BE01" "N07BC01" "N02AE01" "N07BC51" "N02AF02" "N07BC06"<br>**Laxatives:** "A06AB02" "A06AA02 (whenever in combination with a stimulant laxative)" "A06AC01" "A06AB06" "P02CB01" "A06AB58" "A06AB08" "A06AB05" "A06AB07" "A06AB53" "A06AA51" "A06AB04" "A06AD04" "A06AC01" "A03AA04" "A06AC03" "A06AC53" "A06AC06" "A06AD15" "A06AD" "A11G (whenever in combination with macrogol)" "A06A G06" "A06AA02" "A06AG10 " "A06AA51" "A06AA01" "A06AX" |
| **18** | **Nitrofurantoin:** "J01XE01" |
| **19** | **Long-acting Sulfonylureas:** "A10BB01" "A10BB02" |
| **20** | **Non-steroidal anti-inflammatory drugs:** M01A" (excluding topical formulations, suppositories, eye drops, and plasters) |
| **21** | **Non-steroidal anti-inflammatory drugs:** see criteria 20<br>**Low dose aspirin:** "B06AC06"<br>**H2-receptor antagonist:** "A02BA"<br>**Selective serotonin reuptake inhibitors:** "N06AB"<br>**Proton pump inhibitors: "**A02BC" "M01AE53" "A02BD" "M01AE52" |
| **22** | **Selective serotonin reuptake inhibitors:** "N06AB"<br>**Proton pump inhibitors: "**A02BC" "M01AE53" "A02BD" "M01AE52"<br>**Opioids:** "N02AA01" "N02AA51" "N01AB02" "N02AA02" "N02AA03" "N02AA05" "N02AA55" "N02AJ" "N02AA08" "N02AA10" "N02AB02" "N02AB52" "N01AH01" "N02AB03" "N01AH51" "N02AD01" "N02BE01" "N02AC54" "N02AC01" "N02AC04" "N02AD02" "N07BC01" "N02AE01" "N07BC51" "N02A F02" "N02AX02" "N02AX" |

**Abbreviations:** ATC: Anatomical therapeutic chemical;

### References[MLF1]


1. Corticosteroid Conversion Calculator - ClinCalc.com. Accessed April 11, 2023. https://clincalc.com/corticosteroids/

2. Kuenzig ME, Rezaie A, Kaplan GG, et al. Budesonide for the Induction and Maintenance of Remission in Crohn's Disease: Systematic Review and Meta-Analysis for the Cochrane Collaboration. *J Can Assoc Gastroenterol*. 2018;1(4):159-173. doi:10.1093/jcag/gwy018

3. Reilev M, Hallas J, Thomsen Ernst M, Nielsen GL, Bonderup OK. Long-term oral budesonide treatment and risk of osteoporotic fractures in patients with microscopic colitis. *Aliment Pharmacol Ther*. 2020;51(6):644-651. doi:10.1111/apt.15648

4. Chronic pain (primary and secondary) in over 16s: assessment of all chronic pain and management of chronic primary pain. *Chronic Pain*.

5. Ble A, Masoli JAH, Barry HE, et al. Any versus long-term prescribing of high risk medications in older people using 2012 Beers Criteria: results from three cross-sectional samples of primary care records for 2003/4, 2007/8 and 2011/12. *BMC Geriatr*. 2015;15(1):146. doi:10.1186/s12877-015-0143-8

6. ekpg_nitrofurantoin-safety-monitoring-guidance-prescribers-final-jan2020.pdf. Accessed April 13, 2023. http://www.eastkentformulary.nhs.uk/media/1538/ekpg_nitrofurantoin-safety-monitoring-guidance-prescribers-final-jan2020.pdf

7. Atrial fibrillation: diagnosis and management. *Atr Fibrillation*. Published online 2021.

8. Chronic heart failure in adults: diagnosis and management.

9. depression-in-adults-treatment-and-management-pdf-66143832307909.pdf. Accessed April 20, 2023. https://www.nice.org.uk/guidance/ng222/resources/depression-in-adults-treatment-and-management-pdf-66143832307909

10. Overview | Neuropathic pain in adults: pharmacological management in non-specialist settings | Guidance | NICE. Published November 20, 2013. Accessed April 14, 2023. https://www.nice.org.uk/guidance/cg173

11. hypertension-in-adults-pdf-2098552495813.pdf. Accessed April 18, 2023. https://www.nice.org.uk/guidance/qs28/resources/hypertension-in-adults-pdf-2098552495813
